# Statin Use And The Risk Of Venous Thromboembolism In Women Taking Hormone Therapy

**DOI:** 10.1101/2023.05.20.23290202

**Authors:** John W. Davis, Susan C. Weller, Laura Porterfield, Lu Chen, Gregg Wilkinson

**Author notes:** Corresponding Author: Susan C. Weller, Dept Population Health Science, School of Public and Population Health, University of Texas Medical Branch, 300 Harborside Dr., Galveston, Tx 77555-1153.

## Abstract

**Importance:** While hormone therapy (HT) in perimenopausal women can increase risk for venous thromboembolism (VTE), it is unclear to what extent statins may reduce HT-related risk.

**Objective:** To estimate VTE risk in 50–64-year-old women taking HT with or without statins and by statin intensity.

**Design:** A nested case-control study.

**Setting:** A US, commercially insured claims database.

**Participants:** Women aged 50-64 with at least one year of continuous membership between 2008-2019.

**Exposure:** Filled prescriptions for estrogens, progestogens, and statins were recorded in 12 months prior to index. Recent HT was defined as any estrogen or progestogen exposure within 60 days before the index date. Current statin exposure was defined as 90 or more days of continuous exposure prior to and including the index date. Statin intensity was defined by the statin exposure 30 days prior to index.

**Main Outcomes and Measures:** Cases were identified with VTE diagnoses (ICD codes) preceded by at least 12 months without VTE and followed within 30 days by anticoagulation, an inferior vena cava filter placement, or death. Controls were matched to cases (10:1) on date and age. Conditional logistic regression models estimated risk for HT and statin exposures with odds ratios (OR), adjusted for comorbidities.

**Results:** The total sample (mean age 57.5) included 20,359 cases and 203,590 matched controls; 9% had recent HT exposure and 16% had current statin exposure. In adjusted models, the OR for any recent HT exposure was 1.51 (95%CI:1.43-1.60) compared to no recent HT exposure. The OR for current statin therapy was 0.88 (95%CI:0.84-0.93) compared to no current statin exposure. The OR was 1.53 (95%CI:1.44-1.63) for those with HT without statins, 1.25 (95%CI:1.10-1.43) with HT with statins, and 0.89 (95%CI:0.85-0.94) with statins without HT compared to no recent HT and no current statins. HT with statin therapy had a 18% significantly lower OR than HT without statins (OR=0.82,95%CI:0.71-0.94) and greater risk reduction with higher intensity statins.

**Conclusions and Relevance:** In this case-control study, statin therapy was associated with reduced risk of VTE in women taking HT with greater risk reduction with high-intensity statins.

**Key Points:** *Question:* What is the impact of statin therapy in perimenopausal women exposed to hormone therapy?

*Findings:* In this case-control study of approximately 225,000 women aged 50-64, the VTE odds ratio was 53% higher in women recently exposed to HT without current statin therapy and 25% higher with recent HT exposure and current statin therapy compared to women without recent HT and current statin exposures,

*Meaning:* These findings suggest statins may reduce risk of VTE in women exposed to HT and further, HT may not be contraindicated in women taking statins.

## INTRODUCTION

Menopause-associated symptoms, such as ‘hot flashes’, vaginal dryness, disruptions in sleep patterns, and cognitive changes, are common and can affect quality of life.^1–3^ Hormone therapy (HT) is an effective treatment for many of these symptoms.^4,5^ However, concern for venous thromboembolism (VTE), stroke, or myocardial infarction (MI) risk prevents many symptomatic women from receiving HT.^6–8^ HT may double the risk for VTE,^9–11^ although the clinical trials were conducted with oral, conjugated equine estrogen (CEE)^9,12^ and newer studies suggest risk may be lower with other types of estrogen,^13–17^ routes of administration,^15,17–19^ and earlier initiation of therapy.^20–22^

In the decades since HT trials were conducted, statin therapy has shown efficacy in reducing risk for major cardiovascular events^23,24^ and VTE,^25,26^ but few studies have estimated statin’s effect on HT-associated VTE risk. Studies of concomitant HT and statin therapy suggest that the anti-inflammatory and anti-thrombotic effects of statin therapy may mediate risk of adverse cardiovascular events associated with HT. A UK study (1987-2008) in post-menopausal women found VTE risk from oral HT exposure for those not on statin therapy was 51% higher and for those on statin therapy was 21% higher than for those not exposed to oral HT.^27^ Similar reductions have been found for other adverse HT outcomes.^28, 29, 30^

Studies suggesting statins may reduce the increased risk of VTE associated with HT need validation in the US population because of different formulary, prescribing patterns, and higher VTE incidence. VTE incidence in the UK ranged from 150-175 cases (2017-2019), while US rates were 214 cases per 100,000 person-years (2015-2019).^31^ Also, statin use increased from 16% (2002-2003) to 26% (2012-2013) in US women over 40,^32^ while estrogen products (oral, vaginal, and transdermal) in women over 50 decreased from approximately 12.5% (2006) to 9.2% (2015) with larger reductions in oral estrogen products.^33^ Thus, this study explored the possible mitigating effect of statin therapy on VTE risk in US women 50-64 years of age exposed to exogenous hormones.

## METHODS

### Dataset and Cohort Selection

This nested case-control study was designed within Optum’s de-identified Clinformatics® Data Mart Database containing claims for approximately 62 million unique enrollees. Excluding Medicare Advantage subscribers, there are approximately 15 million annual members, a third of whom have continuous enrollment for ≥3 years. Because VTE incidence is relatively low, Clinformatics® is one of the few US databases sufficiently large to study VTE risk factors. The database contains information on outpatient/inpatient diagnoses and filled prescriptions. The age and sex distribution in the database is similar to the US population, but minorities are slightly underrepresented (13% vs. 19% Hispanic and 10% vs. 14% Black).^34^

The study cohort was limited to women 50-64 years of age with >1years of continuous enrollment between 1/1/2007-12/31/2019. Observations without sex (0.01%) or age (0.00007%) were excluded. Cases and matched controls were selected between 1/1/2008-12/1/2019 to allow a 12-month lookback period and 30-day follow-up period. The age range was selected because of the average age at menopause,^35^ higher rates of menopausal HT in this range,^33^ and the tendency to transition from commercial insurance to Medicare at 65. The study focused on statin therapy and HT as part of a larger project on VTE (design details, including ICD and CPT codes, reported previously,^15^and following STROBE guidelines).^36^ The University of Texas Medical Branch Institutional Review Board approved this study and waived informed consent because the data were de-identified.

### Cases and Controls

Cases were defined as women with a VTE diagnostic code during the observation period followed by a prescription for an anti-coagulant (excluding heparin flushes), intravascular vena cava filter, or death within 30 days of the VTE diagnosis (index date).^31^ Controls were randomly sampled (without replacement) and matched to cases by index date (month) and age (+/-2 years) at a 10:1 ratio. A large ratio was used in order to estimate low-frequency categories. Women were excluded with < 12 months of enrollment data, an acute or chronic VTE diagnosis, or an IVC filter placement in the year prior to the index date; or exposure to anticoagulants within14 days prior to the index date.

### HT Exposure

HT exposure was defined by a filled prescription for any estrogen and/or progestogen and the duration of the prescription (30 or 90 days). Exposure was evaluated regardless of route (oral/non-oral) or indication (e.g., menopausal HT or contraception). Timing of HT exposure was defined as recent (≤60 days before index) or past/none, because previous research indicated VTE risk diminished after 60 days.^15^ National Drug Codes (NDCs) for hormones were identified using Red Book^37^ and/or from a list supplied by the Food and Drug Administration^33^ (list published previously^15^).

### Statin Exposure

Statin exposure was defined by duration and continuity of filled prescriptions. Current statin exposure was defined as continuous filled prescriptions for ≥90 days prior to and including the index date. The comparison category was shorter, discontinuous, or no exposure in the past year. Intensity of therapy 30 days prior to the index date was defined as high (atorvastatin ≥40mg or rosuvastatin ≥20mg) or low/moderate (all other doses) according to AHA guidelines.^38^ Red Book^37^ NDCs were used to identify statin prescriptions, formulations, and doses (**Appendix**).

### Statistical Analysis

Risk for VTE with statin and/or HT exposures was estimated with adjusted odds ratios (ORs) and 95% confidence intervals.^39^ Because of the matched design, conditional logistic regression models were used.^17,27,30^ Covariates included region of residence, age at index date, history of cancer (except non-melanoma skin cancer; none/nonmetastatic/metastatic), history of prothrombotic conditions or other types of thrombosis (i.e., superficial thrombophlebitis), and varicose veins within the previous year; and within 30 days of the index date: hospitalization/surgery or trauma. In addition, coronary artery disease, stroke, lipid disorders, and smoking within the past year were included, as they may affect likelihood of prescribing HT or statin therapy. Finally, the Elixhauser set of comorbidities was included to control for general health status.^40^ The Elixhauser index has been validated for predicting mortality and includes chronic conditions such as obesity, liver disease, diabetes, etc.^41^ Known VTE risk factors (cancers and hypercoagulable conditions) were removed from the index and used as separate covariates. The final Elixhauser comorbidity index was summed and coded into terciles: 0, 1-2, and 3+ comorbidities.

Models estimated the association of HT and statin exposures on VTE risk, controlling for covariates. Sensitivity analyses examined the stability of main effects across stratified subsamples with different comorbidity burdens. Because conditional and unconditional analyses can yield similar results when few matching criteria are used,^42^ unconditional analyses were used on stratified subsamples to avoid loss of observations. An interaction term in the full sample with a conditional analysis assessed whether effect modification occurred between statin and HT exposures.

To estimate the combined effect of current statin and recent HT exposures across the distribution of risk factors, women were coded into four groups by their HT and statin exposure status. ORs were estimated for HT exposure without statin therapy, HT exposure with statin therapy, and statin therapy without HT exposure with reference to women with neither recent HT nor current statin therapy. Another model estimated the effect of statin intensity with additional subgroups for lower and higher intensity therapy.

## RESULTS

### Sample Characteristics

Of the 74,600 patients identified with a first, acute VTE and 22,380,610 eligible controls in the Clinformatics® database, we identified 20,359 cases that met inclusion criteria, and all were successfully matched to controls (n=203,590; flow chart for sample selection published previously^15^). Women in the study sample were approximately equally distributed in five-year age intervals (mean age 57.5). Cases had a higher proportion of comorbidities and VTE risk factors than controls (**Table 1**): 56% of cases and 22% of controls had three or more comorbidities, 26% vs. 5% had cancer, 13% vs. 5% had CAD, and 23% vs. 9% smoked. Among the VTE cases, 54% (n=10,995) were pulmonary emboli (PE) (with or without deep vein thrombosis, DVT), 46% (n=9,364) were DVT without PE, and 6% (n=1,197) died within 30 days following diagnosis.

**Table 1.** Sample Description.

| Variable | VTE Cases (n=20,359) | Comparators (n=203,590) |
| --- | --- | --- |
| **VTE Index Date** |  |  |
| 2007-2010 | 24.32% (4951) | 24.32% (49510) |
| 2011-2014 | 33.14% (6747) | 33.14% (67470) |
| 2015-2019 | 42.54% (8661) | 42.54% (86610) |
| **Age** |  |  |
| 50-54 | 33.35% (6789) | 34.84% (70927) |
| 55-60 | 34.80% (7084) | 33.52% (68237) |
| 61-65 | 31.86% (6486) | 31.64% (64426) |
| **Region** |  |  |
| Northeast | 8.28% (1686) | 8.85% (18010) |
| Midwest | 27.81% (5661) | 25.55% (52021) |
| South | 43.77% (8912) | 44.77% (91138) |
| West | 19.98% (4067) | 20.11% (40947) |
| Unknown | 0.16% (33) | 0.72% (1474) |
| **Comorbidities** |  |  |
| None | 13.81% (2812) | 37.32% (75981) |
| 1-2 | 30.25% (6158) | 40.23% (81909) |
| 3+ | 55.94% (11389) | 22.45% (45700) |
| **Cancer** |  |  |
| None | 74.00% (15065) | 94.42% (192220) |
| Solid Tumor | 11.11% (2262) | 4.80% (9764) |
| Metastatic | 14.89% (3032) | 0.79% (1606) |
| **Trauma** | 15.03% (3060) | 2.96% (6030) |
| **Hosp./Surgery** | 30.22% (6152) | 2.29% (4672) |
| **Hypercoag. Cond** | 10.99% (2237) | 1.25% (2551) |
| **Varicose Veins** | 2.73% (556) | 0.88% (1790) |
| **CAD** | 13.47% (2743) | 4.80% (9775) |
| **Stroke** | 7.34% (1495) | 2.40% (4886) |
| **Hyperlipidemia** | 46.18% (9402) | 37.35% (76043) |
| **Smoking** | 23.13% (4710) | 9.46% (19263) |

### Statin and Hormone Exposures

Recent exposures to any HT occurred in 9% (n=19,558) of the sample (10.5% cases,8.6% controls). Of those exposures, 76% were menopausal estrogen or estrogen-progestogen combinations (71% oral/29% non-oral), 13% were estrogen-progestin contraceptives, 8% were progesterone only, and 4% were estrogen-testosterone combinations. Those with recent HT exposure tended to be younger (34% unexposed,43% exposed <55 yrs.). For statin therapy, 16% (n=36,238;18.3% cases,16.0% controls) of the sample had current/continuous exposure for at least three months before and including the index date, 8% (n=17,569) had shorter or discontinuous exposures, and 76% (n=170,142) had no exposure in the past year. Shorter/intermittent exposure was not significantly different from no exposure (OR=1.05,95%CI: 0.99-1.12) but was significantly different from three or more months of current/continuous exposure (OR=1.18,95% CI:1.10-1.25), indicating that the intermittent and no exposure groups could be combined. Those with current/continuous statin exposure tended to be older (29% unexposed, 45% exposed >60), have hyperlipidemia diagnoses (29% unexposed, 86% exposed), or have CAD (4% unexposed, 14% exposed) than those without current/continuous exposure. Among those currently exposed to statins, 80% were exposed to low/moderate intensity and 20% were exposed to high intensity in the 30 days prior to the index date.

After adjustment for all covariates in a conditional model, the OR for any recent HT exposure (regardless of statin exposure) was elevated 51% compared to those without recent HT exposure (**Table 2**, Model 1: OR=1.51,95% CI:1.43-1.60). The OR for current/continuous statin therapy (regardless of HT exposure) was 12% lower than less/no statin therapy (OR=0.88,95%CI: 0.84-0.93). There was slightly less reduction with low/moderate intensity (OR=0.90,95%CI: 0.85-0.95) and slightly more with high-intensity (OR=0.82,95% CI:0.75-0.90) statin therapy compared to those without current statin exposure (not shown). Results for HT and statin therapy were consistent across the total sample with and without a conditional analysis, between the total sample and subsamples without cancer or hypercoagulability conditions, and subsamples with above and below median comorbidity burdens (**Table 2**). The one exception was the higher association of HT exposure in subsamples with fewer comorbidities. There was no significant interaction between HT and statin therapy on VTE risk.

**Table 2.** Sub-Analyses Stratified by Comorbidities.

| **VARIABLE\*** | **N** | **Model 1:**<br>**Full Sample<sup>a</sup>**<br>**(conditional)** | **Model 2:**<br>**Full Sample<sup>b</sup>**<br>**(*unconditional*)** | **Model 3:**<br>**Cancer &**<br>**Hypercoag**<br>**Exclusion<sup>c</sup>**<br>**(*unconditional*)** | **Model 4:**<br>**Stratified**<br>**Elixhauser<2<sup>d</sup>**<br>**(unconditional)** | **Model 5:**<br>**Stratified**<br>**Elixhauser ≥2<sup>d</sup>**<br>**(*unconditional*)** |
| --- | --- | --- | --- | --- | --- | --- |
| **Case Count** | 20359 | 20359 | 20359 | 13835 | 5850 | 14509 |
| **Control Count** | 203590 | 203590 | 203590 | 190170 | 124217 | 79373 |
| **HT None or in 61-365d** | 204391 | 1 | 1 | 1 | 1 | 1 |
| **HT Any Recent (0-60d)** | 19558 | 1.51 (1.43, 1.60) | 1.51 (1.43, 1.59) | 1.56 (1.48, 1.65) | 1.90 (1.76, 2.07) | 1.26 (1.17, 1.35) |
| **Statin <3 mo or None** | 187711 | 1 | 1 | 1 | 1 | 1 |
| **Statin ≥3 mo** | 36238 | 0.88 (0.84, 0.93) | 0.88 (0.84, 0.92) | 0.91 (0.86, 0.96) | 0.90 (0.81, 1.01) | 0.89 (0.85, 0.94) |

a. Conditional model adjusted for age, comorbidity score, region, cancer, hospitalization/surgery, trauma, hypercoagulability, varicose veins, coronary artery disease, stroke, lipid disorders, and smoking history.
b. Unconditional model adjusted for age, comorbidity score, region, cancer, hospitalization/surgery, trauma, hypercoagulability, varicose veins, coronary artery disease, stroke, lipid disorders, smoking history, and date.
c. Unconditional model adjusted for age, comorbidity score, region, hospitalization/surgery, trauma, varicose veins, coronary artery disease, stroke, lipid disorders, smoking history, and date.
d. Unconditional model adjusted for age, region, cancer, hospitalization/surgery, trauma, hypercoagulability, varicose veins, coronary artery disease, stroke, lipid disorders, smoking history, and date.

### Patient Groups with HT and/or Statin Exposures

The combined main effects of statin and HT were estimated for the four groups created by HT and statin exposures. As hypothesized, the OR was highest for women exposed to HT without statin therapy compared to the reference group with neither recent HT nor statin exposure (**Table 3**, Model 1). The OR for those with HT exposure but without statin therapy (n=16,350) was 53% elevated (OR=1.53,95%CI:1.44-1.63) over the reference group (n=171,361). The OR for HT exposure combined with statin therapy (n=3,208) was 25% higher (OR=1.25,95%CI:1.10-1.43) than the reference group. A direct comparison between HT with statin therapy and HT without statin therapy showed a 18% significantly lower OR with statin therapy (OR=0.82,95%CI: 0.71-0.94). Finally, the lowest risk was for those exposed to statin therapy without HT exposure; the OR was 11% (OR=0.89,95%CI: 0.85-0.94) lower than the reference group. (See Appendix, eTable 2 for risk factor distributions for the four exposure groups.)

**Table 3:** VTE Odds Ratios for patient Subgroups by Recent HT and Statin Exposures.

| Exposure | Total N | Cases | Controls | Adj Model 1:<br>ORs<sup>a</sup> | Adj Model 2<br>with Statin Intensity:<br>ORs<sup>a</sup> |
| --- | --- | --- | --- | --- | --- |
| **No recent HT &**<br>**No current Statin (Ref)** | 171361 | 14865 | 156496 | 1 | 1 |
| **Recent HT**<br>**(without Current Statin)** | 16350 | 1777 | 14573 | 1.53 (1.44, 1.63)<sup>b</sup> | 1.53 (1.44, 1.63)<sup>c,d</sup> |
| **Recent HT with**<br>**Current Statin** | 3208 | 353 | 2855 | 1.25 (1.10, 1.43)<sup>b</sup> | -- |
| *Low/Mod Statin* | 2668 | 295 | 2373 | -- | 1.29 (1.12, 1.49)<sup>c</sup> |
| *High Statin* | 540 | 58 | 482 | -- | 1.06 (0.77, 1.45)<sup>d</sup> |
| **Current Statin**<br>**(Without Recent HT)** | 33030 | 3364 | 29666 | 0.89 (0.85, 0.94) | -- |
| *Low/Mod Statin* | 26343 | 2599 | 23744 | -- | 0.91 (0.86, 0.96) |
| *High Statin* | 6687 | 765 | 5922 | -- | 0.84 (0.76, 0.92) |

- a. Odds Ratios have been adjusted for age, comorbidity score, region, cancer, hospitalization/surgery, trauma, hypercoagulability, varicose veins, coronary artery disease, stroke, lipid disorders, and smoking history. - b. A direct comparison between recent HT with and without current statin exposure showed a 18% significantly lower OR with statin therapy (OR=0.82, 95% CI: 0.71-0.94). - c. A direct comparison between HT without statin therapy and HT with low/moderate statin intensity showed a 16% reduction (OR=0.84, 95% CI: 0.73-0.98). - d. A direct comparison between HT without statin therapy and high statin intensity showed a 31% reduction (OR=0.69, 95% CI: 0.50-0.95).

When subgroups exposed to statins were subdivided into low/moderate or high-intensity therapy, a larger protective association was observed with high-intensity therapy compared to the reference group without recent HT or statin exposure (**Table 3**, Model 2). ORs were elevated 53% with recent HT exposure without statin (as above) and were elevated 29% with low/moderate statin intensity (OR=1.29,95%CI:1.12-1.49), but were not elevated with high statin intensity (OR=1.06,95%CI:0.77-1.45) compared to those without recent HT and without current statin therapy. A direct comparison between HT without statin therapy and HT with low/moderate statin intensity showed a 16% reduction (OR=0.84,95% CI:0.73-0.98) and comparison to HT with high statin intensity showed a 31% reduction (OR=0.69,95%CI: 0.50-0.95). The OR for those without recent HT exposure showed a 9% reduction with low/moderate intensity but a 16% reduction with high-intensity statin therapy (OR=0.84,95%CI: 0.76-0.92) compared to the reference group without HT and without statin.

## DISCUSSION

This is the first large study in a US claims database to assess risk of VTE from HT and statin exposures in women of post-menopausal age. In weighing the benefits of menopausal symptom relief against reported risks of VTE from clinical trials^6,10^ and cohort studies,^11,17^ this study provides insight into additional factors affecting the risk profile of HT users. Specifically, we found that the risk-benefit profile for HT may be more favorable when taking concomitant statin therapy. In this study, results indicated HT-associated VTEs may be reduced with statin therapy. Recent HT exposure elevated risk 53% without statin exposure, but elevated risk 25% when combined with statin therapy (compared to the reference group without HT and without statin exposure) – a significant 18% reduction. Higher intensity statin therapy reduced risk 31% (compared to the reference group) and mitigated HT-associated risk, providing evidence of a dose-response effect in the association.

The overall risk reduction for those with recent HT and concomitant statin therapy was similar to previous studies, although observational studies conducted two decades ago were limited by the low prevalence of statin usage. This study and a UK study^27^ each showed an almost 20% reduction in HT risk with statin therapy, although the UK study used an earlier time period (1987-2008), broader age range (50-79), and focused on oral menopausal HT exposures. In contrast to this study, the UK study did not detect significant reductions with higher intensity statins. The difference may be due to different definitions of statin intensity and/or the proportion of women exposed to statins. Similarly, the re-analysis of the HERS cohort, showed VTE risk from HT was 75% greater than placebo (HR=1.75,p=0.04) for those unexposed to statins, but women exposed to both statins and HT had a lower, non-significant 34% elevation in risk compared to placebo (HR=1.34,p=0.45),^28^ suggesting insufficient statistical power.

The overall 51% increase in risk for VTE with HT exposure found in this study is consistent with other studies. A large UK case-control study found any menopausal HT exposure in the past 90 days increased the odds ratio 43%, oral exposures increased the odds ratio 58%, and CEE exposures almost doubled the odds ratio compared to no HT exposure.^17^ Clinical trial estimates indicating HT may double the risk for VTE^9,10^ may be higher because of older age of HT initiation and use of oral CEE.^15,17^ This study therefore affirms and expands previous findings.

In this study, HT risk was slightly higher for those with fewer comorbidities and slightly lower for those with more comorbidities compared to no HT exposure. This difference was likely due to greater exposure to contraceptives (and higher risk estrogen and progestin formulations) in the younger US women with fewer comorbidities.^15^ A notable difference between this sample and the UK samples^17,27^ was the number of women in the US sample on higher-risk combined hormone contraceptives.^15^ Women over 50 years of age exposed to combined hormonal contraceptives had five to nine times higher risk of VTE compared to those with no hormone exposure.^15^

The overall reduction in VTE risk with statin therapy observed in this study (12%) is smaller than estimated in previous observational studies but similar to the reduction found in RCTs. Cohort studies suggest statin exposure may reduce VTE risk by 25%,^25^while RCTs indicate a 15% reduction in risk^25^ and accentuation by intensity of therapy.^25,26,43,44^ Differences between cohort and RCT findings may be attributable to differences in patient populations,^45^ as well as duration of statin exposure. Statin effectiveness studies typically have a longer median statin exposure time (approximately four years)^25^ than in this study, where VTE reduction was observed with 90 or more days. In this study, statin therapy did not interact with HT, but instead had an independent reduction in VTE.

### Limitations

There are several limitations to this study. Most importantly, this study is a nested case-control design in an administrative claims database and contains all inherent limitations of a secondary, observational dataset. Although the study cannot directly prove causality, the time-restricted design offered an unbiased estimation of exposure risk for the population in the base cohort. Misclassification was minimized by employing a strict case definition. While others^17,27,30,46^ included “probable” cases in their analyses, we combined diagnostic claim codes with confirmatory events (i.e., hospitalization, death, or anticoagulant prescription within 30 days of diagnosis) to increase diagnostic accuracy and detection of “definite” cases.^47^ We also adjusted for comorbidities that could potentially confound estimates and controlled for CAD, stroke, and lipid disorders to minimize indication bias. However, even with statistical controls to minimize bias, some residual confounding may remain when treatment groups are not randomly assigned.

Other limitations include a limited lookback period, rather than a lifetime medical history, and limited information on patient characteristics. The database contained data on patient age, sex, and region of residence, but lacked data on race/ethnicity, income, educational level, and indications for prescriptions. Race/ethnicity is modestly associated with VTE,^48,49^ but may be mediated by comorbidities, region, and/or socioeconomic status.^49^ In this study, we controlled for comorbidities and region of residence. Socioeconomic status was indirectly controlled for as all patients had private health insurance. The dataset also did not have information on over-the-counter medications, such as aspirin, which may^50^ or may not^51^ have an independent protective association on VTE in this population.

Despite these limitations, medication and diagnostic detail contained in the large administrative claims database allowed for the estimation of associations for low prevalence exposures and outcomes. This was especially true for estimating risk for those with both hormone and statin exposures. Furthermore, there were sufficient women to make inferences about intensity of statin therapy.

## CONCLUSION

In this case-control study of women with VTE and matched controls, we found statin therapy may reduce, although not eliminate, the risk of VTE associated with exogenous hormones. While women who are at higher cardiovascular risk may take statins, and thus may seem unlikely candidates for HT, statin therapy appears to mitigate some of the risk from HT. HT may not be contraindicated in women who are candidates for statin therapy.

## Data Availability

These data are proprietary and are not available for public viewing. Authors can be contacted with reasonable queries on specific questions they may have related to the data.

## Author Contributions

Davis and Weller each had full access to all the data in the study and take responsibility for the integrity of the data and the accuracy of the data analysis.

*Concept and design:* Davis, Weller, Wilkinson, Porterfield.

*Acquisition, analysis, or interpretation of data:* Chen created the dataset and variables, Davis and Weller analyzed the data, and all authors participated in interpretation of results.

*Drafting of the manuscript:* First draft, Davis, and all authors participated in editing/revising.

*Critical revision of the manuscript for important intellectual content:* All authors.

*Statistical analysis:* Davis, Weller.

*Obtained funding:* Weller.

*Administrative, technical, or material support:* Weller.

*Study supervision:* Weller

## Data Sharing

All data used in these analyses are proprietary (Optum Health Systems) and are not publicly available. Authors can provide variable definitions and statistical code and will consider requests for further analyses.

