## Supplemental Tables for "Statin Use And The Risk Of Venous Thromboembolism In Women Taking Hormone Therapy"

**Online Supplement**

**eTable 1: List of Statin Exposures with NDC and Dose…………………………….p 2**

**eTable 2: Demographics and Comorbidities by Case and Exposure……………p 75**

| eTable 1 List of Statin Exposures NDC \| Name \| Dose | | |
| --- | --- | --- |
| 00026288351 | BAYCOL | 0.2 MG |
| 00026288386 | BAYCOL | 0.2 MG |
| 00026288451 | BAYCOL | 0.3 MG |
| 00026288486 | BAYCOL | 0.3 MG |
| 54569458900 | BAYCOL | 0.3 MG |
| 00026288551 | BAYCOL | 0.4 MG |
| 00026288569 | BAYCOL | 0.4 MG |
| 00026288586 | BAYCOL | 0.4 MG |
| 54569486100 | BAYCOL | 0.4 MG |
| 54868443600 | BAYCOL | 0.4 MG |
| 00026288669 | BAYCOL | 0.8 MG |
| 00026288686 | BAYCOL | 0.8 MG |
| 54569518000 | BAYCOL | 0.8 MG |
| 54868440100 | BAYCOL | 0.8 MG |
| 00002477090 | LIVALO | 1 MG |
| 25208020009 | ZYPITAMAG | 1 MG |
| 66869010490 | LIVALO | 1 MG |
| 00071015523 | LIPITOR | 10 MG |
| 00071015534 | LIPITOR | 10 MG |
| 00071015540 | LIPITOR | 10 MG |
| 00093505698 | ATORVASTATIN CALCIUM | 10 MG |
| 00378201505 | ATORVASTATIN CALCIUM | 10 MG |
| 00378201577 | ATORVASTATIN CALCIUM | 10 MG |
| 00378395005 | ATORVASTATIN CALCIUM | 10 MG |
| 00378395007 | ATORVASTATIN CALCIUM | 10 MG |
| 00378395009 | ATORVASTATIN CALCIUM | 10 MG |
| 00378395077 | ATORVASTATIN CALCIUM | 10 MG |
| 00591377410 | ATORVASTATIN CALCIUM | 10 MG |
| 00591377419 | ATORVASTATIN CALCIUM | 10 MG |
| 00781538192 | ATORVASTATIN CALCIUM | 10 MG |
| 00904629061 | ATORVASTATIN CALCIUM | 10 MG |
| 10135064910 | ATORVASTATIN CALCIUM | 10 MG |
| 13411011301 | LIPITOR | 10 MG |
| 13411011303 | LIPITOR | 10 MG |
| 13411011306 | LIPITOR | 10 MG |
| 13411011309 | LIPITOR | 10 MG |
| 13411011315 | LIPITOR | 10 MG |
| 16714087401 | ATORVASTATIN CALCIUM | 10 MG |
| 16714087402 | ATORVASTATIN CALCIUM | 10 MG |
| 16714087403 | ATORVASTATIN CALCIUM | 10 MG |
| 16729004417 | ATORVASTATIN CALCIUM | 10 MG |
| 33261095900 | ATORVASTATIN CALCIUM | 10 MG |
| 33261095930 | ATORVASTATIN CALCIUM | 10 MG |
| 33261095960 | ATORVASTATIN CALCIUM | 10 MG |
| 33261095990 | ATORVASTATIN CALCIUM | 10 MG |
| 33358021001 | LIPITOR | 10 MG |
| 33358021030 | LIPITOR | 10 MG |
| 33358021060 | LIPITOR | 10 MG |
| 33358021090 | LIPITOR | 10 MG |
| 35356086018 | ATORVASTATIN CALCIUM | 10 MG |
| 35356086030 | ATORVASTATIN CALCIUM | 10 MG |
| 35356086090 | ATORVASTATIN CALCIUM | 10 MG |
| 42254030730 | ATORVASTATIN CALCIUM | 10 MG |
| 42254039130 | ATORVASTATIN CALCIUM | 10 MG |
| 42291014310 | ATORVASTATIN CALCIUM | 10 MG |
| 42291014390 | ATORVASTATIN CALCIUM | 10 MG |
| 43063037330 | ATORVASTATIN CALCIUM | 10 MG |
| 43063047430 | ATORVASTATIN CALCIUM | 10 MG |
| 49999039230 | LIPITOR | 10 MG |
| 49999039290 | LIPITOR | 10 MG |
| 50090125400 | ATORVASTATIN CALCIUM | 10 MG |
| 50090125401 | ATORVASTATIN CALCIUM | 10 MG |
| 50090125500 | ATORVASTATIN CALCIUM | 10 MG |
| 50090125501 | ATORVASTATIN CALCIUM | 10 MG |
| 50268009311 | ATORVASTATIN CALCIUM AVPAK | 10 MG |
| 50268009315 | ATORVASTATIN CALCIUM AVPAK | 10 MG |
| 51079020801 | ATORVASTATIN CALCIUM | 10 MG |
| 51079020820 | ATORVASTATIN CALCIUM | 10 MG |
| 51079040901 | ATORVASTATIN CALCIUM | 10 MG |
| 51079040920 | ATORVASTATIN CALCIUM | 10 MG |
| 51407007810 | ATORVASTATIN CALCIUM | 10 MG |
| 51407007890 | ATORVASTATIN CALCIUM | 10 MG |
| 51655022624 | LIPITOR | 10 MG |
| 51655061030 | ATORVASTATIN CALCIUM | 10 MG |
| 51655061352 | ATORVASTATIN CALCIUM | 10 MG |
| 51655061452 | ATORVASTATIN CALCIUM | 10 MG |
| 52959075990 | LIPITOR | 10 MG |
| 54569446600 | LIPITOR | 10 MG |
| 54569446601 | LIPITOR | 10 MG |
| 54569446602 | LIPITOR | 10 MG |
| 54569628200 | ATORVASTATIN CALCIUM | 10 MG |
| 54569628201 | ATORVASTATIN CALCIUM | 10 MG |
| 54868393400 | LIPITOR | 10 MG |
| 54868393401 | LIPITOR | 10 MG |
| 54868393402 | LIPITOR | 10 MG |
| 54868393403 | LIPITOR | 10 MG |
| 54868393404 | LIPITOR | 10 MG |
| 54868631900 | ATORVASTATIN CALCIUM | 10 MG |
| 55111012105 | ATORVASTATIN CALCIUM | 10 MG |
| 55111012190 | ATORVASTATIN CALCIUM | 10 MG |
| 55175532503 | LIPITOR | 10 MG |
| 55175532509 | LIPITOR | 10 MG |
| 55289087030 | LIPITOR | 10 MG |
| 55700047790 | ATORVASTATIN CALCIUM | 10 MG |
| 55700065930 | ATORVASTATIN CALCIUM | 10 MG |
| 55887062430 | LIPITOR | 10 MG |
| 55887062460 | LIPITOR | 10 MG |
| 55887062482 | LIPITOR | 10 MG |
| 55887062490 | LIPITOR | 10 MG |
| 57866861501 | LIPITOR | 10 MG |
| 58864060830 | LIPITOR | 10 MG |
| 59762015501 | ATORVASTATIN CALCIUM | 10 MG |
| 59762015502 | ATORVASTATIN CALCIUM | 10 MG |
| 60429032301 | ATORVASTATIN CALCIUM | 10 MG |
| 60429032310 | ATORVASTATIN CALCIUM | 10 MG |
| 60429032377 | ATORVASTATIN CALCIUM | 10 MG |
| 60429032390 | ATORVASTATIN CALCIUM | 10 MG |
| 60505257808 | ATORVASTATIN CALCIUM | 10 MG |
| 60505257809 | ATORVASTATIN CALCIUM | 10 MG |
| 60760035330 | ATORVASTATIN CALCIUM | 10 MG |
| 60760035390 | ATORVASTATIN CALCIUM | 10 MG |
| 60760090330 | ATORVASTATIN CALCIUM | 10 MG |
| 60760090390 | ATORVASTATIN CALCIUM | 10 MG |
| 61919054030 | ATORVASTATIN CALCIUM | 10 MG |
| 61919095630 | ATORVASTATIN CALCIUM | 10 MG |
| 61919095690 | ATORVASTATIN CALCIUM | 10 MG |
| 62175089043 | ATORVASTATIN CALCIUM | 10 MG |
| 62175089046 | ATORVASTATIN CALCIUM | 10 MG |
| 63304082705 | ATORVASTATIN CALCIUM | 10 MG |
| 63304082790 | ATORVASTATIN CALCIUM | 10 MG |
| 63629144601 | LIPITOR | 10 MG |
| 63629144602 | LIPITOR | 10 MG |
| 66105011309 | LIPITOR | 10 MG |
| 66116027630 | LIPITOR | 10 MG |
| 67801030103 | LIPITOR | 10 MG |
| 67877051110 | ATORVASTATIN CALCIUM | 10 MG |
| 67877051190 | ATORVASTATIN CALCIUM | 10 MG |
| 68071039930 | LIPITOR | 10 MG |
| 68071091430 | ATORVASTATIN CALCIUM | 10 MG |
| 68084009701 | ATORVASTATIN CALCIUM | 10 MG |
| 68084009711 | ATORVASTATIN CALCIUM | 10 MG |
| 68084056401 | ATORVASTATIN CALCIUM | 10 MG |
| 68115083630 | LIPITOR | 10 MG |
| 68115083690 | LIPITOR | 10 MG |
| 68258600003 | LIPITOR | 10 MG |
| 68258600009 | LIPITOR | 10 MG |
| 68382024910 | ATORVASTATIN CALCIUM | 10 MG |
| 68382024916 | ATORVASTATIN CALCIUM | 10 MG |
| 68645040270 | ATORVASTATIN CALCIUM | 10 MG |
| 68645045854 | ATORVASTATIN CALCIUM | 10 MG |
| 68645045870 | ATORVASTATIN CALCIUM | 10 MG |
| 68645048054 | ATORVASTATIN CALCIUM | 10 MG |
| 68645048070 | ATORVASTATIN CALCIUM | 10 MG |
| 69097089705 | ATORVASTATIN CALCIUM | 10 MG |
| 69097089715 | ATORVASTATIN CALCIUM | 10 MG |
| 69097094405 | ATORVASTATIN CALCIUM | 10 MG |
| 69097094415 | ATORVASTATIN CALCIUM | 10 MG |
| 70377002711 | ATORVASTATIN CALCIUM | 10 MG |
| 70377002713 | ATORVASTATIN CALCIUM | 10 MG |
| 70882010630 | ATORVASTATIN CALCIUM | 10 MG |
| 70882011930 | ATORVASTATIN CALCIUM | 10 MG |
| 70934007030 | ATORVASTATIN CALCIUM | 10 MG |
| 71205024630 | ATORVASTATIN CALCIUM | 10 MG |
| 71205024690 | ATORVASTATIN CALCIUM | 10 MG |
| 71335016901 | ATORVASTATIN CALCIUM | 10 MG |
| 71335016902 | ATORVASTATIN CALCIUM | 10 MG |
| 71335016903 | ATORVASTATIN CALCIUM | 10 MG |
| 71335016904 | ATORVASTATIN CALCIUM | 10 MG |
| 71399051001 | ATORVASTATIN CALCIUM | 10 MG |
| 72205002205 | ATORVASTATIN CALCIUM | 10 MG |
| 72205002290 | ATORVASTATIN CALCIUM | 10 MG |
| 76519107503 | ATORVASTATIN CALCIUM | 10 MG |
| 00006073061 | MEVACOR | 10 MG |
| 00093092606 | LOVASTATIN | 10 MG |
| 00093092610 | LOVASTATIN | 10 MG |
| 00093092619 | LOVASTATIN | 10 MG |
| 00093092693 | LOVASTATIN | 10 MG |
| 00185007001 | LOVASTATIN | 10 MG |
| 00185007005 | LOVASTATIN | 10 MG |
| 00185007010 | LOVASTATIN | 10 MG |
| 00185007060 | LOVASTATIN | 10 MG |
| 00228263306 | LOVASTATIN | 10 MG |
| 00228263350 | LOVASTATIN | 10 MG |
| 00378651091 | LOVASTATIN | 10 MG |
| 00781132305 | LOVASTATIN | 10 MG |
| 00781132360 | LOVASTATIN | 10 MG |
| 00904558152 | LOVASTATIN | 10 MG |
| 10544023590 | LOVASTATIN | 10 MG |
| 21695053430 | LOVASTATIN | 10 MG |
| 23490583802 | LOVASTATIN | 10 MG |
| 23490583806 | LOVASTATIN | 10 MG |
| 23490583809 | LOVASTATIN | 10 MG |
| 33261054702 | LOVASTATIN | 10 MG |
| 33261054730 | LOVASTATIN | 10 MG |
| 33261054760 | LOVASTATIN | 10 MG |
| 33261054790 | LOVASTATIN | 10 MG |
| 33358022330 | LOVASTATIN | 10 MG |
| 42254010630 | LOVASTATIN | 10 MG |
| 42254010690 | LOVASTATIN | 10 MG |
| 42291037590 | LOVASTATIN | 10 MG |
| 43063049330 | LOVASTATIN | 10 MG |
| 43063073190 | LOVASTATIN | 10 MG |
| 45963063301 | LOVASTATIN | 10 MG |
| 45963063304 | LOVASTATIN | 10 MG |
| 49884075401 | LOVASTATIN | 10 MG |
| 49884075402 | LOVASTATIN | 10 MG |
| 49884075410 | LOVASTATIN | 10 MG |
| 49999029330 | LOVASTATIN | 10 MG |
| 49999029360 | LOVASTATIN | 10 MG |
| 49999029390 | LOVASTATIN | 10 MG |
| 50090256300 | LOVASTATIN | 10 MG |
| 50090256301 | LOVASTATIN | 10 MG |
| 50090326800 | LOVASTATIN | 10 MG |
| 50090326801 | LOVASTATIN | 10 MG |
| 50090339601 | LOVASTATIN | 10 MG |
| 50268051011 | LOVASTATIN AVPAK | 10 MG |
| 50268051015 | LOVASTATIN AVPAK | 10 MG |
| 51079097401 | LOVASTATIN | 10 MG |
| 51079097420 | LOVASTATIN | 10 MG |
| 51655001326 | LOVASTATIN | 10 MG |
| 52959097400 | LOVASTATIN | 10 MG |
| 52959097430 | LOVASTATIN | 10 MG |
| 53217030402 | LOVASTATIN | 10 MG |
| 53217030430 | LOVASTATIN | 10 MG |
| 53217030490 | LOVASTATIN | 10 MG |
| 53489060701 | LOVASTATIN | 10 MG |
| 53489060706 | LOVASTATIN | 10 MG |
| 54458084616 | LOVASTATIN | 10 MG |
| 54458091610 | LOVASTATIN | 10 MG |
| 54458093810 | LOVASTATIN | 10 MG |
| 54458093816 | LOVASTATIN | 10 MG |
| 54458098410 | LOVASTATIN | 10 MG |
| 54569458400 | MEVACOR | 10 MG |
| 54569534500 | LOVASTATIN | 10 MG |
| 54569534501 | LOVASTATIN | 10 MG |
| 54868196800 | MEVACOR | 10 MG |
| 54868459300 | LOVASTATIN | 10 MG |
| 54868459301 | LOVASTATIN | 10 MG |
| 54868459302 | LOVASTATIN | 10 MG |
| 55887035030 | LOVASTATIN | 10 MG |
| 57866640001 | LOVASTATIN | 10 MG |
| 58016097900 | LOVASTATIN | 10 MG |
| 58016097902 | LOVASTATIN | 10 MG |
| 58016097920 | LOVASTATIN | 10 MG |
| 58016097930 | LOVASTATIN | 10 MG |
| 58016097960 | LOVASTATIN | 10 MG |
| 58016097990 | LOVASTATIN | 10 MG |
| 58864078130 | LOVASTATIN | 10 MG |
| 60429024810 | LOVASTATIN | 10 MG |
| 60429024860 | LOVASTATIN | 10 MG |
| 60429040010 | LOVASTATIN | 10 MG |
| 60429040060 | LOVASTATIN | 10 MG |
| 60429040090 | LOVASTATIN | 10 MG |
| 60505017700 | LOVASTATIN | 10 MG |
| 60760037130 | LOVASTATIN | 10 MG |
| 61442014101 | LOVASTATIN | 10 MG |
| 61442014110 | LOVASTATIN | 10 MG |
| 61442014160 | LOVASTATIN | 10 MG |
| 61919031190 | LOVASTATIN | 10 MG |
| 62022062730 | ALTOPREV | 10 MG |
| 62037079101 | LOVASTATIN | 10 MG |
| 62037079160 | LOVASTATIN | 10 MG |
| 63187081430 | LOVASTATIN | 10 MG |
| 63187081490 | LOVASTATIN | 10 MG |
| 63629358301 | LOVASTATIN | 10 MG |
| 63629358302 | LOVASTATIN | 10 MG |
| 63629358303 | LOVASTATIN | 10 MG |
| 63739028010 | LOVASTATIN | 10 MG |
| 63739028015 | LOVASTATIN | 10 MG |
| 66336060205 | LOVASTATIN | 10 MG |
| 66336060230 | LOVASTATIN | 10 MG |
| 66336060290 | LOVASTATIN | 10 MG |
| 68001021300 | LOVASTATIN | 10 MG |
| 68001021306 | LOVASTATIN | 10 MG |
| 68001021308 | LOVASTATIN | 10 MG |
| 68001031400 | LOVASTATIN | 10 MG |
| 68001031408 | LOVASTATIN | 10 MG |
| 68084013101 | LOVASTATIN | 10 MG |
| 68084055801 | LOVASTATIN | 10 MG |
| 68084055811 | LOVASTATIN | 10 MG |
| 68115021830 | LOVASTATIN | 10 MG |
| 68180046701 | LOVASTATIN | 10 MG |
| 68180046703 | LOVASTATIN | 10 MG |
| 68180046707 | LOVASTATIN | 10 MG |
| 68645057690 | LOVASTATIN | 10 MG |
| 00003015450 | PRAVACHOL | 10 MG |
| 00003015451 | PRAVACHOL | 10 MG |
| 00003515405 | PRAVACHOL | 10 MG |
| 00003515406 | PRAVACHOL | 10 MG |
| 00093077110 | PRAVASTATIN SODIUM | 10 MG |
| 00093077198 | PRAVASTATIN SODIUM | 10 MG |
| 00378055277 | PRAVASTATIN SODIUM | 10 MG |
| 00378821010 | PRAVASTATIN SODIUM | 10 MG |
| 00378821077 | PRAVASTATIN SODIUM | 10 MG |
| 00591001310 | PRAVASTATIN SODIUM | 10 MG |
| 00591001319 | PRAVASTATIN SODIUM | 10 MG |
| 00781523110 | PRAVASTATIN SODIUM | 10 MG |
| 00781523192 | PRAVASTATIN SODIUM | 10 MG |
| 00904589161 | PRAVASTATIN SODIUM | 10 MG |
| 00904611361 | PRAVASTATIN SODIUM | 10 MG |
| 10544044030 | PRAVASTATIN SODIUM | 10 MG |
| 12280003890 | PRAVACHOL | 10 MG |
| 16252052690 | PRAVASTATIN SODIUM | 10 MG |
| 16729000815 | PRAVASTATIN SODIUM | 10 MG |
| 16729000816 | PRAVASTATIN SODIUM | 10 MG |
| 21695017830 | PRAVASTATIN SODIUM | 10 MG |
| 23490935003 | PRAVASTATIN SODIUM | 10 MG |
| 23490935006 | PRAVASTATIN SODIUM | 10 MG |
| 23490935009 | PRAVASTATIN SODIUM | 10 MG |
| 35356092130 | PRAVASTATIN SODIUM | 10 MG |
| 42254042430 | PRAVASTATIN SODIUM | 10 MG |
| 42291066510 | PRAVASTATIN SODIUM | 10 MG |
| 42291066590 | PRAVASTATIN SODIUM | 10 MG |
| 42549070830 | PRAVASTATIN SODIUM | 10 MG |
| 49884017609 | PRAVASTATIN SODIUM | 10 MG |
| 49884017610 | PRAVASTATIN SODIUM | 10 MG |
| 50090159400 | PRAVASTATIN SODIUM | 10 MG |
| 50090159401 | PRAVASTATIN SODIUM | 10 MG |
| 50090325800 | PRAVASTATIN SODIUM | 10 MG |
| 50090325801 | PRAVASTATIN SODIUM | 10 MG |
| 50111076117 | PRAVASTATIN SODIUM | 10 MG |
| 50268067211 | PRAVASTATIN SODIUM AVPAK | 10 MG |
| 50268067215 | PRAVASTATIN SODIUM AVPAK | 10 MG |
| 53217020330 | PRAVASTATIN SODIUM | 10 MG |
| 53217020390 | PRAVASTATIN SODIUM | 10 MG |
| 54458092710 | PRAVASTATIN SODIUM | 10 MG |
| 54458092712 | PRAVASTATIN SODIUM | 10 MG |
| 54458092716 | PRAVASTATIN SODIUM | 10 MG |
| 54458098709 | PRAVASTATIN SODIUM | 10 MG |
| 54569384000 | PRAVACHOL | 10 MG |
| 54569434600 | PRAVACHOL | 10 MG |
| 54569434601 | PRAVACHOL | 10 MG |
| 54569642800 | PRAVASTATIN SODIUM | 10 MG |
| 54569642801 | PRAVASTATIN SODIUM | 10 MG |
| 54569859800 | PRAVACHOL | 10 MG |
| 54868228701 | PRAVACHOL | 10 MG |
| 54868228702 | PRAVACHOL | 10 MG |
| 54868557600 | PRAVASTATIN SODIUM | 10 MG |
| 54868557601 | PRAVASTATIN SODIUM | 10 MG |
| 55111022905 | PRAVASTATIN SODIUM | 10 MG |
| 55111022990 | PRAVASTATIN SODIUM | 10 MG |
| 55289010430 | PRAVACHOL | 10 MG |
| 57237016405 | PRAVASTATIN SODIUM | 10 MG |
| 57237016490 | PRAVASTATIN SODIUM | 10 MG |
| 58864065330 | PRAVACHOL | 10 MG |
| 60429036705 | PRAVASTATIN SODIUM | 10 MG |
| 60429036745 | PRAVASTATIN SODIUM | 10 MG |
| 60429036790 | PRAVASTATIN SODIUM | 10 MG |
| 60505016805 | PRAVASTATIN SODIUM | 10 MG |
| 60505016809 | PRAVASTATIN SODIUM | 10 MG |
| 60687016901 | PRAVASTATIN SODIUM | 10 MG |
| 60687016911 | PRAVASTATIN SODIUM | 10 MG |
| 63304059590 | PRAVASTATIN SODIUM | 10 MG |
| 63629458801 | PRAVASTATIN SODIUM | 10 MG |
| 66105012001 | PRAVACHOL | 10 MG |
| 66105012003 | PRAVACHOL | 10 MG |
| 66105012006 | PRAVACHOL | 10 MG |
| 66105012009 | PRAVACHOL | 10 MG |
| 66105012015 | PRAVACHOL | 10 MG |
| 66105515405 | PRAVACHOL | 10 MG |
| 68084018601 | PRAVASTATIN SODIUM | 10 MG |
| 68084050001 | PRAVASTATIN SODIUM | 10 MG |
| 68084050011 | PRAVASTATIN SODIUM | 10 MG |
| 68180048502 | PRAVASTATIN SODIUM | 10 MG |
| 68180048509 | PRAVASTATIN SODIUM | 10 MG |
| 68258604903 | PRAVASTATIN SODIUM | 10 MG |
| 68382007005 | PRAVASTATIN SODIUM | 10 MG |
| 68382007016 | PRAVASTATIN SODIUM | 10 MG |
| 68462019505 | PRAVASTATIN SODIUM | 10 MG |
| 68462019590 | PRAVASTATIN SODIUM | 10 MG |
| 68788741303 | PRAVASTATIN SODIUM | 10 MG |
| 68788741306 | PRAVASTATIN SODIUM | 10 MG |
| 68788741309 | PRAVASTATIN SODIUM | 10 MG |
| 71335005601 | PRAVASTATIN SODIUM | 10 MG |
| 00093757198 | ROSUVASTATIN CALCIUM | 10 MG |
| 00310075139 | CRESTOR | 10 MG |
| 00310075190 | CRESTOR | 10 MG |
| 00378220377 | ROSUVASTATIN CALCIUM | 10 MG |
| 00781540192 | ROSUVASTATIN CALCIUM | 10 MG |
| 00904660361 | ROSUVASTATIN CALCIUM | 10 MG |
| 00904677961 | ROSUVASTATIN CALCIUM | 10 MG |
| 12280016415 | CRESTOR | 10 MG |
| 12280016490 | CRESTOR | 10 MG |
| 13668018030 | ROSUVASTATIN CALCIUM | 10 MG |
| 13668018090 | ROSUVASTATIN CALCIUM | 10 MG |
| 16252061630 | ROSUVASTATIN CALCIUM | 10 MG |
| 16252061650 | ROSUVASTATIN CALCIUM | 10 MG |
| 16252061690 | ROSUVASTATIN CALCIUM | 10 MG |
| 16590041130 | CRESTOR | 10 MG |
| 16729028515 | ROSUVASTATIN CALCIUM | 10 MG |
| 16729028517 | ROSUVASTATIN CALCIUM | 10 MG |
| 21695028790 | CRESTOR | 10 MG |
| 27808015601 | ROSUVASTATIN CALCIUM | 10 MG |
| 31722088390 | ROSUVASTATIN CALCIUM | 10 MG |
| 42291074390 | ROSUVASTATIN CALCIUM | 10 MG |
| 42292003001 | ROSUVASTATIN CALCIUM | 10 MG |
| 42292003020 | ROSUVASTATIN CALCIUM | 10 MG |
| 47335058381 | ROSUVASTATIN CALCIUM | 10 MG |
| 47335098583 | EZALLOR SPRINKLE | 10 MG |
| 47463009630 | CRESTOR | 10 MG |
| 49884026109 | ROSUVASTATIN CALCIUM | 10 MG |
| 49999087330 | CRESTOR | 10 MG |
| 49999087390 | CRESTOR | 10 MG |
| 50090245101 | ROSUVASTATIN CALCIUM | 10 MG |
| 50090272300 | ROSUVASTATIN CALCIUM | 10 MG |
| 50090272301 | ROSUVASTATIN CALCIUM | 10 MG |
| 50090317700 | ROSUVASTATIN CALCIUM | 10 MG |
| 50090317701 | ROSUVASTATIN CALCIUM | 10 MG |
| 50268070911 | ROSUVASTATIN CALCIUM AVPAK | 10 MG |
| 50268070915 | ROSUVASTATIN CALCIUM AVPAK | 10 MG |
| 51407015490 | ROSUVASTATIN CALCIUM | 10 MG |
| 53217029530 | ROSUVASTATIN CALCIUM | 10 MG |
| 53217029590 | ROSUVASTATIN CALCIUM | 10 MG |
| 54569560000 | CRESTOR | 10 MG |
| 54569560001 | CRESTOR | 10 MG |
| 54569667400 | ROSUVASTATIN CALCIUM | 10 MG |
| 54569667401 | ROSUVASTATIN CALCIUM | 10 MG |
| 54868496300 | CRESTOR | 10 MG |
| 54868496301 | CRESTOR | 10 MG |
| 54868496302 | CRESTOR | 10 MG |
| 54868496303 | CRESTOR | 10 MG |
| 55048009630 | CRESTOR | 10 MG |
| 55289093530 | CRESTOR | 10 MG |
| 55700051618 | ROSUVASTATIN CALCIUM | 10 MG |
| 55700057430 | ROSUVASTATIN CALCIUM | 10 MG |
| 57237016990 | ROSUVASTATIN CALCIUM | 10 MG |
| 57237016999 | ROSUVASTATIN CALCIUM | 10 MG |
| 58016003700 | CRESTOR | 10 MG |
| 58016003730 | CRESTOR | 10 MG |
| 58016003760 | CRESTOR | 10 MG |
| 58016003790 | CRESTOR | 10 MG |
| 60429084390 | ROSUVASTATIN CALCIUM | 10 MG |
| 60505450309 | ROSUVASTATIN CALCIUM | 10 MG |
| 60687024501 | ROSUVASTATIN CALCIUM | 10 MG |
| 60687024511 | ROSUVASTATIN CALCIUM | 10 MG |
| 63187086430 | ROSUVASTATIN CALCIUM | 10 MG |
| 63187086490 | ROSUVASTATIN CALCIUM | 10 MG |
| 63629338101 | CRESTOR | 10 MG |
| 63629338102 | CRESTOR | 10 MG |
| 63629338103 | CRESTOR | 10 MG |
| 63629338104 | CRESTOR | 10 MG |
| 65862029490 | ROSUVASTATIN CALCIUM | 10 MG |
| 66105098803 | CRESTOR | 10 MG |
| 67877044005 | ROSUVASTATIN CALCIUM | 10 MG |
| 67877044090 | ROSUVASTATIN CALCIUM | 10 MG |
| 68071043330 | CRESTOR | 10 MG |
| 68258601603 | CRESTOR | 10 MG |
| 68462026290 | ROSUVASTATIN CALCIUM | 10 MG |
| 70377000712 | ROSUVASTATIN CALCIUM | 10 MG |
| 70377000713 | ROSUVASTATIN CALCIUM | 10 MG |
| 71205000830 | ROSUVASTATIN CALCIUM | 10 MG |
| 71205005230 | ROSUVASTATIN CALCIUM | 10 MG |
| 71205005290 | ROSUVASTATIN CALCIUM | 10 MG |
| 71335060601 | ROSUVASTATIN CALCIUM | 10 MG |
| 71335060602 | ROSUVASTATIN CALCIUM | 10 MG |
| 72205000390 | ROSUVASTATIN CALCIUM | 10 MG |
| 72205000399 | ROSUVASTATIN CALCIUM | 10 MG |
| 76519116209 | ROSUVASTATIN CALCIUM | 10 MG |
| 00006073528 | ZOCOR | 10 MG |
| 00006073531 | ZOCOR | 10 MG |
| 00006073554 | ZOCOR | 10 MG |
| 00006073561 | ZOCOR | 10 MG |
| 00006073582 | ZOCOR | 10 MG |
| 00006073587 | ZOCOR | 10 MG |
| 00093715310 | SIMVASTATIN | 10 MG |
| 00093715319 | SIMVASTATIN | 10 MG |
| 00093715331 | SIMVASTATIN | 10 MG |
| 00093715356 | SIMVASTATIN | 10 MG |
| 00093715393 | SIMVASTATIN | 10 MG |
| 00093715398 | SIMVASTATIN | 10 MG |
| 00406206603 | SIMVASTATIN | 10 MG |
| 00406206605 | SIMVASTATIN | 10 MG |
| 00406206610 | SIMVASTATIN | 10 MG |
| 00406206660 | SIMVASTATIN | 10 MG |
| 00406206690 | SIMVASTATIN | 10 MG |
| 00781507131 | SIMVASTATIN | 10 MG |
| 00781507192 | SIMVASTATIN | 10 MG |
| 00904580061 | SIMVASTATIN | 10 MG |
| 13411016201 | ZOCOR | 10 MG |
| 13411016203 | ZOCOR | 10 MG |
| 13411016206 | ZOCOR | 10 MG |
| 13411016209 | ZOCOR | 10 MG |
| 13411016215 | ZOCOR | 10 MG |
| 16252050630 | SIMVASTATIN | 10 MG |
| 16252050650 | SIMVASTATIN | 10 MG |
| 16252050690 | SIMVASTATIN | 10 MG |
| 16714068201 | SIMVASTATIN | 10 MG |
| 16714068202 | SIMVASTATIN | 10 MG |
| 16714068203 | SIMVASTATIN | 10 MG |
| 16729000410 | SIMVASTATIN | 10 MG |
| 16729000415 | SIMVASTATIN | 10 MG |
| 16729000417 | SIMVASTATIN | 10 MG |
| 21695073930 | SIMVASTATIN | 10 MG |
| 21695073990 | SIMVASTATIN | 10 MG |
| 23490935303 | SIMVASTATIN | 10 MG |
| 23490935306 | SIMVASTATIN | 10 MG |
| 23490935309 | SIMVASTATIN | 10 MG |
| 24658021110 | SIMVASTATIN | 10 MG |
| 24658021130 | SIMVASTATIN | 10 MG |
| 24658021145 | SIMVASTATIN | 10 MG |
| 24658021190 | SIMVASTATIN | 10 MG |
| 24658030110 | SIMVASTATIN | 10 MG |
| 24658030115 | SIMVASTATIN | 10 MG |
| 24658030130 | SIMVASTATIN | 10 MG |
| 24658030145 | SIMVASTATIN | 10 MG |
| 24658030190 | SIMVASTATIN | 10 MG |
| 31722051110 | SIMVASTATIN | 10 MG |
| 31722051190 | SIMVASTATIN | 10 MG |
| 33261054600 | SIMVASTATIN | 10 MG |
| 33261054602 | SIMVASTATIN | 10 MG |
| 33261054630 | SIMVASTATIN | 10 MG |
| 33261054660 | SIMVASTATIN | 10 MG |
| 33261054690 | SIMVASTATIN | 10 MG |
| 35356060430 | SIMVASTATIN | 10 MG |
| 42254012930 | SIMVASTATIN | 10 MG |
| 42254012990 | SIMVASTATIN | 10 MG |
| 42549071590 | SIMVASTATIN | 10 MG |
| 42571001005 | SIMVASTATIN | 10 MG |
| 42571001090 | SIMVASTATIN | 10 MG |
| 43063016230 | SIMVASTATIN | 10 MG |
| 43063072730 | SIMVASTATIN | 10 MG |
| 43063072790 | SIMVASTATIN | 10 MG |
| 45802009301 | SIMVASTATIN | 10 MG |
| 45802009365 | SIMVASTATIN | 10 MG |
| 45802009375 | SIMVASTATIN | 10 MG |
| 50090127700 | SIMVASTATIN | 10 MG |
| 50090127701 | SIMVASTATIN | 10 MG |
| 50268071311 | SIMVASTATIN AVPAK | 10 MG |
| 50268071315 | SIMVASTATIN AVPAK | 10 MG |
| 50742013710 | SIMVASTATIN | 10 MG |
| 51079045401 | SIMVASTATIN | 10 MG |
| 51079045420 | SIMVASTATIN | 10 MG |
| 51079068601 | SIMVASTATIN | 10 MG |
| 51079068620 | SIMVASTATIN | 10 MG |
| 52343002299 | SIMVASTATIN | 10 MG |
| 52959098830 | SIMVASTATIN | 10 MG |
| 54458090010 | SIMVASTATIN | 10 MG |
| 54458093410 | SIMVASTATIN | 10 MG |
| 54458093416 | SIMVASTATIN | 10 MG |
| 54569418000 | ZOCOR | 10 MG |
| 54569418001 | ZOCOR | 10 MG |
| 54569630200 | SIMVASTATIN | 10 MG |
| 54569630201 | SIMVASTATIN | 10 MG |
| 54868263900 | ZOCOR | 10 MG |
| 54868263901 | ZOCOR | 10 MG |
| 54868562700 | SIMVASTATIN | 10 MG |
| 54868562701 | SIMVASTATIN | 10 MG |
| 55045365508 | SIMVASTATIN | 10 MG |
| 55111019805 | SIMVASTATIN | 10 MG |
| 55111019830 | SIMVASTATIN | 10 MG |
| 55111019890 | SIMVASTATIN | 10 MG |
| 55111073510 | SIMVASTATIN | 10 MG |
| 55111073530 | SIMVASTATIN | 10 MG |
| 55111073590 | SIMVASTATIN | 10 MG |
| 55289033814 | SIMVASTATIN | 10 MG |
| 55289033830 | SIMVASTATIN | 10 MG |
| 55289033890 | SIMVASTATIN | 10 MG |
| 55700022330 | SIMVASTATIN | 10 MG |
| 55700051590 | SIMVASTATIN | 10 MG |
| 55887086130 | SIMVASTATIN | 10 MG |
| 55887086160 | SIMVASTATIN | 10 MG |
| 55887086190 | SIMVASTATIN | 10 MG |
| 57866798601 | ZOCOR | 10 MG |
| 58016000800 | SIMVASTATIN | 10 MG |
| 58016000830 | SIMVASTATIN | 10 MG |
| 58016000860 | SIMVASTATIN | 10 MG |
| 58016000890 | SIMVASTATIN | 10 MG |
| 58016036400 | ZOCOR | 10 MG |
| 58016036430 | ZOCOR | 10 MG |
| 58016036460 | ZOCOR | 10 MG |
| 58016036490 | ZOCOR | 10 MG |
| 60760037930 | SIMVASTATIN | 10 MG |
| 60760037990 | SIMVASTATIN | 10 MG |
| 63304079010 | SIMVASTATIN | 10 MG |
| 63304079030 | SIMVASTATIN | 10 MG |
| 63304079090 | SIMVASTATIN | 10 MG |
| 63739042010 | SIMVASTATIN | 10 MG |
| 63739043610 | SIMVASTATIN | 10 MG |
| 63739057110 | SIMVASTATIN | 10 MG |
| 65862005126 | SIMVASTATIN | 10 MG |
| 65862005130 | SIMVASTATIN | 10 MG |
| 65862005190 | SIMVASTATIN | 10 MG |
| 65862005199 | SIMVASTATIN | 10 MG |
| 66267126001 | SIMVASTATIN | 10 MG |
| 68071071630 | SIMVASTATIN | 10 MG |
| 68071171109 | SIMVASTATIN | 10 MG |
| 68084016201 | SIMVASTATIN | 10 MG |
| 68084051101 | SIMVASTATIN | 10 MG |
| 68084051111 | SIMVASTATIN | 10 MG |
| 68115072030 | ZOCOR | 10 MG |
| 68180047801 | SIMVASTATIN | 10 MG |
| 68180047802 | SIMVASTATIN | 10 MG |
| 68180047803 | SIMVASTATIN | 10 MG |
| 68258600903 | SIMVASTATIN | 10 MG |
| 68258600909 | SIMVASTATIN | 10 MG |
| 68382006605 | SIMVASTATIN | 10 MG |
| 68382006606 | SIMVASTATIN | 10 MG |
| 68382006610 | SIMVASTATIN | 10 MG |
| 68382006614 | SIMVASTATIN | 10 MG |
| 68382006616 | SIMVASTATIN | 10 MG |
| 68382006624 | SIMVASTATIN | 10 MG |
| 70377000212 | SIMVASTATIN | 10 MG |
| 70377000214 | SIMVASTATIN | 10 MG |
| 70377000215 | SIMVASTATIN | 10 MG |
| 00069216030 | CADUET | 10 MG-10 MG |
| 00378451705 | AMLODIPINE BESYLATE-ATORVASTATIN CA | 10 MG-10 MG |
| 00378451793 | AMLODIPINE BESYLATE-ATORVASTATIN CA | 10 MG-10 MG |
| 00378616805 | AMLODIPINE BESYLATE-ATORVASTATIN CA | 10 MG-10 MG |
| 00378616877 | AMLODIPINE BESYLATE-ATORVASTATIN CA | 10 MG-10 MG |
| 00378616893 | AMLODIPINE BESYLATE-ATORVASTATIN CA | 10 MG-10 MG |
| 12280039730 | CADUET | 10 MG-10 MG |
| 43598032130 | AMLODIPINE BESYLATE-ATORVASTATIN CA | 10 MG-10 MG |
| 43598032190 | AMLODIPINE BESYLATE-ATORVASTATIN CA | 10 MG-10 MG |
| 54569588100 | CADUET | 10 MG-10 MG |
| 54868556700 | CADUET | 10 MG-10 MG |
| 59762673001 | AMLODIPINE BESYLATE-ATORVASTATIN CA | 10 MG-10 MG |
| 59762673005 | AMLODIPINE BESYLATE-ATORVASTATIN CA | 10 MG-10 MG |
| 59762673007 | AMLODIPINE BESYLATE-ATORVASTATIN CA | 10 MG-10 MG |
| 63304059030 | AMLODIPINE BESYLATE-ATORVASTATIN CA | 10 MG-10 MG |
| 66582032030 | LIPTRUZET | 10 MG-10 MG |
| 66582032054 | LIPTRUZET | 10 MG-10 MG |
| 00115138503 | EZETIMIBE-SIMVASTATIN | 10 MG-10 MG |
| 00115138508 | EZETIMIBE-SIMVASTATIN | 10 MG-10 MG |
| 00115138510 | EZETIMIBE-SIMVASTATIN | 10 MG-10 MG |
| 12280038630 | VYTORIN | 10 MG-10 MG |
| 43598058310 | EZETIMIBE-SIMVASTATIN | 10 MG-10 MG |
| 43598058330 | EZETIMIBE-SIMVASTATIN | 10 MG-10 MG |
| 43598058390 | EZETIMIBE-SIMVASTATIN | 10 MG-10 MG |
| 43598074210 | EZETIMIBE-SIMVASTATIN | 10 MG-10 MG |
| 43598074230 | EZETIMIBE-SIMVASTATIN | 10 MG-10 MG |
| 43598074290 | EZETIMIBE-SIMVASTATIN | 10 MG-10 MG |
| 45963056508 | EZETIMIBE-SIMVASTATIN | 10 MG-10 MG |
| 45963056530 | EZETIMIBE-SIMVASTATIN | 10 MG-10 MG |
| 51407019010 | EZETIMIBE-SIMVASTATIN | 10 MG-10 MG |
| 51407019030 | EZETIMIBE-SIMVASTATIN | 10 MG-10 MG |
| 51407019090 | EZETIMIBE-SIMVASTATIN | 10 MG-10 MG |
| 54569576800 | VYTORIN | 10 MG-10 MG |
| 54868525000 | VYTORIN | 10 MG-10 MG |
| 60429087910 | EZETIMIBE-SIMVASTATIN | 10 MG-10 MG |
| 60429087930 | EZETIMIBE-SIMVASTATIN | 10 MG-10 MG |
| 60429087990 | EZETIMIBE-SIMVASTATIN | 10 MG-10 MG |
| 62559070030 | EZETIMIBE-SIMVASTATIN | 10 MG-10 MG |
| 62559070090 | EZETIMIBE-SIMVASTATIN | 10 MG-10 MG |
| 66582031128 | VYTORIN | 10 MG-10 MG |
| 66582031131 | VYTORIN | 10 MG-10 MG |
| 66582031154 | VYTORIN | 10 MG-10 MG |
| 66582031182 | VYTORIN | 10 MG-10 MG |
| 67877050730 | EZETIMIBE-SIMVASTATIN | 10 MG-10 MG |
| 67877050790 | EZETIMIBE-SIMVASTATIN | 10 MG-10 MG |
| 69238115503 | EZETIMIBE-SIMVASTATIN | 10 MG-10 MG |
| 69238115509 | EZETIMIBE-SIMVASTATIN | 10 MG-10 MG |
| 00006075331 | JUVISYNC | 10 MG-100 MG |
| 00006075354 | JUVISYNC | 10 MG-100 MG |
| 00006075382 | JUVISYNC | 10 MG-100 MG |
| 00069218030 | CADUET | 10 MG-20 MG |
| 00378451805 | AMLODIPINE BESYLATE-ATORVASTATIN CA | 10 MG-20 MG |
| 00378451893 | AMLODIPINE BESYLATE-ATORVASTATIN CA | 10 MG-20 MG |
| 00378616905 | AMLODIPINE BESYLATE-ATORVASTATIN CA | 10 MG-20 MG |
| 00378616977 | AMLODIPINE BESYLATE-ATORVASTATIN CA | 10 MG-20 MG |
| 00378616993 | AMLODIPINE BESYLATE-ATORVASTATIN CA | 10 MG-20 MG |
| 12280039830 | CADUET | 10 MG-20 MG |
| 43598031830 | AMLODIPINE BESYLATE-ATORVASTATIN CA | 10 MG-20 MG |
| 43598031890 | AMLODIPINE BESYLATE-ATORVASTATIN CA | 10 MG-20 MG |
| 54569595100 | CADUET | 10 MG-20 MG |
| 54868520900 | CADUET | 10 MG-20 MG |
| 54868520901 | CADUET | 10 MG-20 MG |
| 59762673101 | AMLODIPINE BESYLATE-ATORVASTATIN CA | 10 MG-20 MG |
| 59762673105 | AMLODIPINE BESYLATE-ATORVASTATIN CA | 10 MG-20 MG |
| 59762673107 | AMLODIPINE BESYLATE-ATORVASTATIN CA | 10 MG-20 MG |
| 63304059130 | AMLODIPINE BESYLATE-ATORVASTATIN CA | 10 MG-20 MG |
| 00115138603 | EZETIMIBE-SIMVASTATIN | 10 MG-20 MG |
| 00115138608 | EZETIMIBE-SIMVASTATIN | 10 MG-20 MG |
| 00115138610 | EZETIMIBE-SIMVASTATIN | 10 MG-20 MG |
| 12280038530 | VYTORIN | 10 MG-20 MG |
| 12280038590 | VYTORIN | 10 MG-20 MG |
| 21695032530 | VYTORIN | 10 MG-20 MG |
| 43598058410 | EZETIMIBE-SIMVASTATIN | 10 MG-20 MG |
| 43598058430 | EZETIMIBE-SIMVASTATIN | 10 MG-20 MG |
| 43598058490 | EZETIMIBE-SIMVASTATIN | 10 MG-20 MG |
| 43598074410 | EZETIMIBE-SIMVASTATIN | 10 MG-20 MG |
| 43598074430 | EZETIMIBE-SIMVASTATIN | 10 MG-20 MG |
| 43598074490 | EZETIMIBE-SIMVASTATIN | 10 MG-20 MG |
| 45963056608 | EZETIMIBE-SIMVASTATIN | 10 MG-20 MG |
| 45963056630 | EZETIMIBE-SIMVASTATIN | 10 MG-20 MG |
| 49999095730 | VYTORIN | 10 MG-20 MG |
| 51407019110 | EZETIMIBE-SIMVASTATIN | 10 MG-20 MG |
| 51407019130 | EZETIMIBE-SIMVASTATIN | 10 MG-20 MG |
| 51407019190 | EZETIMIBE-SIMVASTATIN | 10 MG-20 MG |
| 54569576600 | VYTORIN | 10 MG-20 MG |
| 54868518700 | VYTORIN | 10 MG-20 MG |
| 54868518701 | VYTORIN | 10 MG-20 MG |
| 54868518702 | VYTORIN | 10 MG-20 MG |
| 55048082130 | VYTORIN | 10 MG-20 MG |
| 55289098021 | VYTORIN | 10 MG-20 MG |
| 55887088230 | VYTORIN | 10 MG-20 MG |
| 60429088010 | EZETIMIBE-SIMVASTATIN | 10 MG-20 MG |
| 60429088030 | EZETIMIBE-SIMVASTATIN | 10 MG-20 MG |
| 60429088090 | EZETIMIBE-SIMVASTATIN | 10 MG-20 MG |
| 62559070130 | EZETIMIBE-SIMVASTATIN | 10 MG-20 MG |
| 62559070190 | EZETIMIBE-SIMVASTATIN | 10 MG-20 MG |
| 66582031228 | VYTORIN | 10 MG-20 MG |
| 66582031231 | VYTORIN | 10 MG-20 MG |
| 66582031254 | VYTORIN | 10 MG-20 MG |
| 66582031282 | VYTORIN | 10 MG-20 MG |
| 66582031287 | VYTORIN | 10 MG-20 MG |
| 67877050830 | EZETIMIBE-SIMVASTATIN | 10 MG-20 MG |
| 67877050890 | EZETIMIBE-SIMVASTATIN | 10 MG-20 MG |
| 68258697003 | VYTORIN | 10 MG-20 MG |
| 69238115603 | EZETIMIBE-SIMVASTATIN | 10 MG-20 MG |
| 69238115609 | EZETIMIBE-SIMVASTATIN | 10 MG-20 MG |
| 00069225030 | CADUET | 10 MG-40 MG |
| 00378451905 | AMLODIPINE BESYLATE-ATORVASTATIN CA | 10 MG-40 MG |
| 00378451993 | AMLODIPINE BESYLATE-ATORVASTATIN CA | 10 MG-40 MG |
| 00378617005 | AMLODIPINE BESYLATE-ATORVASTATIN CA | 10 MG-40 MG |
| 00378617077 | AMLODIPINE BESYLATE-ATORVASTATIN CA | 10 MG-40 MG |
| 00378617093 | AMLODIPINE BESYLATE-ATORVASTATIN CA | 10 MG-40 MG |
| 43598031530 | AMLODIPINE BESYLATE-ATORVASTATIN CA | 10 MG-40 MG |
| 43598031590 | AMLODIPINE BESYLATE-ATORVASTATIN CA | 10 MG-40 MG |
| 54569609900 | CADUET | 10 MG-40 MG |
| 54868520000 | CADUET | 10 MG-40 MG |
| 54868520001 | CADUET | 10 MG-40 MG |
| 59762673201 | AMLODIPINE BESYLATE-ATORVASTATIN CA | 10 MG-40 MG |
| 59762673205 | AMLODIPINE BESYLATE-ATORVASTATIN CA | 10 MG-40 MG |
| 59762673207 | AMLODIPINE BESYLATE-ATORVASTATIN CA | 10 MG-40 MG |
| 63304050030 | AMLODIPINE BESYLATE-ATORVASTATIN CA | 10 MG-40 MG |
| 00115138702 | EZETIMIBE-SIMVASTATIN | 10 MG-40 MG |
| 00115138708 | EZETIMIBE-SIMVASTATIN | 10 MG-40 MG |
| 00115138710 | EZETIMIBE-SIMVASTATIN | 10 MG-40 MG |
| 12280018130 | VYTORIN | 10 MG-40 MG |
| 12280018190 | VYTORIN | 10 MG-40 MG |
| 21695033930 | VYTORIN | 10 MG-40 MG |
| 43598058510 | EZETIMIBE-SIMVASTATIN | 10 MG-40 MG |
| 43598058530 | EZETIMIBE-SIMVASTATIN | 10 MG-40 MG |
| 43598058590 | EZETIMIBE-SIMVASTATIN | 10 MG-40 MG |
| 43598074305 | EZETIMIBE-SIMVASTATIN | 10 MG-40 MG |
| 43598074330 | EZETIMIBE-SIMVASTATIN | 10 MG-40 MG |
| 43598074390 | EZETIMIBE-SIMVASTATIN | 10 MG-40 MG |
| 45963056708 | EZETIMIBE-SIMVASTATIN | 10 MG-40 MG |
| 45963056730 | EZETIMIBE-SIMVASTATIN | 10 MG-40 MG |
| 49999095830 | VYTORIN | 10 MG-40 MG |
| 51407019205 | EZETIMIBE-SIMVASTATIN | 10 MG-40 MG |
| 51407019230 | EZETIMIBE-SIMVASTATIN | 10 MG-40 MG |
| 51407019290 | EZETIMIBE-SIMVASTATIN | 10 MG-40 MG |
| 54569564800 | VYTORIN | 10 MG-40 MG |
| 54868518900 | VYTORIN | 10 MG-40 MG |
| 54868518901 | VYTORIN | 10 MG-40 MG |
| 55048082230 | VYTORIN | 10 MG-40 MG |
| 55289028030 | VYTORIN | 10 MG-40 MG |
| 55887033330 | VYTORIN | 10 MG-40 MG |
| 60429088105 | EZETIMIBE-SIMVASTATIN | 10 MG-40 MG |
| 60429088110 | EZETIMIBE-SIMVASTATIN | 10 MG-40 MG |
| 60429088130 | EZETIMIBE-SIMVASTATIN | 10 MG-40 MG |
| 60429088190 | EZETIMIBE-SIMVASTATIN | 10 MG-40 MG |
| 62559070230 | EZETIMIBE-SIMVASTATIN | 10 MG-40 MG |
| 62559070290 | EZETIMIBE-SIMVASTATIN | 10 MG-40 MG |
| 66582031331 | VYTORIN | 10 MG-40 MG |
| 66582031352 | VYTORIN | 10 MG-40 MG |
| 66582031354 | VYTORIN | 10 MG-40 MG |
| 66582031374 | VYTORIN | 10 MG-40 MG |
| 66582031386 | VYTORIN | 10 MG-40 MG |
| 67877050930 | EZETIMIBE-SIMVASTATIN | 10 MG-40 MG |
| 67877050990 | EZETIMIBE-SIMVASTATIN | 10 MG-40 MG |
| 68258698403 | VYTORIN | 10 MG-40 MG |
| 69238115703 | EZETIMIBE-SIMVASTATIN | 10 MG-40 MG |
| 69238115709 | EZETIMIBE-SIMVASTATIN | 10 MG-40 MG |
| 00006053331 | JUVISYNC | 10 MG-50 MG |
| 00006053354 | JUVISYNC | 10 MG-50 MG |
| 00069227030 | CADUET | 10 MG-80 MG |
| 00378452093 | AMLODIPINE BESYLATE-ATORVASTATIN CA | 10 MG-80 MG |
| 00378617177 | AMLODIPINE BESYLATE-ATORVASTATIN CA | 10 MG-80 MG |
| 00378617193 | AMLODIPINE BESYLATE-ATORVASTATIN CA | 10 MG-80 MG |
| 43598031330 | AMLODIPINE BESYLATE-ATORVASTATIN CA | 10 MG-80 MG |
| 54868552300 | CADUET | 10 MG-80 MG |
| 54868552301 | CADUET | 10 MG-80 MG |
| 54868633500 | AMLODIPINE BESYLATE-ATORVASTATIN CA | 10 MG-80 MG |
| 59762673301 | AMLODIPINE BESYLATE-ATORVASTATIN CA | 10 MG-80 MG |
| 63304060330 | AMLODIPINE BESYLATE-ATORVASTATIN CA | 10 MG-80 MG |
| 00115138802 | EZETIMIBE-SIMVASTATIN | 10 MG-80 MG |
| 00115138808 | EZETIMIBE-SIMVASTATIN | 10 MG-80 MG |
| 00115138810 | EZETIMIBE-SIMVASTATIN | 10 MG-80 MG |
| 21695082730 | VYTORIN | 10 MG-80 MG |
| 43598058610 | EZETIMIBE-SIMVASTATIN | 10 MG-80 MG |
| 43598058630 | EZETIMIBE-SIMVASTATIN | 10 MG-80 MG |
| 43598058690 | EZETIMIBE-SIMVASTATIN | 10 MG-80 MG |
| 43598074530 | EZETIMIBE-SIMVASTATIN | 10 MG-80 MG |
| 43598074590 | EZETIMIBE-SIMVASTATIN | 10 MG-80 MG |
| 45963056808 | EZETIMIBE-SIMVASTATIN | 10 MG-80 MG |
| 45963056830 | EZETIMIBE-SIMVASTATIN | 10 MG-80 MG |
| 51407019305 | EZETIMIBE-SIMVASTATIN | 10 MG-80 MG |
| 51407019330 | EZETIMIBE-SIMVASTATIN | 10 MG-80 MG |
| 51407019390 | EZETIMIBE-SIMVASTATIN | 10 MG-80 MG |
| 54868525900 | VYTORIN | 10 MG-80 MG |
| 54868525901 | VYTORIN | 10 MG-80 MG |
| 55048082330 | VYTORIN | 10 MG-80 MG |
| 55289052030 | VYTORIN | 10 MG-80 MG |
| 60429088205 | EZETIMIBE-SIMVASTATIN | 10 MG-80 MG |
| 60429088230 | EZETIMIBE-SIMVASTATIN | 10 MG-80 MG |
| 60429088290 | EZETIMIBE-SIMVASTATIN | 10 MG-80 MG |
| 62559070330 | EZETIMIBE-SIMVASTATIN | 10 MG-80 MG |
| 62559070390 | EZETIMIBE-SIMVASTATIN | 10 MG-80 MG |
| 66582031531 | VYTORIN | 10 MG-80 MG |
| 66582031552 | VYTORIN | 10 MG-80 MG |
| 66582031554 | VYTORIN | 10 MG-80 MG |
| 66582031566 | VYTORIN | 10 MG-80 MG |
| 66582031574 | VYTORIN | 10 MG-80 MG |
| 67877051030 | EZETIMIBE-SIMVASTATIN | 10 MG-80 MG |
| 67877051090 | EZETIMIBE-SIMVASTATIN | 10 MG-80 MG |
| 69238115803 | EZETIMIBE-SIMVASTATIN | 10 MG-80 MG |
| 69238115809 | EZETIMIBE-SIMVASTATIN | 10 MG-80 MG |
| 00074331690 | SIMCOR | 1000 MG-20 MG |
| 00074345590 | SIMCOR | 1000 MG-20 MG |
| 54868590400 | SIMCOR | 1000 MG-20 MG |
| 54868590401 | SIMCOR | 1000 MG-20 MG |
| 00074345790 | SIMCOR | 1000 MG-40 MG |
| 54868616900 | SIMCOR | 1000 MG-40 MG |
| 00002477190 | LIVALO | 2 MG |
| 25208020109 | ZYPITAMAG | 2 MG |
| 66869020407 | LIVALO | 2 MG |
| 66869020490 | LIVALO | 2 MG |
| 00069296030 | CADUET | 2.5 MG-10 MG |
| 00378451093 | AMLODIPINE BESYLATE-ATORVASTATIN CA | 2.5 MG-10 MG |
| 00378616177 | AMLODIPINE BESYLATE-ATORVASTATIN CA | 2.5 MG-10 MG |
| 00378616193 | AMLODIPINE BESYLATE-ATORVASTATIN CA | 2.5 MG-10 MG |
| 43598032330 | AMLODIPINE BESYLATE-ATORVASTATIN CA | 2.5 MG-10 MG |
| 59762671001 | AMLODIPINE BESYLATE-ATORVASTATIN CA | 2.5 MG-10 MG |
| 63304050130 | AMLODIPINE BESYLATE-ATORVASTATIN CA | 2.5 MG-10 MG |
| 00069297030 | CADUET | 2.5 MG-20 MG |
| 00378451193 | AMLODIPINE BESYLATE-ATORVASTATIN CA | 2.5 MG-20 MG |
| 00378616277 | AMLODIPINE BESYLATE-ATORVASTATIN CA | 2.5 MG-20 MG |
| 00378616293 | AMLODIPINE BESYLATE-ATORVASTATIN CA | 2.5 MG-20 MG |
| 43598032030 | AMLODIPINE BESYLATE-ATORVASTATIN CA | 2.5 MG-20 MG |
| 59762671101 | AMLODIPINE BESYLATE-ATORVASTATIN CA | 2.5 MG-20 MG |
| 63304050230 | AMLODIPINE BESYLATE-ATORVASTATIN CA | 2.5 MG-20 MG |
| 00069298030 | CADUET | 2.5 MG-40 MG |
| 00378451293 | AMLODIPINE BESYLATE-ATORVASTATIN CA | 2.5 MG-40 MG |
| 00378616377 | AMLODIPINE BESYLATE-ATORVASTATIN CA | 2.5 MG-40 MG |
| 00378616393 | AMLODIPINE BESYLATE-ATORVASTATIN CA | 2.5 MG-40 MG |
| 43598031730 | AMLODIPINE BESYLATE-ATORVASTATIN CA | 2.5 MG-40 MG |
| 54868569900 | CADUET | 2.5 MG-40 MG |
| 59762671201 | AMLODIPINE BESYLATE-ATORVASTATIN CA | 2.5 MG-40 MG |
| 63304050330 | AMLODIPINE BESYLATE-ATORVASTATIN CA | 2.5 MG-40 MG |
| 00071015623 | LIPITOR | 20 MG |
| 00071015640 | LIPITOR | 20 MG |
| 00071015694 | LIPITOR | 20 MG |
| 00093505998 | ATORVASTATIN CALCIUM | 20 MG |
| 00378201705 | ATORVASTATIN CALCIUM | 20 MG |
| 00378201777 | ATORVASTATIN CALCIUM | 20 MG |
| 00378395105 | ATORVASTATIN CALCIUM | 20 MG |
| 00378395107 | ATORVASTATIN CALCIUM | 20 MG |
| 00378395109 | ATORVASTATIN CALCIUM | 20 MG |
| 00378395177 | ATORVASTATIN CALCIUM | 20 MG |
| 00591377510 | ATORVASTATIN CALCIUM | 20 MG |
| 00591377519 | ATORVASTATIN CALCIUM | 20 MG |
| 00781538292 | ATORVASTATIN CALCIUM | 20 MG |
| 00904629161 | ATORVASTATIN CALCIUM | 20 MG |
| 10135065005 | ATORVASTATIN CALCIUM | 20 MG |
| 13411011401 | LIPITOR | 20 MG |
| 13411011403 | LIPITOR | 20 MG |
| 13411011406 | LIPITOR | 20 MG |
| 13411011409 | LIPITOR | 20 MG |
| 13411011415 | LIPITOR | 20 MG |
| 16714087501 | ATORVASTATIN CALCIUM | 20 MG |
| 16714087502 | ATORVASTATIN CALCIUM | 20 MG |
| 16714087503 | ATORVASTATIN CALCIUM | 20 MG |
| 16729004517 | ATORVASTATIN CALCIUM | 20 MG |
| 33261097200 | ATORVASTATIN CALCIUM | 20 MG |
| 33261097230 | ATORVASTATIN CALCIUM | 20 MG |
| 33261097260 | ATORVASTATIN CALCIUM | 20 MG |
| 33261097290 | ATORVASTATIN CALCIUM | 20 MG |
| 33261097299 | ATORVASTATIN CALCIUM | 20 MG |
| 35356089418 | ATORVASTATIN CALCIUM | 20 MG |
| 35356089430 | ATORVASTATIN CALCIUM | 20 MG |
| 35356089490 | ATORVASTATIN CALCIUM | 20 MG |
| 42254026130 | ATORVASTATIN CALCIUM | 20 MG |
| 42254026145 | ATORVASTATIN CALCIUM | 20 MG |
| 42254026190 | ATORVASTATIN CALCIUM | 20 MG |
| 42254038230 | ATORVASTATIN CALCIUM | 20 MG |
| 42254038290 | ATORVASTATIN CALCIUM | 20 MG |
| 42291014410 | ATORVASTATIN CALCIUM | 20 MG |
| 42291014490 | ATORVASTATIN CALCIUM | 20 MG |
| 43063049630 | ATORVASTATIN CALCIUM | 20 MG |
| 43063049660 | ATORVASTATIN CALCIUM | 20 MG |
| 49999046730 | LIPITOR | 20 MG |
| 49999046790 | LIPITOR | 20 MG |
| 50090125700 | ATORVASTATIN CALCIUM | 20 MG |
| 50090125701 | ATORVASTATIN CALCIUM | 20 MG |
| 50090125800 | ATORVASTATIN CALCIUM | 20 MG |
| 50090125801 | ATORVASTATIN CALCIUM | 20 MG |
| 50268009411 | ATORVASTATIN CALCIUM AVPAK | 20 MG |
| 50268009415 | ATORVASTATIN CALCIUM AVPAK | 20 MG |
| 50436998803 | ATORVASTATIN CALCIUM | 20 MG |
| 51079020901 | ATORVASTATIN CALCIUM | 20 MG |
| 51079020920 | ATORVASTATIN CALCIUM | 20 MG |
| 51079041001 | ATORVASTATIN CALCIUM | 20 MG |
| 51079041020 | ATORVASTATIN CALCIUM | 20 MG |
| 51407007905 | ATORVASTATIN CALCIUM | 20 MG |
| 51407007990 | ATORVASTATIN CALCIUM | 20 MG |
| 51655092030 | ATORVASTATIN CALCIUM | 20 MG |
| 52959076090 | LIPITOR | 20 MG |
| 53217020930 | ATORVASTATIN CALCIUM | 20 MG |
| 53217020960 | ATORVASTATIN CALCIUM | 20 MG |
| 53217020990 | ATORVASTATIN CALCIUM | 20 MG |
| 53217020999 | ATORVASTATIN CALCIUM | 20 MG |
| 54569446700 | LIPITOR | 20 MG |
| 54569446701 | LIPITOR | 20 MG |
| 54569628300 | ATORVASTATIN CALCIUM | 20 MG |
| 54569628301 | ATORVASTATIN CALCIUM | 20 MG |
| 54868394600 | LIPITOR | 20 MG |
| 54868394601 | LIPITOR | 20 MG |
| 54868394602 | LIPITOR | 20 MG |
| 54868394603 | LIPITOR | 20 MG |
| 54868394604 | LIPITOR | 20 MG |
| 54868632000 | ATORVASTATIN CALCIUM | 20 MG |
| 55111012205 | ATORVASTATIN CALCIUM | 20 MG |
| 55111012290 | ATORVASTATIN CALCIUM | 20 MG |
| 55289080030 | LIPITOR | 20 MG |
| 55700054830 | ATORVASTATIN CALCIUM | 20 MG |
| 55700054890 | ATORVASTATIN CALCIUM | 20 MG |
| 55887073030 | LIPITOR | 20 MG |
| 55887073060 | LIPITOR | 20 MG |
| 55887073090 | LIPITOR | 20 MG |
| 58864068530 | LIPITOR | 20 MG |
| 59762015601 | ATORVASTATIN CALCIUM | 20 MG |
| 59762015602 | ATORVASTATIN CALCIUM | 20 MG |
| 60429032401 | ATORVASTATIN CALCIUM | 20 MG |
| 60429032410 | ATORVASTATIN CALCIUM | 20 MG |
| 60429032477 | ATORVASTATIN CALCIUM | 20 MG |
| 60429032490 | ATORVASTATIN CALCIUM | 20 MG |
| 60505257908 | ATORVASTATIN CALCIUM | 20 MG |
| 60505257909 | ATORVASTATIN CALCIUM | 20 MG |
| 60760035430 | ATORVASTATIN CALCIUM | 20 MG |
| 60760035490 | ATORVASTATIN CALCIUM | 20 MG |
| 60760090430 | ATORVASTATIN CALCIUM | 20 MG |
| 61919025830 | ATORVASTATIN CALCIUM | 20 MG |
| 61919025890 | ATORVASTATIN CALCIUM | 20 MG |
| 61919063230 | ATORVASTATIN CALCIUM | 20 MG |
| 61919091630 | ATORVASTATIN CALCIUM | 20 MG |
| 62175089143 | ATORVASTATIN CALCIUM | 20 MG |
| 62175089146 | ATORVASTATIN CALCIUM | 20 MG |
| 63304082805 | ATORVASTATIN CALCIUM | 20 MG |
| 63304082890 | ATORVASTATIN CALCIUM | 20 MG |
| 63629144701 | LIPITOR | 20 MG |
| 63629484901 | ATORVASTATIN CALCIUM | 20 MG |
| 63629484902 | ATORVASTATIN CALCIUM | 20 MG |
| 63629484903 | ATORVASTATIN CALCIUM | 20 MG |
| 66105011409 | LIPITOR | 20 MG |
| 67801040230 | LIPITOR | 20 MG |
| 67877051210 | ATORVASTATIN CALCIUM | 20 MG |
| 67877051290 | ATORVASTATIN CALCIUM | 20 MG |
| 68071015430 | LIPITOR | 20 MG |
| 68071091530 | ATORVASTATIN CALCIUM | 20 MG |
| 68084009801 | ATORVASTATIN CALCIUM | 20 MG |
| 68084009811 | ATORVASTATIN CALCIUM | 20 MG |
| 68084056501 | ATORVASTATIN CALCIUM | 20 MG |
| 68115049430 | LIPITOR | 20 MG |
| 68115049460 | LIPITOR | 20 MG |
| 68115080090 | LIPITOR | 20 MG |
| 68258600103 | LIPITOR | 20 MG |
| 68258600109 | LIPITOR | 20 MG |
| 68382025010 | ATORVASTATIN CALCIUM | 20 MG |
| 68382025016 | ATORVASTATIN CALCIUM | 20 MG |
| 68645040370 | ATORVASTATIN CALCIUM | 20 MG |
| 68645045954 | ATORVASTATIN CALCIUM | 20 MG |
| 68645045970 | ATORVASTATIN CALCIUM | 20 MG |
| 68645048154 | ATORVASTATIN CALCIUM | 20 MG |
| 68645048170 | ATORVASTATIN CALCIUM | 20 MG |
| 68645048254 | ATORVASTATIN CALCIUM | 20 MG |
| 69097089805 | ATORVASTATIN CALCIUM | 20 MG |
| 69097089812 | ATORVASTATIN CALCIUM | 20 MG |
| 69097094505 | ATORVASTATIN CALCIUM | 20 MG |
| 69097094512 | ATORVASTATIN CALCIUM | 20 MG |
| 70377002811 | ATORVASTATIN CALCIUM | 20 MG |
| 70377002813 | ATORVASTATIN CALCIUM | 20 MG |
| 70882010230 | ATORVASTATIN CALCIUM | 20 MG |
| 70882012030 | ATORVASTATIN CALCIUM | 20 MG |
| 70934006930 | ATORVASTATIN CALCIUM | 20 MG |
| 71205024730 | ATORVASTATIN CALCIUM | 20 MG |
| 71205024790 | ATORVASTATIN CALCIUM | 20 MG |
| 71335000401 | ATORVASTATIN CALCIUM | 20 MG |
| 71335000402 | ATORVASTATIN CALCIUM | 20 MG |
| 71335000403 | ATORVASTATIN CALCIUM | 20 MG |
| 71335000404 | ATORVASTATIN CALCIUM | 20 MG |
| 71335000405 | ATORVASTATIN CALCIUM | 20 MG |
| 71399052005 | ATORVASTATIN CALCIUM | 20 MG |
| 72205002305 | ATORVASTATIN CALCIUM | 20 MG |
| 72205002390 | ATORVASTATIN CALCIUM | 20 MG |
| 76519107809 | ATORVASTATIN CALCIUM | 20 MG |
| 00078017605 | LESCOL | 20 MG |
| 00078017615 | LESCOL | 20 MG |
| 00093744201 | FLUVASTATIN | 20 MG |
| 00093744256 | FLUVASTATIN | 20 MG |
| 00378802077 | FLUVASTATIN | 20 MG |
| 00378802093 | FLUVASTATIN | 20 MG |
| 13411011101 | LESCOL | 20 MG |
| 13411011102 | LESCOL | 20 MG |
| 13411011103 | LESCOL | 20 MG |
| 13411011106 | LESCOL | 20 MG |
| 13411011110 | LESCOL | 20 MG |
| 54569382100 | LESCOL | 20 MG |
| 54569382101 | LESCOL | 20 MG |
| 54868332900 | LESCOL | 20 MG |
| 55175300203 | LESCOL | 20 MG |
| 55289074060 | LESCOL | 20 MG |
| 66105014701 | LESCOL | 20 MG |
| 66105014703 | LESCOL | 20 MG |
| 66105014706 | LESCOL | 20 MG |
| 66105014709 | LESCOL | 20 MG |
| 66105014710 | LESCOL | 20 MG |
| 00006073128 | MEVACOR | 20 MG |
| 00006073137 | MEVACOR | 20 MG |
| 00006073161 | MEVACOR | 20 MG |
| 00006073178 | MEVACOR | 20 MG |
| 00006073182 | MEVACOR | 20 MG |
| 00006073187 | MEVACOR | 20 MG |
| 00006073194 | MEVACOR | 20 MG |
| 00006073198 | MEVACOR | 20 MG |
| 00093057606 | LOVASTATIN | 20 MG |
| 00093057610 | LOVASTATIN | 20 MG |
| 00093057619 | LOVASTATIN | 20 MG |
| 00093057693 | LOVASTATIN | 20 MG |
| 00185007201 | LOVASTATIN | 20 MG |
| 00185007210 | LOVASTATIN | 20 MG |
| 00185007260 | LOVASTATIN | 20 MG |
| 00228263406 | LOVASTATIN | 20 MG |
| 00228263450 | LOVASTATIN | 20 MG |
| 00378652005 | LOVASTATIN | 20 MG |
| 00378652091 | LOVASTATIN | 20 MG |
| 00781121010 | LOVASTATIN | 20 MG |
| 00781121060 | LOVASTATIN | 20 MG |
| 00904558252 | LOVASTATIN | 20 MG |
| 10544024130 | LOVASTATIN | 20 MG |
| 10544024630 | LOVASTATIN | 20 MG |
| 12280010860 | LOVASTATIN | 20 MG |
| 16590054730 | LOVASTATIN | 20 MG |
| 16590054760 | LOVASTATIN | 20 MG |
| 16590054772 | LOVASTATIN | 20 MG |
| 16590054790 | LOVASTATIN | 20 MG |
| 21695053530 | LOVASTATIN | 20 MG |
| 21695053590 | LOVASTATIN | 20 MG |
| 23490583900 | LOVASTATIN | 20 MG |
| 23490583901 | LOVASTATIN | 20 MG |
| 33261054802 | LOVASTATIN | 20 MG |
| 33261054830 | LOVASTATIN | 20 MG |
| 33261054860 | LOVASTATIN | 20 MG |
| 33261054890 | LOVASTATIN | 20 MG |
| 33358022500 | LOVASTATIN | 20 MG |
| 33358022530 | LOVASTATIN | 20 MG |
| 33358022560 | LOVASTATIN | 20 MG |
| 35356088530 | LOVASTATIN | 20 MG |
| 35356088560 | LOVASTATIN | 20 MG |
| 35356088590 | LOVASTATIN | 20 MG |
| 42254002830 | LOVASTATIN | 20 MG |
| 42254002890 | LOVASTATIN | 20 MG |
| 42291037690 | LOVASTATIN | 20 MG |
| 42554002830 | LOVASTATIN | 20 MG |
| 43063069290 | LOVASTATIN | 20 MG |
| 43063069293 | LOVASTATIN | 20 MG |
| 43063098330 | LOVASTATIN | 20 MG |
| 45963063401 | LOVASTATIN | 20 MG |
| 45963063404 | LOVASTATIN | 20 MG |
| 49884075501 | LOVASTATIN | 20 MG |
| 49884075502 | LOVASTATIN | 20 MG |
| 49884075510 | LOVASTATIN | 20 MG |
| 49999047030 | LOVASTATIN | 20 MG |
| 49999047060 | LOVASTATIN | 20 MG |
| 49999047090 | LOVASTATIN | 20 MG |
| 50090075900 | LOVASTATIN | 20 MG |
| 50090075902 | LOVASTATIN | 20 MG |
| 50268051111 | LOVASTATIN AVPAK | 20 MG |
| 50268051115 | LOVASTATIN AVPAK | 20 MG |
| 51079097501 | LOVASTATIN | 20 MG |
| 51079097520 | LOVASTATIN | 20 MG |
| 51079097530 | LOVASTATIN | 20 MG |
| 51079097556 | LOVASTATIN | 20 MG |
| 52959072030 | LOVASTATIN | 20 MG |
| 52959072060 | LOVASTATIN | 20 MG |
| 52959072090 | LOVASTATIN | 20 MG |
| 53489060801 | LOVASTATIN | 20 MG |
| 53489060806 | LOVASTATIN | 20 MG |
| 53489060810 | LOVASTATIN | 20 MG |
| 54458084516 | LOVASTATIN | 20 MG |
| 54458087110 | LOVASTATIN | 20 MG |
| 54458091510 | LOVASTATIN | 20 MG |
| 54458093710 | LOVASTATIN | 20 MG |
| 54458093716 | LOVASTATIN | 20 MG |
| 54458098310 | LOVASTATIN | 20 MG |
| 54569061300 | MEVACOR | 20 MG |
| 54569061301 | MEVACOR | 20 MG |
| 54569061302 | MEVACOR | 20 MG |
| 54569061303 | MEVACOR | 20 MG |
| 54569061304 | MEVACOR | 20 MG |
| 54569534600 | LOVASTATIN | 20 MG |
| 54569534602 | LOVASTATIN | 20 MG |
| 54569801100 | MEVACOR | 20 MG |
| 54569886600 | LOVASTATIN | 20 MG |
| 54868068601 | MEVACOR | 20 MG |
| 54868068602 | MEVACOR | 20 MG |
| 54868068603 | MEVACOR | 20 MG |
| 54868068604 | MEVACOR | 20 MG |
| 54868458500 | LOVASTATIN | 20 MG |
| 54868458501 | LOVASTATIN | 20 MG |
| 54868458502 | LOVASTATIN | 20 MG |
| 54868458503 | LOVASTATIN | 20 MG |
| 55045301401 | LOVASTATIN | 20 MG |
| 55045301402 | LOVASTATIN | 20 MG |
| 55045301406 | LOVASTATIN | 20 MG |
| 55045301408 | LOVASTATIN | 20 MG |
| 55045301409 | LOVASTATIN | 20 MG |
| 55048039530 | LOVASTATIN | 20 MG |
| 55175504606 | MEVACOR | 20 MG |
| 55289040030 | MEVACOR | 20 MG |
| 55289088130 | LOVASTATIN | 20 MG |
| 55289088190 | LOVASTATIN | 20 MG |
| 55700024430 | LOVASTATIN | 20 MG |
| 55700024460 | LOVASTATIN | 20 MG |
| 55700024490 | LOVASTATIN | 20 MG |
| 55887097430 | LOVASTATIN | 20 MG |
| 57866660101 | LOVASTATIN | 20 MG |
| 58016090000 | LOVASTATIN | 20 MG |
| 58016090002 | LOVASTATIN | 20 MG |
| 58016090030 | LOVASTATIN | 20 MG |
| 58016090060 | LOVASTATIN | 20 MG |
| 58016090090 | LOVASTATIN | 20 MG |
| 58864078030 | LOVASTATIN | 20 MG |
| 58864078060 | LOVASTATIN | 20 MG |
| 59630062830 | ALTOPREV | 20 MG |
| 60429024910 | LOVASTATIN | 20 MG |
| 60429024960 | LOVASTATIN | 20 MG |
| 60429040110 | LOVASTATIN | 20 MG |
| 60429040160 | LOVASTATIN | 20 MG |
| 60429040190 | LOVASTATIN | 20 MG |
| 60505017800 | LOVASTATIN | 20 MG |
| 60760037030 | LOVASTATIN | 20 MG |
| 61442014201 | LOVASTATIN | 20 MG |
| 61442014205 | LOVASTATIN | 20 MG |
| 61442014210 | LOVASTATIN | 20 MG |
| 61442014260 | LOVASTATIN | 20 MG |
| 61919054730 | LOVASTATIN | 20 MG |
| 61919054790 | LOVASTATIN | 20 MG |
| 61919067671 | LOVASTATIN | 20 MG |
| 62022062830 | ALTOPREV | 20 MG |
| 62022077030 | ALTOCOR | 20 MG |
| 62037079201 | LOVASTATIN | 20 MG |
| 62037079260 | LOVASTATIN | 20 MG |
| 63629146401 | LOVASTATIN | 20 MG |
| 63629146402 | LOVASTATIN | 20 MG |
| 63629146403 | LOVASTATIN | 20 MG |
| 63739028103 | LOVASTATIN | 20 MG |
| 63739028110 | LOVASTATIN | 20 MG |
| 63739028115 | LOVASTATIN | 20 MG |
| 63874036301 | LOVASTATIN | 20 MG |
| 63874036310 | LOVASTATIN | 20 MG |
| 63874036320 | LOVASTATIN | 20 MG |
| 63874036330 | LOVASTATIN | 20 MG |
| 63874036360 | LOVASTATIN | 20 MG |
| 63874036390 | LOVASTATIN | 20 MG |
| 66116027730 | LOVASTATIN | 20 MG |
| 66336031005 | LOVASTATIN | 20 MG |
| 66336031030 | LOVASTATIN | 20 MG |
| 66336031060 | LOVASTATIN | 20 MG |
| 66336031090 | LOVASTATIN | 20 MG |
| 67046045030 | LOVASTATIN | 20 MG |
| 68001022400 | LOVASTATIN | 20 MG |
| 68001022406 | LOVASTATIN | 20 MG |
| 68001022408 | LOVASTATIN | 20 MG |
| 68001031500 | LOVASTATIN | 20 MG |
| 68001031508 | LOVASTATIN | 20 MG |
| 68084013201 | LOVASTATIN | 20 MG |
| 68084055901 | LOVASTATIN | 20 MG |
| 68084055911 | LOVASTATIN | 20 MG |
| 68115021930 | LOVASTATIN | 20 MG |
| 68115021960 | LOVASTATIN | 20 MG |
| 68180046801 | LOVASTATIN | 20 MG |
| 68180046803 | LOVASTATIN | 20 MG |
| 68180046805 | LOVASTATIN | 20 MG |
| 68180046807 | LOVASTATIN | 20 MG |
| 68645056690 | LOVASTATIN | 20 MG |
| 70515062830 | ALTOPREV | 20 MG |
| 00003017850 | PRAVACHOL | 20 MG |
| 00003017851 | PRAVACHOL | 20 MG |
| 00003517805 | PRAVACHOL | 20 MG |
| 00003517806 | PRAVACHOL | 20 MG |
| 00003517875 | PRAVACHOL | 20 MG |
| 00093720110 | PRAVASTATIN SODIUM | 20 MG |
| 00093720198 | PRAVASTATIN SODIUM | 20 MG |
| 00378055477 | PRAVASTATIN SODIUM | 20 MG |
| 00378822010 | PRAVASTATIN SODIUM | 20 MG |
| 00378822077 | PRAVASTATIN SODIUM | 20 MG |
| 00591001410 | PRAVASTATIN SODIUM | 20 MG |
| 00591001419 | PRAVASTATIN SODIUM | 20 MG |
| 00781523210 | PRAVASTATIN SODIUM | 20 MG |
| 00781523292 | PRAVASTATIN SODIUM | 20 MG |
| 00904589261 | PRAVASTATIN SODIUM | 20 MG |
| 00904611461 | PRAVASTATIN SODIUM | 20 MG |
| 10544050530 | PRAVASTATIN SODIUM | 20 MG |
| 13411011801 | PRAVACHOL | 20 MG |
| 13411011802 | PRAVACHOL | 20 MG |
| 13411011803 | PRAVACHOL | 20 MG |
| 13411011806 | PRAVACHOL | 20 MG |
| 13411011809 | PRAVACHOL | 20 MG |
| 16252052750 | PRAVASTATIN SODIUM | 20 MG |
| 16252052790 | PRAVASTATIN SODIUM | 20 MG |
| 16729000915 | PRAVASTATIN SODIUM | 20 MG |
| 16729000916 | PRAVASTATIN SODIUM | 20 MG |
| 16729000917 | PRAVASTATIN SODIUM | 20 MG |
| 21695017930 | PRAVASTATIN SODIUM | 20 MG |
| 21695017990 | PRAVASTATIN SODIUM | 20 MG |
| 23490935103 | PRAVASTATIN SODIUM | 20 MG |
| 23490935106 | PRAVASTATIN SODIUM | 20 MG |
| 23490935109 | PRAVASTATIN SODIUM | 20 MG |
| 33261086700 | PRAVASTATIN SODIUM | 20 MG |
| 33261086730 | PRAVASTATIN SODIUM | 20 MG |
| 33261086760 | PRAVASTATIN SODIUM | 20 MG |
| 33261086790 | PRAVASTATIN SODIUM | 20 MG |
| 35356091930 | PRAVASTATIN SODIUM | 20 MG |
| 35356091990 | PRAVASTATIN SODIUM | 20 MG |
| 42254020230 | PRAVASTATIN SODIUM | 20 MG |
| 42254020290 | PRAVASTATIN SODIUM | 20 MG |
| 42254042530 | PRAVASTATIN SODIUM | 20 MG |
| 42254042590 | PRAVASTATIN SODIUM | 20 MG |
| 42291066710 | PRAVASTATIN SODIUM | 20 MG |
| 42291066790 | PRAVASTATIN SODIUM | 20 MG |
| 43063014330 | PRAVASTATIN SODIUM | 20 MG |
| 43063044330 | PRAVASTATIN SODIUM | 20 MG |
| 43063080730 | PRAVASTATIN SODIUM | 20 MG |
| 49884017909 | PRAVASTATIN SODIUM | 20 MG |
| 49884017910 | PRAVASTATIN SODIUM | 20 MG |
| 50090202500 | PRAVASTATIN SODIUM | 20 MG |
| 50090202501 | PRAVASTATIN SODIUM | 20 MG |
| 50090254500 | PRAVASTATIN SODIUM | 20 MG |
| 50090254501 | PRAVASTATIN SODIUM | 20 MG |
| 50111076203 | PRAVASTATIN SODIUM | 20 MG |
| 50111076217 | PRAVASTATIN SODIUM | 20 MG |
| 50268067311 | PRAVASTATIN SODIUM AVPAK | 20 MG |
| 50268067315 | PRAVASTATIN SODIUM AVPAK | 20 MG |
| 51079045801 | PRAVASTATIN SODIUM | 20 MG |
| 51079045820 | PRAVASTATIN SODIUM | 20 MG |
| 51655007152 | PRAVASTATIN SODIUM | 20 MG |
| 52959099030 | PRAVASTATIN SODIUM | 20 MG |
| 52959099090 | PRAVASTATIN SODIUM | 20 MG |
| 54458086910 | PRAVASTATIN SODIUM | 20 MG |
| 54458090802 | PRAVASTATIN SODIUM | 20 MG |
| 54458092610 | PRAVASTATIN SODIUM | 20 MG |
| 54458092616 | PRAVASTATIN SODIUM | 20 MG |
| 54458098610 | PRAVASTATIN SODIUM | 20 MG |
| 54569371500 | PRAVACHOL | 20 MG |
| 54569371501 | PRAVACHOL | 20 MG |
| 54569371502 | PRAVACHOL | 20 MG |
| 54569371503 | PRAVACHOL | 20 MG |
| 54569407100 | PRAVACHOL | 20 MG |
| 54569579300 | PRAVASTATIN SODIUM | 20 MG |
| 54569579301 | PRAVASTATIN SODIUM | 20 MG |
| 54569851000 | PRAVACHOL | 20 MG |
| 54569851001 | PRAVACHOL | 20 MG |
| 54868228800 | PRAVACHOL | 20 MG |
| 54868228801 | PRAVACHOL | 20 MG |
| 54868228802 | PRAVACHOL | 20 MG |
| 54868557700 | PRAVASTATIN SODIUM | 20 MG |
| 54868557701 | PRAVASTATIN SODIUM | 20 MG |
| 55048059730 | PRAVASTATIN SODIUM | 20 MG |
| 55111023005 | PRAVASTATIN SODIUM | 20 MG |
| 55111023090 | PRAVASTATIN SODIUM | 20 MG |
| 55175539003 | PRAVACHOL | 20 MG |
| 55289087130 | PRAVACHOL | 20 MG |
| 55887020330 | PRAVASTATIN | 20 MG |
| 55887020390 | PRAVASTATIN | 20 MG |
| 57237016505 | PRAVASTATIN SODIUM | 20 MG |
| 57237016590 | PRAVASTATIN SODIUM | 20 MG |
| 58016001300 | PRAVASTATIN | 20 MG |
| 58016001330 | PRAVASTATIN | 20 MG |
| 58016001360 | PRAVASTATIN | 20 MG |
| 58016001390 | PRAVASTATIN | 20 MG |
| 58016042500 | PRAVACHOL | 20 MG |
| 58016042530 | PRAVACHOL | 20 MG |
| 58016042560 | PRAVACHOL | 20 MG |
| 58016042590 | PRAVACHOL | 20 MG |
| 60429036805 | PRAVASTATIN SODIUM | 20 MG |
| 60429036845 | PRAVASTATIN SODIUM | 20 MG |
| 60429036890 | PRAVASTATIN SODIUM | 20 MG |
| 60505016907 | PRAVASTATIN SODIUM | 20 MG |
| 60505016909 | PRAVASTATIN SODIUM | 20 MG |
| 60687017801 | PRAVASTATIN SODIUM | 20 MG |
| 60687017811 | PRAVASTATIN SODIUM | 20 MG |
| 60760007730 | PRAVASTATIN SODIUM | 20 MG |
| 60760007790 | PRAVASTATIN SODIUM | 20 MG |
| 60760042190 | PRAVASTATIN SODIUM | 20 MG |
| 61919073190 | PRAVASTATIN SODIUM | 20 MG |
| 63304059690 | PRAVASTATIN SODIUM | 20 MG |
| 63629356301 | PRAVASTATIN SODIUM | 20 MG |
| 63739064910 | PRAVASTATIN SODIUM | 20 MG |
| 63739064941 | PRAVASTATIN SODIUM | 20 MG |
| 66105012101 | PRAVACHOL | 20 MG |
| 66105012103 | PRAVACHOL | 20 MG |
| 66105012106 | PRAVACHOL | 20 MG |
| 66105012109 | PRAVACHOL | 20 MG |
| 66105012115 | PRAVACHOL | 20 MG |
| 66116023830 | PRAVACHOL | 20 MG |
| 66336068530 | PRAVASTATIN SODIUM | 20 MG |
| 66336068590 | PRAVASTATIN SODIUM | 20 MG |
| 68084018701 | PRAVASTATIN SODIUM | 20 MG |
| 68084050101 | PRAVASTATIN SODIUM | 20 MG |
| 68084050111 | PRAVASTATIN SODIUM | 20 MG |
| 68180048602 | PRAVASTATIN SODIUM | 20 MG |
| 68180048609 | PRAVASTATIN SODIUM | 20 MG |
| 68382007105 | PRAVASTATIN SODIUM | 20 MG |
| 68382007116 | PRAVASTATIN SODIUM | 20 MG |
| 68462019605 | PRAVASTATIN SODIUM | 20 MG |
| 68462019690 | PRAVASTATIN SODIUM | 20 MG |
| 71205014930 | PRAVASTATIN SODIUM | 20 MG |
| 00093757298 | ROSUVASTATIN CALCIUM | 20 MG |
| 00310075239 | CRESTOR | 20 MG |
| 00310075290 | CRESTOR | 20 MG |
| 00378220477 | ROSUVASTATIN CALCIUM | 20 MG |
| 00781540292 | ROSUVASTATIN CALCIUM | 20 MG |
| 00904660461 | ROSUVASTATIN CALCIUM | 20 MG |
| 00904678061 | ROSUVASTATIN CALCIUM | 20 MG |
| 12280035130 | CRESTOR | 20 MG |
| 12280035190 | CRESTOR | 20 MG |
| 13668018130 | ROSUVASTATIN CALCIUM | 20 MG |
| 13668018190 | ROSUVASTATIN CALCIUM | 20 MG |
| 16252061730 | ROSUVASTATIN CALCIUM | 20 MG |
| 16252061750 | ROSUVASTATIN CALCIUM | 20 MG |
| 16252061790 | ROSUVASTATIN CALCIUM | 20 MG |
| 16729028615 | ROSUVASTATIN CALCIUM | 20 MG |
| 16729028617 | ROSUVASTATIN CALCIUM | 20 MG |
| 21695028890 | CRESTOR | 20 MG |
| 27808015701 | ROSUVASTATIN CALCIUM | 20 MG |
| 31722088490 | ROSUVASTATIN CALCIUM | 20 MG |
| 42291074490 | ROSUVASTATIN CALCIUM | 20 MG |
| 42292003101 | ROSUVASTATIN CALCIUM | 20 MG |
| 42292003120 | ROSUVASTATIN CALCIUM | 20 MG |
| 47335058481 | ROSUVASTATIN CALCIUM | 20 MG |
| 47335098683 | EZALLOR SPRINKLE | 20 MG |
| 49884026209 | ROSUVASTATIN CALCIUM | 20 MG |
| 49999099230 | CRESTOR | 20 MG |
| 49999099290 | CRESTOR | 20 MG |
| 50090272400 | ROSUVASTATIN CALCIUM | 20 MG |
| 50090272401 | ROSUVASTATIN CALCIUM | 20 MG |
| 50268071011 | ROSUVASTATIN CALCIUM AVPAK | 20 MG |
| 50268071015 | ROSUVASTATIN CALCIUM AVPAK | 20 MG |
| 51407015590 | ROSUVASTATIN CALCIUM | 20 MG |
| 53217011330 | CRESTOR | 20 MG |
| 53217011390 | CRESTOR | 20 MG |
| 53217029630 | ROSUVASTATIN CALCIUM | 20 MG |
| 53217029690 | ROSUVASTATIN CALCIUM | 20 MG |
| 54569567200 | CRESTOR | 20 MG |
| 54569567202 | CRESTOR | 20 MG |
| 54569667500 | ROSUVASTATIN CALCIUM | 20 MG |
| 54569667501 | ROSUVASTATIN CALCIUM | 20 MG |
| 54868508500 | CRESTOR | 20 MG |
| 54868508501 | CRESTOR | 20 MG |
| 54868508502 | CRESTOR | 20 MG |
| 54868508503 | CRESTOR | 20 MG |
| 54868508504 | CRESTOR | 20 MG |
| 55048009730 | CRESTOR | 20 MG |
| 55289093230 | CRESTOR | 20 MG |
| 55700057530 | ROSUVASTATIN CALCIUM | 20 MG |
| 57237017090 | ROSUVASTATIN CALCIUM | 20 MG |
| 57237017099 | ROSUVASTATIN CALCIUM | 20 MG |
| 58016005200 | CRESTOR | 20 MG |
| 58016005230 | CRESTOR | 20 MG |
| 58016005260 | CRESTOR | 20 MG |
| 58016005290 | CRESTOR | 20 MG |
| 60429084490 | ROSUVASTATIN CALCIUM | 20 MG |
| 60505450409 | ROSUVASTATIN CALCIUM | 20 MG |
| 60687025601 | ROSUVASTATIN CALCIUM | 20 MG |
| 60687025611 | ROSUVASTATIN CALCIUM | 20 MG |
| 63187086530 | ROSUVASTATIN CALCIUM | 20 MG |
| 63187086590 | ROSUVASTATIN CALCIUM | 20 MG |
| 65862029590 | ROSUVASTATIN CALCIUM | 20 MG |
| 66336067430 | CRESTOR | 20 MG |
| 67877044105 | ROSUVASTATIN CALCIUM | 20 MG |
| 67877044190 | ROSUVASTATIN CALCIUM | 20 MG |
| 68071026330 | CRESTOR | 20 MG |
| 68462026390 | ROSUVASTATIN CALCIUM | 20 MG |
| 70377000812 | ROSUVASTATIN CALCIUM | 20 MG |
| 70377000813 | ROSUVASTATIN CALCIUM | 20 MG |
| 71205004490 | ROSUVASTATIN CALCIUM | 20 MG |
| 71205007730 | ROSUVASTATIN CALCIUM | 20 MG |
| 71205009990 | ROSUVASTATIN CALCIUM | 20 MG |
| 71205027930 | ROSUVASTATIN CALCIUM | 20 MG |
| 71335030201 | ROSUVASTATIN CALCIUM | 20 MG |
| 71335030202 | ROSUVASTATIN CALCIUM | 20 MG |
| 71335030203 | ROSUVASTATIN CALCIUM | 20 MG |
| 72205000490 | ROSUVASTATIN CALCIUM | 20 MG |
| 72205000499 | ROSUVASTATIN CALCIUM | 20 MG |
| 76519114903 | ROSUVASTATIN CALCIUM | 20 MG |
| 00006074028 | ZOCOR | 20 MG |
| 00006074031 | ZOCOR | 20 MG |
| 00006074054 | ZOCOR | 20 MG |
| 00006074061 | ZOCOR | 20 MG |
| 00006074082 | ZOCOR | 20 MG |
| 00006074087 | ZOCOR | 20 MG |
| 00093715410 | SIMVASTATIN | 20 MG |
| 00093715419 | SIMVASTATIN | 20 MG |
| 00093715431 | SIMVASTATIN | 20 MG |
| 00093715456 | SIMVASTATIN | 20 MG |
| 00093715493 | SIMVASTATIN | 20 MG |
| 00093715498 | SIMVASTATIN | 20 MG |
| 00406206703 | SIMVASTATIN | 20 MG |
| 00406206705 | SIMVASTATIN | 20 MG |
| 00406206710 | SIMVASTATIN | 20 MG |
| 00406206760 | SIMVASTATIN | 20 MG |
| 00406206790 | SIMVASTATIN | 20 MG |
| 00781507231 | SIMVASTATIN | 20 MG |
| 00781507292 | SIMVASTATIN | 20 MG |
| 00904580161 | SIMVASTATIN | 20 MG |
| 10544048630 | SIMVASTATIN | 20 MG |
| 13411013201 | ZOCOR | 20 MG |
| 13411013203 | ZOCOR | 20 MG |
| 13411013206 | ZOCOR | 20 MG |
| 13411013209 | ZOCOR | 20 MG |
| 13411013215 | ZOCOR | 20 MG |
| 16252050730 | SIMVASTATIN | 20 MG |
| 16252050750 | SIMVASTATIN | 20 MG |
| 16252050790 | SIMVASTATIN | 20 MG |
| 16590044630 | SIMVASTATIN | 20 MG |
| 16714068301 | SIMVASTATIN | 20 MG |
| 16714068302 | SIMVASTATIN | 20 MG |
| 16714068303 | SIMVASTATIN | 20 MG |
| 16729000510 | SIMVASTATIN | 20 MG |
| 16729000515 | SIMVASTATIN | 20 MG |
| 16729000517 | SIMVASTATIN | 20 MG |
| 21695074030 | SIMVASTATIN | 20 MG |
| 21695074090 | SIMVASTATIN | 20 MG |
| 23490935403 | SIMVASTATIN | 20 MG |
| 23490935406 | SIMVASTATIN | 20 MG |
| 23490935409 | SIMVASTATIN | 20 MG |
| 24658021210 | SIMVASTATIN | 20 MG |
| 24658021230 | SIMVASTATIN | 20 MG |
| 24658021245 | SIMVASTATIN | 20 MG |
| 24658021290 | SIMVASTATIN | 20 MG |
| 24658030210 | SIMVASTATIN | 20 MG |
| 24658030215 | SIMVASTATIN | 20 MG |
| 24658030230 | SIMVASTATIN | 20 MG |
| 24658030245 | SIMVASTATIN | 20 MG |
| 24658030290 | SIMVASTATIN | 20 MG |
| 31722051210 | SIMVASTATIN | 20 MG |
| 31722051290 | SIMVASTATIN | 20 MG |
| 33261054102 | SIMVASTATIN | 20 MG |
| 33261054130 | SIMVASTATIN | 20 MG |
| 33261054160 | SIMVASTATIN | 20 MG |
| 33261054190 | SIMVASTATIN | 20 MG |
| 35356066730 | SIMVASTATIN | 20 MG |
| 35356078118 | SIMVASTATIN | 20 MG |
| 35356078130 | SIMVASTATIN | 20 MG |
| 35356078190 | SIMVASTATIN | 20 MG |
| 42254006030 | SIMVASTATIN | 20 MG |
| 42254006090 | SIMVASTATIN | 20 MG |
| 42571002010 | SIMVASTATIN | 20 MG |
| 42571002090 | SIMVASTATIN | 20 MG |
| 43063000801 | SIMVASTATIN | 20 MG |
| 43063000830 | SIMVASTATIN | 20 MG |
| 43063000890 | SIMVASTATIN | 20 MG |
| 45802038401 | SIMVASTATIN | 20 MG |
| 45802038465 | SIMVASTATIN | 20 MG |
| 45802038475 | SIMVASTATIN | 20 MG |
| 45802038493 | SIMVASTATIN | 20 MG |
| 45865042130 | SIMVASTATIN | 20 MG |
| 45865042151 | SIMVASTATIN | 20 MG |
| 45865042160 | SIMVASTATIN | 20 MG |
| 45865042190 | SIMVASTATIN | 20 MG |
| 49999030630 | ZOCOR | 20 MG |
| 49999088930 | SIMVASTATIN | 20 MG |
| 49999088960 | SIMVASTATIN | 20 MG |
| 49999088990 | SIMVASTATIN | 20 MG |
| 50090099901 | SIMVASTATIN | 20 MG |
| 50090099902 | SIMVASTATIN | 20 MG |
| 50090099903 | SIMVASTATIN | 20 MG |
| 50268071411 | SIMVASTATIN AVPAK | 20 MG |
| 50268071415 | SIMVASTATIN AVPAK | 20 MG |
| 50436012202 | SIMVASTATIN | 20 MG |
| 50742013810 | SIMVASTATIN | 20 MG |
| 51079039301 | SIMVASTATIN | 20 MG |
| 51079039320 | SIMVASTATIN | 20 MG |
| 51079045501 | SIMVASTATIN | 20 MG |
| 51079045520 | SIMVASTATIN | 20 MG |
| 52343002390 | SIMVASTATIN | 20 MG |
| 52343002399 | SIMVASTATIN | 20 MG |
| 52959098930 | SIMVASTATIN | 20 MG |
| 52959098990 | SIMVASTATIN | 20 MG |
| 54458089910 | SIMVASTATIN | 20 MG |
| 54458092804 | SIMVASTATIN | 20 MG |
| 54458093310 | SIMVASTATIN | 20 MG |
| 54458093316 | SIMVASTATIN | 20 MG |
| 54569440300 | ZOCOR | 20 MG |
| 54569583300 | SIMVASTATIN | 20 MG |
| 54569583301 | SIMVASTATIN | 20 MG |
| 54569583302 | SIMVASTATIN | 20 MG |
| 54569583303 | SIMVASTATIN | 20 MG |
| 54868310400 | ZOCOR | 20 MG |
| 54868310401 | ZOCOR | 20 MG |
| 54868562800 | SIMVASTATIN | 20 MG |
| 54868562801 | SIMVASTATIN | 20 MG |
| 54868562802 | SIMVASTATIN | 20 MG |
| 55048077530 | SIMVASTATIN | 20 MG |
| 55048077590 | SIMVASTATIN | 20 MG |
| 55111019905 | SIMVASTATIN | 20 MG |
| 55111019910 | SIMVASTATIN | 20 MG |
| 55111019930 | SIMVASTATIN | 20 MG |
| 55111019990 | SIMVASTATIN | 20 MG |
| 55111074010 | SIMVASTATIN | 20 MG |
| 55111074030 | SIMVASTATIN | 20 MG |
| 55111074090 | SIMVASTATIN | 20 MG |
| 55289029314 | SIMVASTATIN | 20 MG |
| 55289029330 | SIMVASTATIN | 20 MG |
| 55289029390 | SIMVASTATIN | 20 MG |
| 55700002130 | SIMVASTATIN | 20 MG |
| 55700002190 | SIMVASTATIN | 20 MG |
| 55700017830 | SIMVASTATIN | 20 MG |
| 55700054990 | SIMVASTATIN | 20 MG |
| 55887032730 | SIMVASTATIN | 20 MG |
| 55887032760 | SIMVASTATIN | 20 MG |
| 55887032790 | SIMVASTATIN | 20 MG |
| 57866393601 | SIMVASTATIN | 20 MG |
| 57866798201 | ZOCOR | 20 MG |
| 58016000700 | SIMVASTATIN | 20 MG |
| 58016000730 | SIMVASTATIN | 20 MG |
| 58016000760 | SIMVASTATIN | 20 MG |
| 58016000790 | SIMVASTATIN | 20 MG |
| 58016038500 | ZOCOR | 20 MG |
| 58016038530 | ZOCOR | 20 MG |
| 58016038560 | ZOCOR | 20 MG |
| 58016038590 | ZOCOR | 20 MG |
| 58864076030 | ZOCOR | 20 MG |
| 60760000530 | SIMVASTATIN | 20 MG |
| 60760000590 | SIMVASTATIN | 20 MG |
| 61919044630 | SIMVASTATIN | 20 MG |
| 61919044660 | SIMVASTATIN | 20 MG |
| 61919044690 | SIMVASTATIN | 20 MG |
| 63304079110 | SIMVASTATIN | 20 MG |
| 63304079130 | SIMVASTATIN | 20 MG |
| 63304079190 | SIMVASTATIN | 20 MG |
| 63629339301 | SIMVASTATIN | 20 MG |
| 63629339302 | SIMVASTATIN | 20 MG |
| 63629339303 | SIMVASTATIN | 20 MG |
| 63629339304 | SIMVASTATIN | 20 MG |
| 63739042110 | SIMVASTATIN | 20 MG |
| 63739043704 | SIMVASTATIN | 20 MG |
| 63739043710 | SIMVASTATIN | 20 MG |
| 63739057210 | SIMVASTATIN | 20 MG |
| 65862005226 | SIMVASTATIN | 20 MG |
| 65862005230 | SIMVASTATIN | 20 MG |
| 65862005290 | SIMVASTATIN | 20 MG |
| 65862005299 | SIMVASTATIN | 20 MG |
| 66105050503 | ZOCOR | 20 MG |
| 66267126101 | SIMVASTATIN | 20 MG |
| 66336095430 | SIMVASTATIN | 20 MG |
| 66336095490 | SIMVASTATIN | 20 MG |
| 68071069930 | SIMVASTATIN | 20 MG |
| 68084016301 | SIMVASTATIN | 20 MG |
| 68084051201 | SIMVASTATIN | 20 MG |
| 68084051211 | SIMVASTATIN | 20 MG |
| 68115067230 | ZOCOR | 20 MG |
| 68180047901 | SIMVASTATIN | 20 MG |
| 68180047902 | SIMVASTATIN | 20 MG |
| 68180047903 | SIMVASTATIN | 20 MG |
| 68382006705 | SIMVASTATIN | 20 MG |
| 68382006706 | SIMVASTATIN | 20 MG |
| 68382006710 | SIMVASTATIN | 20 MG |
| 68382006714 | SIMVASTATIN | 20 MG |
| 68382006716 | SIMVASTATIN | 20 MG |
| 68382006724 | SIMVASTATIN | 20 MG |
| 68645026154 | SIMVASTATIN | 20 MG |
| 68645047054 | SIMVASTATIN | 20 MG |
| 70377000312 | SIMVASTATIN | 20 MG |
| 70377000314 | SIMVASTATIN | 20 MG |
| 70377000315 | SIMVASTATIN | 20 MG |
| 29273040104 | FLOLIPID | 20 MG/5 ML |
| 66582032130 | LIPTRUZET | 20 MG-10 MG |
| 66582032154 | LIPTRUZET | 20 MG-10 MG |
| 00006075731 | JUVISYNC | 20 MG-100 MG |
| 00006075754 | JUVISYNC | 20 MG-100 MG |
| 00006075782 | JUVISYNC | 20 MG-100 MG |
| 00074300790 | ADVICOR | 20 MG-1000 MG |
| 54868508700 | ADVICOR | 20 MG-1000 MG |
| 60598000890 | ADVICOR | 20 MG-1000 MG |
| 00006053531 | JUVISYNC | 20 MG-50 MG |
| 00006053554 | JUVISYNC | 20 MG-50 MG |
| 00074300590 | ADVICOR | 20 MG-500 MG |
| 54868480700 | ADVICOR | 20 MG-500 MG |
| 54868480701 | ADVICOR | 20 MG-500 MG |
| 60598000690 | ADVICOR | 20 MG-500 MG |
| 00074307290 | ADVICOR | 20 MG-750 MG |
| 54868480702 | ADVICOR | 20 MG-750 MG |
| 54868499900 | ADVICOR | 20 MG-750 MG |
| 54868499901 | ADVICOR | 20 MG-750 MG |
| 60598000790 | ADVICOR | 20 MG-750 MG |
| 00003516911 | PRAVIGARD PAC | 325 MG; 20 MG |
| 00003517411 | PRAVIGARD PAC | 325 MG; 40 MG |
| 00003518411 | PRAVIGARD PAC | 325 MG; 80 MG |
| 00002477290 | LIVALO | 4 MG |
| 25208020209 | ZYPITAMAG | 4 MG |
| 66869040407 | LIVALO | 4 MG |
| 66869040490 | LIVALO | 4 MG |
| 00071015723 | LIPITOR | 40 MG |
| 00071015740 | LIPITOR | 40 MG |
| 00071015773 | LIPITOR | 40 MG |
| 00071015788 | LIPITOR | 40 MG |
| 00093505898 | ATORVASTATIN CALCIUM | 40 MG |
| 00378212105 | ATORVASTATIN CALCIUM | 40 MG |
| 00378212177 | ATORVASTATIN CALCIUM | 40 MG |
| 00378395205 | ATORVASTATIN CALCIUM | 40 MG |
| 00378395207 | ATORVASTATIN CALCIUM | 40 MG |
| 00378395209 | ATORVASTATIN CALCIUM | 40 MG |
| 00378395277 | ATORVASTATIN CALCIUM | 40 MG |
| 00591377605 | ATORVASTATIN CALCIUM | 40 MG |
| 00591377619 | ATORVASTATIN CALCIUM | 40 MG |
| 00781538492 | ATORVASTATIN CALCIUM | 40 MG |
| 00904629261 | ATORVASTATIN CALCIUM | 40 MG |
| 10135065110 | ATORVASTATIN CALCIUM | 40 MG |
| 13411011501 | LIPITOR | 40 MG |
| 13411011503 | LIPITOR | 40 MG |
| 13411011506 | LIPITOR | 40 MG |
| 13411011509 | LIPITOR | 40 MG |
| 13411011515 | LIPITOR | 40 MG |
| 16714087601 | ATORVASTATIN CALCIUM | 40 MG |
| 16714087602 | ATORVASTATIN CALCIUM | 40 MG |
| 16714087603 | ATORVASTATIN CALCIUM | 40 MG |
| 16729004617 | ATORVASTATIN CALCIUM | 40 MG |
| 21695025590 | LIPITOR | 40 MG |
| 33261097300 | ATORVASTATIN CALCIUM | 40 MG |
| 33261097330 | ATORVASTATIN CALCIUM | 40 MG |
| 33261097360 | ATORVASTATIN CALCIUM | 40 MG |
| 33261097390 | ATORVASTATIN CALCIUM | 40 MG |
| 35356092930 | ATORVASTATIN CALCIUM | 40 MG |
| 42254001930 | ATORVASTATIN CALCIUM | 40 MG |
| 42254001945 | ATORVASTATIN CALCIUM | 40 MG |
| 42254001990 | ATORVASTATIN CALCIUM | 40 MG |
| 42254037930 | ATORVASTATIN CALCIUM | 40 MG |
| 42254037990 | ATORVASTATIN CALCIUM | 40 MG |
| 42291014550 | ATORVASTATIN CALCIUM | 40 MG |
| 42291014590 | ATORVASTATIN CALCIUM | 40 MG |
| 43063045330 | ATORVASTATIN CALCIUM | 40 MG |
| 49999046830 | LIPITOR | 40 MG |
| 49999046890 | LIPITOR | 40 MG |
| 50090126000 | ATORVASTATIN CALCIUM | 40 MG |
| 50090126001 | ATORVASTATIN CALCIUM | 40 MG |
| 50090126100 | ATORVASTATIN CALCIUM | 40 MG |
| 50090126101 | ATORVASTATIN CALCIUM | 40 MG |
| 50090343800 | ATORVASTATIN CALCIUM | 40 MG |
| 50090343801 | ATORVASTATIN CALCIUM | 40 MG |
| 50268009511 | ATORVASTATIN CALCIUM AVPAK | 40 MG |
| 50268009515 | ATORVASTATIN CALCIUM AVPAK | 40 MG |
| 50436998901 | ATORVASTATIN CALCIUM | 40 MG |
| 51079021001 | ATORVASTATIN CALCIUM | 40 MG |
| 51079021020 | ATORVASTATIN CALCIUM | 40 MG |
| 51079041101 | ATORVASTATIN CALCIUM | 40 MG |
| 51079041120 | ATORVASTATIN CALCIUM | 40 MG |
| 51407008010 | ATORVASTATIN CALCIUM | 40 MG |
| 51407008090 | ATORVASTATIN CALCIUM | 40 MG |
| 51655064030 | ATORVASTATIN CALCIUM | 40 MG |
| 51655065152 | ATORVASTATIN CALCIUM | 40 MG |
| 52959004630 | LIPITOR | 40 MG |
| 54458088110 | ATORVASTATIN CALCIUM | 40 MG |
| 54458088116 | ATORVASTATIN CALCIUM | 40 MG |
| 54569458700 | LIPITOR | 40 MG |
| 54569458701 | LIPITOR | 40 MG |
| 54569628400 | ATORVASTATIN CALCIUM | 40 MG |
| 54569628401 | ATORVASTATIN CALCIUM | 40 MG |
| 54868422900 | LIPITOR | 40 MG |
| 54868422901 | LIPITOR | 40 MG |
| 54868422902 | LIPITOR | 40 MG |
| 54868422903 | LIPITOR | 40 MG |
| 54868632100 | ATORVASTATIN CALCIUM | 40 MG |
| 55111012305 | ATORVASTATIN CALCIUM | 40 MG |
| 55111012390 | ATORVASTATIN CALCIUM | 40 MG |
| 55289086130 | LIPITOR | 40 MG |
| 55887092990 | LIPITOR | 40 MG |
| 58864062315 | LIPITOR | 40 MG |
| 58864062330 | LIPITOR | 40 MG |
| 59762015701 | ATORVASTATIN CALCIUM | 40 MG |
| 59762015702 | ATORVASTATIN CALCIUM | 40 MG |
| 60429032501 | ATORVASTATIN CALCIUM | 40 MG |
| 60429032505 | ATORVASTATIN CALCIUM | 40 MG |
| 60429032577 | ATORVASTATIN CALCIUM | 40 MG |
| 60429032590 | ATORVASTATIN CALCIUM | 40 MG |
| 60505258008 | ATORVASTATIN CALCIUM | 40 MG |
| 60505258009 | ATORVASTATIN CALCIUM | 40 MG |
| 60760035530 | ATORVASTATIN CALCIUM | 40 MG |
| 60760035590 | ATORVASTATIN CALCIUM | 40 MG |
| 60760070930 | ATORVASTATIN CALCIUM | 40 MG |
| 60760090530 | ATORVASTATIN CALCIUM | 40 MG |
| 60760090590 | ATORVASTATIN CALCIUM | 40 MG |
| 61919030330 | ATORVASTATIN CALCIUM | 40 MG |
| 61919030390 | ATORVASTATIN CALCIUM | 40 MG |
| 62175089241 | ATORVASTATIN CALCIUM | 40 MG |
| 62175089246 | ATORVASTATIN CALCIUM | 40 MG |
| 63304082905 | ATORVASTATIN CALCIUM | 40 MG |
| 63304082990 | ATORVASTATIN CALCIUM | 40 MG |
| 63629486501 | ATORVASTATIN CALCIUM | 40 MG |
| 66105011509 | LIPITOR | 40 MG |
| 67801031403 | LIPITOR | 40 MG |
| 67877051310 | ATORVASTATIN CALCIUM | 40 MG |
| 67877051390 | ATORVASTATIN CALCIUM | 40 MG |
| 68071031030 | LIPITOR | 40 MG |
| 68071091630 | ATORVASTATIN CALCIUM | 40 MG |
| 68084009901 | ATORVASTATIN CALCIUM | 40 MG |
| 68084009911 | ATORVASTATIN CALCIUM | 40 MG |
| 68084058901 | ATORVASTATIN CALCIUM | 40 MG |
| 68115066815 | LIPITOR | 40 MG |
| 68115066830 | LIPITOR | 40 MG |
| 68115066890 | LIPITOR | 40 MG |
| 68258600203 | LIPITOR | 40 MG |
| 68258600209 | LIPITOR | 40 MG |
| 68382025110 | ATORVASTATIN CALCIUM | 40 MG |
| 68382025116 | ATORVASTATIN CALCIUM | 40 MG |
| 68645041754 | ATORVASTATIN CALCIUM | 40 MG |
| 68645046054 | ATORVASTATIN CALCIUM | 40 MG |
| 68645048354 | ATORVASTATIN CALCIUM | 40 MG |
| 68645056854 | ATORVASTATIN CALCIUM | 40 MG |
| 69097089905 | ATORVASTATIN CALCIUM | 40 MG |
| 69097089915 | ATORVASTATIN CALCIUM | 40 MG |
| 69097094605 | ATORVASTATIN CALCIUM | 40 MG |
| 69097094615 | ATORVASTATIN CALCIUM | 40 MG |
| 70377002911 | ATORVASTATIN CALCIUM | 40 MG |
| 70377002913 | ATORVASTATIN CALCIUM | 40 MG |
| 70882010930 | ATORVASTATIN CALCIUM | 40 MG |
| 70934009130 | ATORVASTATIN CALCIUM | 40 MG |
| 71205026490 | ATORVASTATIN CALCIUM | 40 MG |
| 71335010201 | ATORVASTATIN CALCIUM | 40 MG |
| 71335010202 | ATORVASTATIN CALCIUM | 40 MG |
| 71335010203 | ATORVASTATIN CALCIUM | 40 MG |
| 71335010204 | ATORVASTATIN CALCIUM | 40 MG |
| 71399054001 | ATORVASTATIN CALCIUM | 40 MG |
| 72205002405 | ATORVASTATIN CALCIUM | 40 MG |
| 72205002490 | ATORVASTATIN CALCIUM | 40 MG |
| 76519106303 | ATORVASTATIN CALCIUM | 40 MG |
| 00078023405 | LESCOL | 40 MG |
| 00078023415 | LESCOL | 40 MG |
| 00093744301 | FLUVASTATIN | 40 MG |
| 00093744356 | FLUVASTATIN | 40 MG |
| 00378802177 | FLUVASTATIN | 40 MG |
| 00378802193 | FLUVASTATIN | 40 MG |
| 54569476100 | LESCOL | 40 MG |
| 54569476101 | LESCOL | 40 MG |
| 54868422400 | LESCOL | 40 MG |
| 54868422401 | LESCOL | 40 MG |
| 55289047630 | LESCOL | 40 MG |
| 00006073261 | MEVACOR | 40 MG |
| 00006073282 | MEVACOR | 40 MG |
| 00006073287 | MEVACOR | 40 MG |
| 00006073294 | MEVACOR | 40 MG |
| 00093092806 | LOVASTATIN | 40 MG |
| 00093092810 | LOVASTATIN | 40 MG |
| 00093092819 | LOVASTATIN | 40 MG |
| 00093092893 | LOVASTATIN | 40 MG |
| 00185007401 | LOVASTATIN | 40 MG |
| 00185007410 | LOVASTATIN | 40 MG |
| 00185007460 | LOVASTATIN | 40 MG |
| 00228263506 | LOVASTATIN | 40 MG |
| 00228263550 | LOVASTATIN | 40 MG |
| 00378654005 | LOVASTATIN | 40 MG |
| 00378654091 | LOVASTATIN | 40 MG |
| 00781121310 | LOVASTATIN | 40 MG |
| 00781121360 | LOVASTATIN | 40 MG |
| 00904558352 | LOVASTATIN | 40 MG |
| 10544024230 | LOVASTATIN | 40 MG |
| 16590094130 | LOVASTATIN | 40 MG |
| 21695053630 | LOVASTATIN | 40 MG |
| 21695053690 | LOVASTATIN | 40 MG |
| 23490584001 | LOVASTATIN | 40 MG |
| 23490584002 | LOVASTATIN | 40 MG |
| 33261054900 | LOVASTATIN | 40 MG |
| 33261054902 | LOVASTATIN | 40 MG |
| 33261054930 | LOVASTATIN | 40 MG |
| 33261054960 | LOVASTATIN | 40 MG |
| 33261054990 | LOVASTATIN | 40 MG |
| 33358022630 | LOVASTATIN | 40 MG |
| 42254002530 | LOVASTATIN | 40 MG |
| 42254002590 | LOVASTATIN | 40 MG |
| 42291037710 | LOVASTATIN | 40 MG |
| 42291037790 | LOVASTATIN | 40 MG |
| 43063054814 | LOVASTATIN | 40 MG |
| 43063054830 | LOVASTATIN | 40 MG |
| 43063054890 | LOVASTATIN | 40 MG |
| 43063093930 | LOVASTATIN | 40 MG |
| 45963063501 | LOVASTATIN | 40 MG |
| 45963063504 | LOVASTATIN | 40 MG |
| 49884075601 | LOVASTATIN | 40 MG |
| 49884075602 | LOVASTATIN | 40 MG |
| 49884075610 | LOVASTATIN | 40 MG |
| 49999047100 | LOVASTATIN | 40 MG |
| 49999047130 | LOVASTATIN | 40 MG |
| 49999047160 | LOVASTATIN | 40 MG |
| 49999047190 | LOVASTATIN | 40 MG |
| 50090076200 | LOVASTATIN | 40 MG |
| 50090076202 | LOVASTATIN | 40 MG |
| 50090321600 | LOVASTATIN | 40 MG |
| 50090321602 | LOVASTATIN | 40 MG |
| 50268051211 | LOVASTATIN AVPAK | 40 MG |
| 50268051215 | LOVASTATIN AVPAK | 40 MG |
| 51079097601 | LOVASTATIN | 40 MG |
| 51079097620 | LOVASTATIN | 40 MG |
| 51079097630 | LOVASTATIN | 40 MG |
| 51079097656 | LOVASTATIN | 40 MG |
| 51655028124 | LOVASTATIN | 40 MG |
| 53489060901 | LOVASTATIN | 40 MG |
| 53489060906 | LOVASTATIN | 40 MG |
| 53489060910 | LOVASTATIN | 40 MG |
| 54458084416 | LOVASTATIN | 40 MG |
| 54458087010 | LOVASTATIN | 40 MG |
| 54458091410 | LOVASTATIN | 40 MG |
| 54458093610 | LOVASTATIN | 40 MG |
| 54458093616 | LOVASTATIN | 40 MG |
| 54458098210 | LOVASTATIN | 40 MG |
| 54569325600 | MEVACOR | 40 MG |
| 54569325601 | MEVACOR | 40 MG |
| 54569534700 | LOVASTATIN | 40 MG |
| 54569534702 | LOVASTATIN | 40 MG |
| 54868108700 | MEVACOR | 40 MG |
| 54868108701 | MEVACOR | 40 MG |
| 54868477400 | LOVASTATIN | 40 MG |
| 54868477401 | LOVASTATIN | 40 MG |
| 54868477402 | LOVASTATIN | 40 MG |
| 54868477403 | LOVASTATIN | 40 MG |
| 54868551300 | ALTOPREV | 40 MG |
| 55045301501 | LOVASTATIN | 40 MG |
| 55045301508 | LOVASTATIN | 40 MG |
| 55048039430 | LOVASTATIN | 40 MG |
| 55048039490 | LOVASTATIN | 40 MG |
| 55289054830 | MEVACOR | 40 MG |
| 55289069214 | LOVASTATIN | 40 MG |
| 55289069230 | LOVASTATIN | 40 MG |
| 55289069290 | LOVASTATIN | 40 MG |
| 55887036930 | LOVASTATIN | 40 MG |
| 55887036960 | LOVASTATIN | 40 MG |
| 55887036990 | LOVASTATIN | 40 MG |
| 57866650001 | LOVASTATIN | 40 MG |
| 58016092200 | LOVASTATIN | 40 MG |
| 58016092202 | LOVASTATIN | 40 MG |
| 58016092230 | LOVASTATIN | 40 MG |
| 58016092260 | LOVASTATIN | 40 MG |
| 58016092290 | LOVASTATIN | 40 MG |
| 59630062930 | ALTOPREV | 40 MG |
| 60429025010 | LOVASTATIN | 40 MG |
| 60429025060 | LOVASTATIN | 40 MG |
| 60429025090 | LOVASTATIN | 40 MG |
| 60429040210 | LOVASTATIN | 40 MG |
| 60429040260 | LOVASTATIN | 40 MG |
| 60429040290 | LOVASTATIN | 40 MG |
| 60505017900 | LOVASTATIN | 40 MG |
| 60760037230 | LOVASTATIN | 40 MG |
| 61442014301 | LOVASTATIN | 40 MG |
| 61442014305 | LOVASTATIN | 40 MG |
| 61442014310 | LOVASTATIN | 40 MG |
| 61442014360 | LOVASTATIN | 40 MG |
| 61919094190 | LOVASTATIN | 40 MG |
| 62022062930 | ALTOPREV | 40 MG |
| 62022078030 | ALTOCOR | 40 MG |
| 62037079301 | LOVASTATIN | 40 MG |
| 62037079360 | LOVASTATIN | 40 MG |
| 63629178401 | LOVASTATIN | 40 MG |
| 63629178402 | LOVASTATIN | 40 MG |
| 63739028203 | LOVASTATIN | 40 MG |
| 63739028210 | LOVASTATIN | 40 MG |
| 66267056130 | LOVASTATIN | 40 MG |
| 66267056160 | LOVASTATIN | 40 MG |
| 66267056190 | LOVASTATIN | 40 MG |
| 66336041205 | LOVASTATIN | 40 MG |
| 66336041230 | LOVASTATIN | 40 MG |
| 66336041290 | LOVASTATIN | 40 MG |
| 67046045130 | LOVASTATIN | 40 MG |
| 68001021400 | LOVASTATIN | 40 MG |
| 68001021406 | LOVASTATIN | 40 MG |
| 68001021408 | LOVASTATIN | 40 MG |
| 68001031600 | LOVASTATIN | 40 MG |
| 68001031608 | LOVASTATIN | 40 MG |
| 68084013301 | LOVASTATIN | 40 MG |
| 68084056001 | LOVASTATIN | 40 MG |
| 68115065800 | LOVASTATIN | 40 MG |
| 68180046901 | LOVASTATIN | 40 MG |
| 68180046903 | LOVASTATIN | 40 MG |
| 68180046905 | LOVASTATIN | 40 MG |
| 68180046907 | LOVASTATIN | 40 MG |
| 68645056790 | LOVASTATIN | 40 MG |
| 70515062930 | ALTOPREV | 40 MG |
| 71205019930 | LOVASTATIN | 40 MG |
| 71205019990 | LOVASTATIN | 40 MG |
| 71335004501 | LOVASTATIN | 40 MG |
| 00003019450 | PRAVACHOL | 40 MG |
| 00003519410 | PRAVACHOL | 40 MG |
| 00003519433 | PRAVACHOL | 40 MG |
| 00093720210 | PRAVASTATIN SODIUM | 40 MG |
| 00093720298 | PRAVASTATIN SODIUM | 40 MG |
| 00378055777 | PRAVASTATIN SODIUM | 40 MG |
| 00378824010 | PRAVASTATIN SODIUM | 40 MG |
| 00378824077 | PRAVASTATIN SODIUM | 40 MG |
| 00591001610 | PRAVASTATIN SODIUM | 40 MG |
| 00591001619 | PRAVASTATIN SODIUM | 40 MG |
| 00781523410 | PRAVASTATIN SODIUM | 40 MG |
| 00781523492 | PRAVASTATIN SODIUM | 40 MG |
| 00904589361 | PRAVASTATIN SODIUM | 40 MG |
| 00904611561 | PRAVASTATIN SODIUM | 40 MG |
| 10544050730 | PRAVASTATIN SODIUM | 40 MG |
| 12280033515 | PRAVASTATIN | 40 MG |
| 12280033530 | PRAVASTATIN | 40 MG |
| 12280033590 | PRAVASTATIN | 40 MG |
| 13411011901 | PRAVACHOL | 40 MG |
| 13411011902 | PRAVACHOL | 40 MG |
| 13411011903 | PRAVACHOL | 40 MG |
| 13411011906 | PRAVACHOL | 40 MG |
| 13411011909 | PRAVACHOL | 40 MG |
| 16252052850 | PRAVASTATIN SODIUM | 40 MG |
| 16252052890 | PRAVASTATIN SODIUM | 40 MG |
| 16590054630 | PRAVASTATIN SODIUM | 40 MG |
| 16590054660 | PRAVASTATIN SODIUM | 40 MG |
| 16590054690 | PRAVASTATIN SODIUM | 40 MG |
| 16729001015 | PRAVASTATIN SODIUM | 40 MG |
| 16729001016 | PRAVASTATIN SODIUM | 40 MG |
| 16729001017 | PRAVASTATIN SODIUM | 40 MG |
| 21695018030 | PRAVASTATIN SODIUM | 40 MG |
| 21695018090 | PRAVASTATIN SODIUM | 40 MG |
| 23490935203 | PRAVASTATIN SODIUM | 40 MG |
| 23490935206 | PRAVASTATIN SODIUM | 40 MG |
| 23490935209 | PRAVASTATIN SODIUM | 40 MG |
| 33261086800 | PRAVASTATIN SODIUM | 40 MG |
| 33261086830 | PRAVASTATIN SODIUM | 40 MG |
| 33261086860 | PRAVASTATIN SODIUM | 40 MG |
| 33261086890 | PRAVASTATIN SODIUM | 40 MG |
| 35356012530 | PRAVASTATIN SODIUM | 40 MG |
| 42254013130 | PRAVASTATIN SODIUM | 40 MG |
| 42254013190 | PRAVASTATIN SODIUM | 40 MG |
| 42254043430 | PRAVASTATIN SODIUM | 40 MG |
| 42291066810 | PRAVASTATIN SODIUM | 40 MG |
| 42291066890 | PRAVASTATIN SODIUM | 40 MG |
| 43063019530 | PRAVASTATIN SODIUM | 40 MG |
| 43063044430 | PRAVASTATIN SODIUM | 40 MG |
| 43063080830 | PRAVASTATIN SODIUM | 40 MG |
| 49884018009 | PRAVASTATIN SODIUM | 40 MG |
| 49884018010 | PRAVASTATIN SODIUM | 40 MG |
| 50090202400 | PRAVASTATIN SODIUM | 40 MG |
| 50090202401 | PRAVASTATIN SODIUM | 40 MG |
| 50111076403 | PRAVASTATIN SODIUM | 40 MG |
| 50111076417 | PRAVASTATIN SODIUM | 40 MG |
| 51079078201 | PRAVASTATIN SODIUM | 40 MG |
| 51079078220 | PRAVASTATIN SODIUM | 40 MG |
| 51655007252 | PRAVASTATIN SODIUM | 40 MG |
| 51655007352 | PRAVASTATIN SODIUM | 40 MG |
| 54458086710 | PRAVASTATIN SODIUM | 40 MG |
| 54458092510 | PRAVASTATIN SODIUM | 40 MG |
| 54458092516 | PRAVASTATIN SODIUM | 40 MG |
| 54458098510 | PRAVASTATIN SODIUM | 40 MG |
| 54569461000 | PRAVACHOL | 40 MG |
| 54569579400 | PRAVASTATIN SODIUM | 40 MG |
| 54569579401 | PRAVASTATIN SODIUM | 40 MG |
| 54868327000 | PRAVACHOL | 40 MG |
| 54868327001 | PRAVACHOL | 40 MG |
| 54868327002 | PRAVACHOL | 40 MG |
| 54868557800 | PRAVASTATIN SODIUM | 40 MG |
| 54868557801 | PRAVASTATIN SODIUM | 40 MG |
| 54868557802 | PRAVASTATIN SODIUM | 40 MG |
| 55048059630 | PRAVASTATIN SODIUM | 40 MG |
| 55111023105 | PRAVASTATIN SODIUM | 40 MG |
| 55111023190 | PRAVASTATIN SODIUM | 40 MG |
| 55289087330 | PRAVACHOL | 40 MG |
| 55887019290 | PRAVASTATIN | 40 MG |
| 57237016605 | PRAVASTATIN SODIUM | 40 MG |
| 57237016690 | PRAVASTATIN SODIUM | 40 MG |
| 57866393201 | PRAVASTATIN | 40 MG |
| 58016001200 | PRAVASTATIN | 40 MG |
| 58016001230 | PRAVASTATIN | 40 MG |
| 58016001260 | PRAVASTATIN | 40 MG |
| 58016001290 | PRAVASTATIN | 40 MG |
| 58864074315 | PRAVACHOL | 40 MG |
| 58864074330 | PRAVACHOL | 40 MG |
| 60429036905 | PRAVASTATIN SODIUM | 40 MG |
| 60429036945 | PRAVASTATIN SODIUM | 40 MG |
| 60429036990 | PRAVASTATIN SODIUM | 40 MG |
| 60505017007 | PRAVASTATIN SODIUM | 40 MG |
| 60505017008 | PRAVASTATIN SODIUM | 40 MG |
| 60505017009 | PRAVASTATIN SODIUM | 40 MG |
| 60687019001 | PRAVASTATIN SODIUM | 40 MG |
| 60687019011 | PRAVASTATIN SODIUM | 40 MG |
| 60760007830 | PRAVASTATIN SODIUM | 40 MG |
| 60760007890 | PRAVASTATIN SODIUM | 40 MG |
| 60760042290 | PRAVASTATIN SODIUM | 40 MG |
| 60760052890 | PRAVASTATIN SODIUM | 40 MG |
| 61919054690 | PRAVASTATIN SODIUM | 40 MG |
| 61919070830 | PRAVASTATIN SODIUM | 40 MG |
| 63304059790 | PRAVASTATIN SODIUM | 40 MG |
| 63629160601 | PRAVASTATIN SODIUM | 40 MG |
| 63629160602 | PRAVASTATIN SODIUM | 40 MG |
| 66105012201 | PRAVACHOL | 40 MG |
| 66105012203 | PRAVACHOL | 40 MG |
| 66105012206 | PRAVACHOL | 40 MG |
| 66105012209 | PRAVACHOL | 40 MG |
| 66105012215 | PRAVACHOL | 40 MG |
| 66336081330 | PRAVASTATIN SODIUM | 40 MG |
| 66336081390 | PRAVASTATIN SODIUM | 40 MG |
| 68084018801 | PRAVASTATIN SODIUM | 40 MG |
| 68084050201 | PRAVASTATIN SODIUM | 40 MG |
| 68084050211 | PRAVASTATIN SODIUM | 40 MG |
| 68115066490 | PRAVACHOL | 40 MG |
| 68180048702 | PRAVASTATIN SODIUM | 40 MG |
| 68180048709 | PRAVASTATIN SODIUM | 40 MG |
| 68382007205 | PRAVASTATIN SODIUM | 40 MG |
| 68382007216 | PRAVASTATIN SODIUM | 40 MG |
| 68462019705 | PRAVASTATIN SODIUM | 40 MG |
| 68462019790 | PRAVASTATIN SODIUM | 40 MG |
| 68788725301 | PRAVASTATIN SODIUM | 40 MG |
| 68788725302 | PRAVASTATIN SODIUM | 40 MG |
| 68788725303 | PRAVASTATIN SODIUM | 40 MG |
| 68788725306 | PRAVASTATIN SODIUM | 40 MG |
| 68788725308 | PRAVASTATIN SODIUM | 40 MG |
| 68788725309 | PRAVASTATIN SODIUM | 40 MG |
| 70934016130 | PRAVASTATIN SODIUM | 40 MG |
| 71205012130 | PRAVASTATIN SODIUM | 40 MG |
| 76519120009 | PRAVASTATIN SODIUM | 40 MG |
| 00093757356 | ROSUVASTATIN CALCIUM | 40 MG |
| 00310075430 | CRESTOR | 40 MG |
| 00378223293 | ROSUVASTATIN CALCIUM | 40 MG |
| 00781540331 | ROSUVASTATIN CALCIUM | 40 MG |
| 00904660561 | ROSUVASTATIN CALCIUM | 40 MG |
| 00904678161 | ROSUVASTATIN CALCIUM | 40 MG |
| 13668018230 | ROSUVASTATIN CALCIUM | 40 MG |
| 16252061830 | ROSUVASTATIN CALCIUM | 40 MG |
| 16252061850 | ROSUVASTATIN CALCIUM | 40 MG |
| 16252061890 | ROSUVASTATIN CALCIUM | 40 MG |
| 16729028710 | ROSUVASTATIN CALCIUM | 40 MG |
| 16729028715 | ROSUVASTATIN CALCIUM | 40 MG |
| 16729028717 | ROSUVASTATIN CALCIUM | 40 MG |
| 21695065930 | CRESTOR | 40 MG |
| 27808015801 | ROSUVASTATIN CALCIUM | 40 MG |
| 31722088530 | ROSUVASTATIN CALCIUM | 40 MG |
| 35356041330 | CRESTOR | 40 MG |
| 42291074590 | ROSUVASTATIN CALCIUM | 40 MG |
| 42292003201 | ROSUVASTATIN CALCIUM | 40 MG |
| 42292003220 | ROSUVASTATIN CALCIUM | 40 MG |
| 47335058583 | ROSUVASTATIN CALCIUM | 40 MG |
| 47335098783 | EZALLOR SPRINKLE | 40 MG |
| 49884026311 | ROSUVASTATIN CALCIUM | 40 MG |
| 50090272500 | ROSUVASTATIN CALCIUM | 40 MG |
| 50090272501 | ROSUVASTATIN CALCIUM | 40 MG |
| 50090343700 | ROSUVASTATIN CALCIUM | 40 MG |
| 50090343701 | ROSUVASTATIN CALCIUM | 40 MG |
| 50268071111 | ROSUVASTATIN CALCIUM AVPAK | 40 MG |
| 50268071115 | ROSUVASTATIN CALCIUM AVPAK | 40 MG |
| 51407015630 | ROSUVASTATIN CALCIUM | 40 MG |
| 54569605401 | CRESTOR | 40 MG |
| 54569667600 | ROSUVASTATIN CALCIUM | 40 MG |
| 54569667601 | ROSUVASTATIN CALCIUM | 40 MG |
| 54868189000 | CRESTOR | 40 MG |
| 54868189001 | CRESTOR | 40 MG |
| 55700053490 | ROSUVASTATIN CALCIUM | 40 MG |
| 57237017105 | ROSUVASTATIN CALCIUM | 40 MG |
| 57237017130 | ROSUVASTATIN CALCIUM | 40 MG |
| 57237017190 | ROSUVASTATIN CALCIUM | 40 MG |
| 58016007100 | CRESTOR | 40 MG |
| 58016007130 | CRESTOR | 40 MG |
| 58016007160 | CRESTOR | 40 MG |
| 58016007190 | CRESTOR | 40 MG |
| 60429084530 | ROSUVASTATIN CALCIUM | 40 MG |
| 60505450503 | ROSUVASTATIN CALCIUM | 40 MG |
| 63187086930 | ROSUVASTATIN CALCIUM | 40 MG |
| 63187087230 | ROSUVASTATIN CALCIUM | 40 MG |
| 65862029630 | ROSUVASTATIN CALCIUM | 40 MG |
| 67877044230 | ROSUVASTATIN CALCIUM | 40 MG |
| 67877044290 | ROSUVASTATIN CALCIUM | 40 MG |
| 68258698303 | CRESTOR | 40 MG |
| 68462026430 | ROSUVASTATIN CALCIUM | 40 MG |
| 70377000911 | ROSUVASTATIN CALCIUM | 40 MG |
| 70377000912 | ROSUVASTATIN CALCIUM | 40 MG |
| 70377000913 | ROSUVASTATIN CALCIUM | 40 MG |
| 71205007830 | ROSUVASTATIN CALCIUM | 40 MG |
| 71205017630 | ROSUVASTATIN CALCIUM | 40 MG |
| 71205017690 | ROSUVASTATIN CALCIUM | 40 MG |
| 71335039001 | ROSUVASTATIN CALCIUM | 40 MG |
| 72205000530 | ROSUVASTATIN CALCIUM | 40 MG |
| 72205000590 | ROSUVASTATIN CALCIUM | 40 MG |
| 72205000599 | ROSUVASTATIN CALCIUM | 40 MG |
| 76519115303 | ROSUVASTATIN CALCIUM | 40 MG |
| 00006074928 | ZOCOR | 40 MG |
| 00006074931 | ZOCOR | 40 MG |
| 00006074954 | ZOCOR | 40 MG |
| 00006074961 | ZOCOR | 40 MG |
| 00006074982 | ZOCOR | 40 MG |
| 00093715510 | SIMVASTATIN | 40 MG |
| 00093715519 | SIMVASTATIN | 40 MG |
| 00093715531 | SIMVASTATIN | 40 MG |
| 00093715556 | SIMVASTATIN | 40 MG |
| 00093715593 | SIMVASTATIN | 40 MG |
| 00093715598 | SIMVASTATIN | 40 MG |
| 00406206803 | SIMVASTATIN | 40 MG |
| 00406206805 | SIMVASTATIN | 40 MG |
| 00406206810 | SIMVASTATIN | 40 MG |
| 00406206860 | SIMVASTATIN | 40 MG |
| 00406206890 | SIMVASTATIN | 40 MG |
| 00781507331 | SIMVASTATIN | 40 MG |
| 00781507392 | SIMVASTATIN | 40 MG |
| 00904580261 | SIMVASTATIN | 40 MG |
| 10544048790 | SIMVASTATIN | 40 MG |
| 13411013301 | ZOCOR | 40 MG |
| 13411013303 | ZOCOR | 40 MG |
| 13411013306 | ZOCOR | 40 MG |
| 13411013309 | ZOCOR | 40 MG |
| 13411013315 | ZOCOR | 40 MG |
| 16252050830 | SIMVASTATIN | 40 MG |
| 16252050850 | SIMVASTATIN | 40 MG |
| 16252050890 | SIMVASTATIN | 40 MG |
| 16590043130 | SIMVASTATIN | 40 MG |
| 16590043190 | SIMVASTATIN | 40 MG |
| 16714068401 | SIMVASTATIN | 40 MG |
| 16714068402 | SIMVASTATIN | 40 MG |
| 16714068403 | SIMVASTATIN | 40 MG |
| 16729000610 | SIMVASTATIN | 40 MG |
| 16729000615 | SIMVASTATIN | 40 MG |
| 16729000617 | SIMVASTATIN | 40 MG |
| 21695074130 | SIMVASTATIN | 40 MG |
| 21695074190 | SIMVASTATIN | 40 MG |
| 23490935503 | SIMVASTATIN | 40 MG |
| 23490935506 | SIMVASTATIN | 40 MG |
| 23490935509 | SIMVASTATIN | 40 MG |
| 24658021310 | SIMVASTATIN | 40 MG |
| 24658021330 | SIMVASTATIN | 40 MG |
| 24658021345 | SIMVASTATIN | 40 MG |
| 24658021390 | SIMVASTATIN | 40 MG |
| 24658030310 | SIMVASTATIN | 40 MG |
| 24658030315 | SIMVASTATIN | 40 MG |
| 24658030330 | SIMVASTATIN | 40 MG |
| 24658030345 | SIMVASTATIN | 40 MG |
| 24658030390 | SIMVASTATIN | 40 MG |
| 31722051310 | SIMVASTATIN | 40 MG |
| 31722051390 | SIMVASTATIN | 40 MG |
| 33261054202 | SIMVASTATIN | 40 MG |
| 33261054230 | SIMVASTATIN | 40 MG |
| 33261054260 | SIMVASTATIN | 40 MG |
| 33261054290 | SIMVASTATIN | 40 MG |
| 35356077530 | SIMVASTATIN | 40 MG |
| 35356077590 | SIMVASTATIN | 40 MG |
| 42254003230 | SIMVASTATIN | 40 MG |
| 42254003245 | SIMVASTATIN | 40 MG |
| 42254003290 | SIMVASTATIN | 40 MG |
| 42254022530 | SIMVASTATIN | 40 MG |
| 42571004010 | SIMVASTATIN | 40 MG |
| 42571004090 | SIMVASTATIN | 40 MG |
| 43063059330 | SIMVASTATIN | 40 MG |
| 43063059390 | SIMVASTATIN | 40 MG |
| 43063072630 | SIMVASTATIN | 40 MG |
| 43063072690 | SIMVASTATIN | 40 MG |
| 45802087901 | SIMVASTATIN | 40 MG |
| 45802087965 | SIMVASTATIN | 40 MG |
| 45802087975 | SIMVASTATIN | 40 MG |
| 45802087993 | SIMVASTATIN | 40 MG |
| 45865046030 | SIMVASTATIN | 40 MG |
| 45865046051 | SIMVASTATIN | 40 MG |
| 45865046060 | SIMVASTATIN | 40 MG |
| 45865046090 | SIMVASTATIN | 40 MG |
| 49999048830 | ZOCOR | 40 MG |
| 49999090315 | SIMVASTATIN | 40 MG |
| 49999090330 | SIMVASTATIN | 40 MG |
| 49999090390 | SIMVASTATIN | 40 MG |
| 50090100001 | SIMVASTATIN | 40 MG |
| 50090100002 | SIMVASTATIN | 40 MG |
| 50090100003 | SIMVASTATIN | 40 MG |
| 50090100004 | SIMVASTATIN | 40 MG |
| 50268071511 | SIMVASTATIN AVPAK | 40 MG |
| 50268071515 | SIMVASTATIN AVPAK | 40 MG |
| 50742013910 | SIMVASTATIN | 40 MG |
| 51079039801 | SIMVASTATIN | 40 MG |
| 51079039820 | SIMVASTATIN | 40 MG |
| 51079045601 | SIMVASTATIN | 40 MG |
| 51079045620 | SIMVASTATIN | 40 MG |
| 52959011230 | ZOCOR | 40 MG |
| 52959094430 | SIMVASTATIN | 40 MG |
| 54458089410 | SIMVASTATIN | 40 MG |
| 54458093210 | SIMVASTATIN | 40 MG |
| 54458093216 | SIMVASTATIN | 40 MG |
| 54569440400 | ZOCOR | 40 MG |
| 54569583400 | SIMVASTATIN | 40 MG |
| 54569583401 | SIMVASTATIN | 40 MG |
| 54569583402 | SIMVASTATIN | 40 MG |
| 54569583403 | SIMVASTATIN | 40 MG |
| 54569583404 | SIMVASTATIN | 40 MG |
| 54868415700 | ZOCOR | 40 MG |
| 54868415701 | ZOCOR | 40 MG |
| 54868415702 | ZOCOR | 40 MG |
| 54868562900 | SIMVASTATIN | 40 MG |
| 54868562901 | SIMVASTATIN | 40 MG |
| 54868562902 | SIMVASTATIN | 40 MG |
| 54868562903 | SIMVASTATIN | 40 MG |
| 54868562904 | SIMVASTATIN | 40 MG |
| 55045310008 | ZOCOR | 40 MG |
| 55048077430 | SIMVASTATIN | 40 MG |
| 55048077490 | SIMVASTATIN | 40 MG |
| 55111020005 | SIMVASTATIN | 40 MG |
| 55111020010 | SIMVASTATIN | 40 MG |
| 55111020030 | SIMVASTATIN | 40 MG |
| 55111020090 | SIMVASTATIN | 40 MG |
| 55111074910 | SIMVASTATIN | 40 MG |
| 55111074930 | SIMVASTATIN | 40 MG |
| 55111074990 | SIMVASTATIN | 40 MG |
| 55289039530 | SIMVASTATIN | 40 MG |
| 55289039590 | SIMVASTATIN | 40 MG |
| 55289087430 | ZOCOR | 40 MG |
| 55700026530 | SIMVASTATIN | 40 MG |
| 55700026590 | SIMVASTATIN | 40 MG |
| 55700042618 | SIMVASTATIN | 40 MG |
| 55700042630 | SIMVASTATIN | 40 MG |
| 55700042690 | SIMVASTATIN | 40 MG |
| 55700055090 | SIMVASTATIN | 40 MG |
| 55887085810 | SIMVASTATIN | 40 MG |
| 55887085830 | SIMVASTATIN | 40 MG |
| 55887085860 | SIMVASTATIN | 40 MG |
| 55887085890 | SIMVASTATIN | 40 MG |
| 57866394901 | SIMVASTATIN | 40 MG |
| 57866798301 | ZOCOR | 40 MG |
| 58016000600 | SIMVASTATIN | 40 MG |
| 58016000630 | SIMVASTATIN | 40 MG |
| 58016000660 | SIMVASTATIN | 40 MG |
| 58016000690 | SIMVASTATIN | 40 MG |
| 58016036500 | ZOCOR | 40 MG |
| 58016036530 | ZOCOR | 40 MG |
| 58016036560 | ZOCOR | 40 MG |
| 58016036590 | ZOCOR | 40 MG |
| 58864068230 | ZOCOR | 40 MG |
| 60687021001 | SIMVASTATIN | 40 MG |
| 60687021011 | SIMVASTATIN | 40 MG |
| 60760000630 | SIMVASTATIN | 40 MG |
| 60760000690 | SIMVASTATIN | 40 MG |
| 61919043130 | SIMVASTATIN | 40 MG |
| 61919043190 | SIMVASTATIN | 40 MG |
| 63187044990 | SIMVASTATIN | 40 MG |
| 63304079210 | SIMVASTATIN | 40 MG |
| 63304079230 | SIMVASTATIN | 40 MG |
| 63304079290 | SIMVASTATIN | 40 MG |
| 63739042210 | SIMVASTATIN | 40 MG |
| 63739043810 | SIMVASTATIN | 40 MG |
| 63739057310 | SIMVASTATIN | 40 MG |
| 65862005322 | SIMVASTATIN | 40 MG |
| 65862005330 | SIMVASTATIN | 40 MG |
| 65862005390 | SIMVASTATIN | 40 MG |
| 65862005399 | SIMVASTATIN | 40 MG |
| 66105050601 | ZOCOR | 40 MG |
| 66105050603 | ZOCOR | 40 MG |
| 66105050606 | ZOCOR | 40 MG |
| 66105050609 | ZOCOR | 40 MG |
| 66105050610 | ZOCOR | 40 MG |
| 66336095330 | SIMVASTATIN | 40 MG |
| 66336095390 | SIMVASTATIN | 40 MG |
| 68084016401 | SIMVASTATIN | 40 MG |
| 68084051301 | SIMVASTATIN | 40 MG |
| 68115077730 | ZOCOR | 40 MG |
| 68115077790 | ZOCOR | 40 MG |
| 68180046403 | SIMVASTATIN | 40 MG |
| 68180046406 | SIMVASTATIN | 40 MG |
| 68180046409 | SIMVASTATIN | 40 MG |
| 68180048001 | SIMVASTATIN | 40 MG |
| 68180048002 | SIMVASTATIN | 40 MG |
| 68180048003 | SIMVASTATIN | 40 MG |
| 68382006805 | SIMVASTATIN | 40 MG |
| 68382006806 | SIMVASTATIN | 40 MG |
| 68382006810 | SIMVASTATIN | 40 MG |
| 68382006814 | SIMVASTATIN | 40 MG |
| 68382006816 | SIMVASTATIN | 40 MG |
| 68382006840 | SIMVASTATIN | 40 MG |
| 68645026254 | SIMVASTATIN | 40 MG |
| 68645047154 | SIMVASTATIN | 40 MG |
| 68645052754 | SIMVASTATIN | 40 MG |
| 70377000412 | SIMVASTATIN | 40 MG |
| 70377000414 | SIMVASTATIN | 40 MG |
| 70377000415 | SIMVASTATIN | 40 MG |
| 71205019230 | SIMVASTATIN | 40 MG |
| 29273040204 | FLOLIPID | 40 MG/5 ML |
| 66582032230 | LIPTRUZET | 40 MG-10 MG |
| 66582032254 | LIPTRUZET | 40 MG-10 MG |
| 00006077331 | JUVISYNC | 40 MG-100 MG |
| 00006077354 | JUVISYNC | 40 MG-100 MG |
| 00006077382 | JUVISYNC | 40 MG-100 MG |
| 00074301090 | ADVICOR | 40 MG-1000 MG |
| 54868565300 | ADVICOR | 40 MG-1000 MG |
| 54868565301 | ADVICOR | 40 MG-1000 MG |
| 60598000990 | ADVICOR | 40 MG-1000 MG |
| 00006053731 | JUVISYNC | 40 MG-50 MG |
| 00006053754 | JUVISYNC | 40 MG-50 MG |
| 00093757098 | ROSUVASTATIN CALCIUM | 5 MG |
| 00310075590 | CRESTOR | 5 MG |
| 00378220177 | ROSUVASTATIN CALCIUM | 5 MG |
| 00781540092 | ROSUVASTATIN CALCIUM | 5 MG |
| 00904660261 | ROSUVASTATIN CALCIUM | 5 MG |
| 00904677861 | ROSUVASTATIN CALCIUM | 5 MG |
| 13668017930 | ROSUVASTATIN CALCIUM | 5 MG |
| 13668017990 | ROSUVASTATIN CALCIUM | 5 MG |
| 16252061530 | ROSUVASTATIN CALCIUM | 5 MG |
| 16252061550 | ROSUVASTATIN CALCIUM | 5 MG |
| 16252061590 | ROSUVASTATIN CALCIUM | 5 MG |
| 16729028415 | ROSUVASTATIN CALCIUM | 5 MG |
| 16729028417 | ROSUVASTATIN CALCIUM | 5 MG |
| 21695075990 | CRESTOR | 5 MG |
| 27808015501 | ROSUVASTATIN CALCIUM | 5 MG |
| 31722088290 | ROSUVASTATIN CALCIUM | 5 MG |
| 35356051930 | CRESTOR | 5 MG |
| 42291074290 | ROSUVASTATIN CALCIUM | 5 MG |
| 42292002901 | ROSUVASTATIN CALCIUM | 5 MG |
| 42292002920 | ROSUVASTATIN CALCIUM | 5 MG |
| 47335058281 | ROSUVASTATIN CALCIUM | 5 MG |
| 47335098483 | EZALLOR SPRINKLE | 5 MG |
| 47463009530 | CRESTOR | 5 MG |
| 49884026009 | ROSUVASTATIN CALCIUM | 5 MG |
| 50090272200 | ROSUVASTATIN CALCIUM | 5 MG |
| 50090272201 | ROSUVASTATIN CALCIUM | 5 MG |
| 50090317600 | ROSUVASTATIN CALCIUM | 5 MG |
| 50090317601 | ROSUVASTATIN CALCIUM | 5 MG |
| 51407015390 | ROSUVASTATIN CALCIUM | 5 MG |
| 53217029730 | ROSUVASTATIN CALCIUM | 5 MG |
| 53217029790 | ROSUVASTATIN CALCIUM | 5 MG |
| 54569574600 | CRESTOR | 5 MG |
| 54569667300 | ROSUVASTATIN CALCIUM | 5 MG |
| 54569667301 | ROSUVASTATIN CALCIUM | 5 MG |
| 54868534100 | CRESTOR | 5 MG |
| 54868534101 | CRESTOR | 5 MG |
| 55048009530 | CRESTOR | 5 MG |
| 57237016805 | ROSUVASTATIN CALCIUM | 5 MG |
| 57237016890 | ROSUVASTATIN CALCIUM | 5 MG |
| 60429084290 | ROSUVASTATIN CALCIUM | 5 MG |
| 60505450209 | ROSUVASTATIN CALCIUM | 5 MG |
| 60687023401 | ROSUVASTATIN CALCIUM | 5 MG |
| 60687023411 | ROSUVASTATIN CALCIUM | 5 MG |
| 63187086090 | ROSUVASTATIN CALCIUM | 5 MG |
| 63629715801 | ROSUVASTATIN CALCIUM | 5 MG |
| 63629715802 | ROSUVASTATIN CALCIUM | 5 MG |
| 65862029390 | ROSUVASTATIN CALCIUM | 5 MG |
| 67877043990 | ROSUVASTATIN CALCIUM | 5 MG |
| 68071078430 | CRESTOR | 5 MG |
| 68258601703 | CRESTOR | 5 MG |
| 68462026190 | ROSUVASTATIN CALCIUM | 5 MG |
| 68788731002 | ROSUVASTATIN CALCIUM | 5 MG |
| 68788731003 | ROSUVASTATIN CALCIUM | 5 MG |
| 68788731006 | ROSUVASTATIN CALCIUM | 5 MG |
| 68788731009 | ROSUVASTATIN CALCIUM | 5 MG |
| 70377000612 | ROSUVASTATIN CALCIUM | 5 MG |
| 70377000613 | ROSUVASTATIN CALCIUM | 5 MG |
| 71335074901 | ROSUVASTATIN CALCIUM | 5 MG |
| 72205000290 | ROSUVASTATIN CALCIUM | 5 MG |
| 72205000299 | ROSUVASTATIN CALCIUM | 5 MG |
| 00006072628 | ZOCOR | 5 MG |
| 00006072631 | ZOCOR | 5 MG |
| 00006072654 | ZOCOR | 5 MG |
| 00006072661 | ZOCOR | 5 MG |
| 00006072682 | ZOCOR | 5 MG |
| 00093715219 | SIMVASTATIN | 5 MG |
| 00093715256 | SIMVASTATIN | 5 MG |
| 00093715293 | SIMVASTATIN | 5 MG |
| 00093715298 | SIMVASTATIN | 5 MG |
| 00406206503 | SIMVASTATIN | 5 MG |
| 00406206505 | SIMVASTATIN | 5 MG |
| 00406206510 | SIMVASTATIN | 5 MG |
| 00406206560 | SIMVASTATIN | 5 MG |
| 00406206590 | SIMVASTATIN | 5 MG |
| 00781507031 | SIMVASTATIN | 5 MG |
| 00781507092 | SIMVASTATIN | 5 MG |
| 13411016101 | ZOCOR | 5 MG |
| 13411016103 | ZOCOR | 5 MG |
| 13411016106 | ZOCOR | 5 MG |
| 13411016109 | ZOCOR | 5 MG |
| 13411016115 | ZOCOR | 5 MG |
| 16252050530 | SIMVASTATIN | 5 MG |
| 16252050550 | SIMVASTATIN | 5 MG |
| 16252050590 | SIMVASTATIN | 5 MG |
| 16714068101 | SIMVASTATIN | 5 MG |
| 16714068102 | SIMVASTATIN | 5 MG |
| 16729015610 | SIMVASTATIN | 5 MG |
| 16729015615 | SIMVASTATIN | 5 MG |
| 16729015617 | SIMVASTATIN | 5 MG |
| 21695073890 | SIMVASTATIN | 5 MG |
| 23490935603 | SIMVASTATIN | 5 MG |
| 23490935606 | SIMVASTATIN | 5 MG |
| 23490935609 | SIMVASTATIN | 5 MG |
| 24658021010 | SIMVASTATIN | 5 MG |
| 24658021030 | SIMVASTATIN | 5 MG |
| 24658021045 | SIMVASTATIN | 5 MG |
| 24658021090 | SIMVASTATIN | 5 MG |
| 24658030010 | SIMVASTATIN | 5 MG |
| 24658030030 | SIMVASTATIN | 5 MG |
| 24658030045 | SIMVASTATIN | 5 MG |
| 24658030090 | SIMVASTATIN | 5 MG |
| 31722051010 | SIMVASTATIN | 5 MG |
| 31722051090 | SIMVASTATIN | 5 MG |
| 42571000590 | SIMVASTATIN | 5 MG |
| 45802092465 | SIMVASTATIN | 5 MG |
| 49999090090 | SIMVASTATIN | 5 MG |
| 50090138700 | SIMVASTATIN | 5 MG |
| 50090254300 | SIMVASTATIN | 5 MG |
| 50268071211 | SIMVASTATIN AVPAK | 5 MG |
| 50268071215 | SIMVASTATIN AVPAK | 5 MG |
| 54569645000 | SIMVASTATIN | 5 MG |
| 54868606600 | SIMVASTATIN | 5 MG |
| 55111019705 | SIMVASTATIN | 5 MG |
| 55111019730 | SIMVASTATIN | 5 MG |
| 55111019790 | SIMVASTATIN | 5 MG |
| 55111072610 | SIMVASTATIN | 5 MG |
| 55111072630 | SIMVASTATIN | 5 MG |
| 55111072690 | SIMVASTATIN | 5 MG |
| 58864073930 | ZOCOR | 5 MG |
| 63304078910 | SIMVASTATIN | 5 MG |
| 63304078930 | SIMVASTATIN | 5 MG |
| 63304078990 | SIMVASTATIN | 5 MG |
| 63739041910 | SIMVASTATIN | 5 MG |
| 63739043510 | SIMVASTATIN | 5 MG |
| 63739057010 | SIMVASTATIN | 5 MG |
| 65862005030 | SIMVASTATIN | 5 MG |
| 65862005090 | SIMVASTATIN | 5 MG |
| 65862005099 | SIMVASTATIN | 5 MG |
| 68084016101 | SIMVASTATIN | 5 MG |
| 68084051001 | SIMVASTATIN | 5 MG |
| 68180048206 | SIMVASTATIN | 5 MG |
| 68180048209 | SIMVASTATIN | 5 MG |
| 68258605003 | SIMVASTATIN | 5 MG |
| 68258698509 | SIMVASTATIN | 5 MG |
| 68382006505 | SIMVASTATIN | 5 MG |
| 68382006506 | SIMVASTATIN | 5 MG |
| 68382006510 | SIMVASTATIN | 5 MG |
| 68382006514 | SIMVASTATIN | 5 MG |
| 68382006516 | SIMVASTATIN | 5 MG |
| 70377000112 | SIMVASTATIN | 5 MG |
| 70377000114 | SIMVASTATIN | 5 MG |
| 70377000115 | SIMVASTATIN | 5 MG |
| 71205007090 | SIMVASTATIN | 5 MG |
| 00069215030 | CADUET | 5 MG-10 MG |
| 00378451305 | AMLODIPINE BESYLATE-ATORVASTATIN CA | 5 MG-10 MG |
| 00378451393 | AMLODIPINE BESYLATE-ATORVASTATIN CA | 5 MG-10 MG |
| 00378616405 | AMLODIPINE BESYLATE-ATORVASTATIN CA | 5 MG-10 MG |
| 00378616477 | AMLODIPINE BESYLATE-ATORVASTATIN CA | 5 MG-10 MG |
| 00378616493 | AMLODIPINE BESYLATE-ATORVASTATIN CA | 5 MG-10 MG |
| 12280039930 | CADUET | 5 MG-10 MG |
| 43598032230 | AMLODIPINE BESYLATE-ATORVASTATIN CA | 5 MG-10 MG |
| 43598032290 | AMLODIPINE BESYLATE-ATORVASTATIN CA | 5 MG-10 MG |
| 49999098930 | CADUET | 5 MG-10 MG |
| 54569570400 | CADUET | 5 MG-10 MG |
| 54868328700 | CADUET | 5 MG-10 MG |
| 54868328701 | CADUET | 5 MG-10 MG |
| 59762672001 | AMLODIPINE BESYLATE-ATORVASTATIN CA | 5 MG-10 MG |
| 59762672005 | AMLODIPINE BESYLATE-ATORVASTATIN CA | 5 MG-10 MG |
| 59762672007 | AMLODIPINE BESYLATE-ATORVASTATIN CA | 5 MG-10 MG |
| 63304058730 | AMLODIPINE BESYLATE-ATORVASTATIN CA | 5 MG-10 MG |
| 00069217030 | CADUET | 5 MG-20 MG |
| 00378451405 | AMLODIPINE BESYLATE-ATORVASTATIN CA | 5 MG-20 MG |
| 00378451493 | AMLODIPINE BESYLATE-ATORVASTATIN CA | 5 MG-20 MG |
| 00378616505 | AMLODIPINE BESYLATE-ATORVASTATIN CA | 5 MG-20 MG |
| 00378616577 | AMLODIPINE BESYLATE-ATORVASTATIN CA | 5 MG-20 MG |
| 00378616593 | AMLODIPINE BESYLATE-ATORVASTATIN CA | 5 MG-20 MG |
| 43598031930 | AMLODIPINE BESYLATE-ATORVASTATIN CA | 5 MG-20 MG |
| 43598031990 | AMLODIPINE BESYLATE-ATORVASTATIN CA | 5 MG-20 MG |
| 54868120700 | CADUET | 5 MG-20 MG |
| 54868120701 | CADUET | 5 MG-20 MG |
| 59762672101 | AMLODIPINE BESYLATE-ATORVASTATIN CA | 5 MG-20 MG |
| 59762672105 | AMLODIPINE BESYLATE-ATORVASTATIN CA | 5 MG-20 MG |
| 59762672107 | AMLODIPINE BESYLATE-ATORVASTATIN CA | 5 MG-20 MG |
| 63304058830 | AMLODIPINE BESYLATE-ATORVASTATIN CA | 5 MG-20 MG |
| 00069219030 | CADUET | 5 MG-40 MG |
| 00378451505 | AMLODIPINE BESYLATE-ATORVASTATIN CA | 5 MG-40 MG |
| 00378451593 | AMLODIPINE BESYLATE-ATORVASTATIN CA | 5 MG-40 MG |
| 00378616605 | AMLODIPINE BESYLATE-ATORVASTATIN CA | 5 MG-40 MG |
| 00378616677 | AMLODIPINE BESYLATE-ATORVASTATIN CA | 5 MG-40 MG |
| 00378616693 | AMLODIPINE BESYLATE-ATORVASTATIN CA | 5 MG-40 MG |
| 43598031630 | AMLODIPINE BESYLATE-ATORVASTATIN CA | 5 MG-40 MG |
| 43598031690 | AMLODIPINE BESYLATE-ATORVASTATIN CA | 5 MG-40 MG |
| 54868517900 | CADUET | 5 MG-40 MG |
| 59762672201 | AMLODIPINE BESYLATE-ATORVASTATIN CA | 5 MG-40 MG |
| 59762672205 | AMLODIPINE BESYLATE-ATORVASTATIN CA | 5 MG-40 MG |
| 59762672207 | AMLODIPINE BESYLATE-ATORVASTATIN CA | 5 MG-40 MG |
| 63304058930 | AMLODIPINE BESYLATE-ATORVASTATIN CA | 5 MG-40 MG |
| 00069226030 | CADUET | 5 MG-80 MG |
| 00378451693 | AMLODIPINE BESYLATE-ATORVASTATIN CA | 5 MG-80 MG |
| 00378616777 | AMLODIPINE BESYLATE-ATORVASTATIN CA | 5 MG-80 MG |
| 00378616793 | AMLODIPINE BESYLATE-ATORVASTATIN CA | 5 MG-80 MG |
| 43598031430 | AMLODIPINE BESYLATE-ATORVASTATIN CA | 5 MG-80 MG |
| 54868542000 | CADUET | 5 MG-80 MG |
| 59762672301 | AMLODIPINE BESYLATE-ATORVASTATIN CA | 5 MG-80 MG |
| 63304049930 | AMLODIPINE BESYLATE-ATORVASTATIN CA | 5 MG-80 MG |
| 00074331290 | SIMCOR | 500 MG-20 MG |
| 54868588600 | SIMCOR | 500 MG-20 MG |
| 54868588601 | SIMCOR | 500 MG-20 MG |
| 00074345903 | SIMCOR | 500 MG-40 MG |
| 00074345990 | SIMCOR | 500 MG-40 MG |
| 54868535800 | ALTOPREV | 60 MG |
| 59630063030 | ALTOPREV | 60 MG |
| 62022063030 | ALTOPREV | 60 MG |
| 62022078130 | ALTOCOR | 60 MG |
| 70515063030 | ALTOPREV | 60 MG |
| 00074331590 | SIMCOR | 750 MG-20 MG |
| 54868590700 | SIMCOR | 750 MG-20 MG |
| 54868590701 | SIMCOR | 750 MG-20 MG |
| 00071015823 | LIPITOR | 80 MG |
| 00071015873 | LIPITOR | 80 MG |
| 00071015888 | LIPITOR | 80 MG |
| 00071015892 | LIPITOR | 80 MG |
| 00093505798 | ATORVASTATIN CALCIUM | 80 MG |
| 00378212205 | ATORVASTATIN CALCIUM | 80 MG |
| 00378212277 | ATORVASTATIN CALCIUM | 80 MG |
| 00378395305 | ATORVASTATIN CALCIUM | 80 MG |
| 00378395307 | ATORVASTATIN CALCIUM | 80 MG |
| 00378395309 | ATORVASTATIN CALCIUM | 80 MG |
| 00378395377 | ATORVASTATIN CALCIUM | 80 MG |
| 00591377705 | ATORVASTATIN CALCIUM | 80 MG |
| 00591377719 | ATORVASTATIN CALCIUM | 80 MG |
| 00781538892 | ATORVASTATIN CALCIUM | 80 MG |
| 00904629304 | ATORVASTATIN CALCIUM | 80 MG |
| 10135065305 | ATORVASTATIN CALCIUM | 80 MG |
| 12280015030 | LIPITOR | 80 MG |
| 16714087701 | ATORVASTATIN CALCIUM | 80 MG |
| 16714087702 | ATORVASTATIN CALCIUM | 80 MG |
| 16714087703 | ATORVASTATIN CALCIUM | 80 MG |
| 16729004716 | ATORVASTATIN CALCIUM | 80 MG |
| 33261099530 | ATORVASTATIN CALCIUM | 80 MG |
| 33261099560 | ATORVASTATIN CALCIUM | 80 MG |
| 33261099590 | ATORVASTATIN CALCIUM | 80 MG |
| 42254026730 | ATORVASTATIN CALCIUM | 80 MG |
| 42254026745 | ATORVASTATIN CALCIUM | 80 MG |
| 42254026790 | ATORVASTATIN CALCIUM | 80 MG |
| 42254039290 | ATORVASTATIN CALCIUM | 80 MG |
| 42291014650 | ATORVASTATIN CALCIUM | 80 MG |
| 42291014690 | ATORVASTATIN CALCIUM | 80 MG |
| 49999088230 | LIPITOR | 80 MG |
| 49999088290 | LIPITOR | 80 MG |
| 50090126400 | ATORVASTATIN CALCIUM | 80 MG |
| 50090126401 | ATORVASTATIN CALCIUM | 80 MG |
| 50090126500 | ATORVASTATIN CALCIUM | 80 MG |
| 50090126501 | ATORVASTATIN CALCIUM | 80 MG |
| 50268009611 | ATORVASTATIN CALCIUM AVPAK | 80 MG |
| 50268009612 | ATORVASTATIN CALCIUM AVPAK | 80 MG |
| 51079021101 | ATORVASTATIN CALCIUM | 80 MG |
| 51079021103 | ATORVASTATIN CALCIUM | 80 MG |
| 51079041201 | ATORVASTATIN CALCIUM | 80 MG |
| 51079041203 | ATORVASTATIN CALCIUM | 80 MG |
| 51407008105 | ATORVASTATIN CALCIUM | 80 MG |
| 51407008190 | ATORVASTATIN CALCIUM | 80 MG |
| 51655088030 | ATORVASTATIN CALCIUM | 80 MG |
| 53217031930 | ATORVASTATIN CALCIUM | 80 MG |
| 53217031990 | ATORVASTATIN CALCIUM | 80 MG |
| 54569538200 | LIPITOR | 80 MG |
| 54569628500 | ATORVASTATIN CALCIUM | 80 MG |
| 54569628501 | ATORVASTATIN CALCIUM | 80 MG |
| 54868493400 | LIPITOR | 80 MG |
| 54868493401 | LIPITOR | 80 MG |
| 54868493402 | LIPITOR | 80 MG |
| 54868493403 | LIPITOR | 80 MG |
| 54868632200 | ATORVASTATIN CALCIUM | 80 MG |
| 55111012405 | ATORVASTATIN CALCIUM | 80 MG |
| 55111012490 | ATORVASTATIN CALCIUM | 80 MG |
| 55700003430 | ATORVASTATIN CALCIUM | 80 MG |
| 58016005100 | LIPITOR | 80 MG |
| 58016005130 | LIPITOR | 80 MG |
| 58016005160 | LIPITOR | 80 MG |
| 58016005190 | LIPITOR | 80 MG |
| 58864083430 | LIPITOR | 80 MG |
| 59762015801 | ATORVASTATIN CALCIUM | 80 MG |
| 59762015802 | ATORVASTATIN CALCIUM | 80 MG |
| 60429032601 | ATORVASTATIN CALCIUM | 80 MG |
| 60429032605 | ATORVASTATIN CALCIUM | 80 MG |
| 60429032633 | ATORVASTATIN CALCIUM | 80 MG |
| 60429032690 | ATORVASTATIN CALCIUM | 80 MG |
| 60505267108 | ATORVASTATIN CALCIUM | 80 MG |
| 60505267109 | ATORVASTATIN CALCIUM | 80 MG |
| 60760035630 | ATORVASTATIN CALCIUM | 80 MG |
| 62175089741 | ATORVASTATIN CALCIUM | 80 MG |
| 62175089746 | ATORVASTATIN CALCIUM | 80 MG |
| 63187065690 | ATORVASTATIN CALCIUM | 80 MG |
| 63187090790 | ATORVASTATIN CALCIUM | 80 MG |
| 63304083005 | ATORVASTATIN CALCIUM | 80 MG |
| 63304083090 | ATORVASTATIN CALCIUM | 80 MG |
| 63629336601 | LIPITOR | 80 MG |
| 63629336602 | LIPITOR | 80 MG |
| 63629336603 | LIPITOR | 80 MG |
| 63629336604 | LIPITOR | 80 MG |
| 67877051405 | ATORVASTATIN CALCIUM | 80 MG |
| 67877051490 | ATORVASTATIN CALCIUM | 80 MG |
| 68084059025 | ATORVASTATIN CALCIUM | 80 MG |
| 68084059095 | ATORVASTATIN CALCIUM | 80 MG |
| 68382025210 | ATORVASTATIN CALCIUM | 80 MG |
| 68382025216 | ATORVASTATIN CALCIUM | 80 MG |
| 68645041854 | ATORVASTATIN CALCIUM | 80 MG |
| 68645046154 | ATORVASTATIN CALCIUM | 80 MG |
| 68645049554 | ATORVASTATIN CALCIUM | 80 MG |
| 69097091105 | ATORVASTATIN CALCIUM | 80 MG |
| 69097091112 | ATORVASTATIN CALCIUM | 80 MG |
| 69097094705 | ATORVASTATIN CALCIUM | 80 MG |
| 69097094712 | ATORVASTATIN CALCIUM | 80 MG |
| 70377003012 | ATORVASTATIN CALCIUM | 80 MG |
| 70377003014 | ATORVASTATIN CALCIUM | 80 MG |
| 71205009890 | ATORVASTATIN CALCIUM | 80 MG |
| 71335029501 | ATORVASTATIN CALCIUM | 80 MG |
| 71335029502 | ATORVASTATIN CALCIUM | 80 MG |
| 71335029503 | ATORVASTATIN CALCIUM | 80 MG |
| 71335058301 | ATORVASTATIN CALCIUM | 80 MG |
| 71335058302 | ATORVASTATIN CALCIUM | 80 MG |
| 71335058303 | ATORVASTATIN CALCIUM | 80 MG |
| 72205002505 | ATORVASTATIN CALCIUM | 80 MG |
| 72205002590 | ATORVASTATIN CALCIUM | 80 MG |
| 76519108403 | ATORVASTATIN CALCIUM | 80 MG |
| 00078035405 | LESCOL XL | 80 MG |
| 00078035415 | LESCOL XL | 80 MG |
| 00093744601 | FLUVASTATIN SODIUM | 80 MG |
| 00093744656 | FLUVASTATIN SODIUM | 80 MG |
| 00378512101 | FLUVASTATIN SODIUM | 80 MG |
| 00378512193 | FLUVASTATIN SODIUM | 80 MG |
| 00781537001 | FLUVASTATIN SODIUM | 80 MG |
| 00781537031 | FLUVASTATIN SODIUM | 80 MG |
| 00781801701 | FLUVASTATIN SODIUM | 80 MG |
| 00781801731 | FLUVASTATIN SODIUM | 80 MG |
| 54569549800 | LESCOL XL | 80 MG |
| 54868460100 | LESCOL XL | 80 MG |
| 00003519510 | PRAVACHOL | 80 MG |
| 00003519533 | PRAVACHOL | 80 MG |
| 00093727010 | PRAVASTATIN SODIUM | 80 MG |
| 00093727098 | PRAVASTATIN SODIUM | 80 MG |
| 00378055377 | PRAVASTATIN SODIUM | 80 MG |
| 00378828005 | PRAVASTATIN SODIUM | 80 MG |
| 00378828077 | PRAVASTATIN SODIUM | 80 MG |
| 00591001905 | PRAVASTATIN SODIUM | 80 MG |
| 00591001919 | PRAVASTATIN SODIUM | 80 MG |
| 00781523592 | PRAVASTATIN SODIUM | 80 MG |
| 16252052950 | PRAVASTATIN SODIUM | 80 MG |
| 16252052990 | PRAVASTATIN SODIUM | 80 MG |
| 16729001115 | PRAVASTATIN SODIUM | 80 MG |
| 16729001116 | PRAVASTATIN SODIUM | 80 MG |
| 33261095300 | PRAVASTATIN SODIUM | 80 MG |
| 33261095330 | PRAVASTATIN SODIUM | 80 MG |
| 33261095360 | PRAVASTATIN SODIUM | 80 MG |
| 33261095390 | PRAVASTATIN SODIUM | 80 MG |
| 42291066910 | PRAVASTATIN SODIUM | 80 MG |
| 42291066945 | PRAVASTATIN SODIUM | 80 MG |
| 42291066990 | PRAVASTATIN SODIUM | 80 MG |
| 42549049090 | PRAVASTATIN SODIUM | 80 MG |
| 50090278500 | PRAVASTATIN SODIUM | 80 MG |
| 50090278501 | PRAVASTATIN SODIUM | 80 MG |
| 50090320400 | PRAVASTATIN SODIUM | 80 MG |
| 50090320401 | PRAVASTATIN SODIUM | 80 MG |
| 54569651000 | PRAVASTATIN SODIUM | 80 MG |
| 54569651001 | PRAVASTATIN SODIUM | 80 MG |
| 54868463400 | PRAVACHOL | 80 MG |
| 54868557900 | PRAVASTATIN SODIUM | 80 MG |
| 54868557901 | PRAVASTATIN SODIUM | 80 MG |
| 55048059830 | PRAVASTATIN SODIUM | 80 MG |
| 55111027405 | PRAVASTATIN SODIUM | 80 MG |
| 55111027490 | PRAVASTATIN SODIUM | 80 MG |
| 57237016705 | PRAVASTATIN SODIUM | 80 MG |
| 57237016790 | PRAVASTATIN SODIUM | 80 MG |
| 60429037005 | PRAVASTATIN SODIUM | 80 MG |
| 60429037045 | PRAVASTATIN SODIUM | 80 MG |
| 60429037090 | PRAVASTATIN SODIUM | 80 MG |
| 60505132305 | PRAVASTATIN SODIUM | 80 MG |
| 60505132309 | PRAVASTATIN SODIUM | 80 MG |
| 61919073490 | PRAVASTATIN SODIUM | 80 MG |
| 63304059805 | PRAVASTATIN SODIUM | 80 MG |
| 63304059890 | PRAVASTATIN SODIUM | 80 MG |
| 68084074625 | PRAVASTATIN SODIUM | 80 MG |
| 68084074695 | PRAVASTATIN SODIUM | 80 MG |
| 68180048802 | PRAVASTATIN SODIUM | 80 MG |
| 68180048809 | PRAVASTATIN SODIUM | 80 MG |
| 68258601303 | PRAVASTATIN SODIUM | 80 MG |
| 68258601309 | PRAVASTATIN SODIUM | 80 MG |
| 68382007305 | PRAVASTATIN SODIUM | 80 MG |
| 68382007316 | PRAVASTATIN SODIUM | 80 MG |
| 68462019805 | PRAVASTATIN SODIUM | 80 MG |
| 68462019890 | PRAVASTATIN SODIUM | 80 MG |
| 68788719301 | PRAVASTATIN SODIUM | 80 MG |
| 68788719302 | PRAVASTATIN SODIUM | 80 MG |
| 68788719303 | PRAVASTATIN SODIUM | 80 MG |
| 68788719306 | PRAVASTATIN SODIUM | 80 MG |
| 68788719308 | PRAVASTATIN SODIUM | 80 MG |
| 68788719309 | PRAVASTATIN SODIUM | 80 MG |
| 68788731601 | PRAVASTATIN SODIUM | 80 MG |
| 68788731602 | PRAVASTATIN SODIUM | 80 MG |
| 68788731603 | PRAVASTATIN SODIUM | 80 MG |
| 68788731606 | PRAVASTATIN SODIUM | 80 MG |
| 68788731608 | PRAVASTATIN SODIUM | 80 MG |
| 68788731609 | PRAVASTATIN SODIUM | 80 MG |
| 00006054328 | ZOCOR | 80 MG |
| 00006054331 | ZOCOR | 80 MG |
| 00006054354 | ZOCOR | 80 MG |
| 00006054361 | ZOCOR | 80 MG |
| 00006054382 | ZOCOR | 80 MG |
| 00093715610 | SIMVASTATIN | 80 MG |
| 00093715619 | SIMVASTATIN | 80 MG |
| 00093715656 | SIMVASTATIN | 80 MG |
| 00093715693 | SIMVASTATIN | 80 MG |
| 00093715698 | SIMVASTATIN | 80 MG |
| 00406206903 | SIMVASTATIN | 80 MG |
| 00406206905 | SIMVASTATIN | 80 MG |
| 00406206910 | SIMVASTATIN | 80 MG |
| 00406206960 | SIMVASTATIN | 80 MG |
| 00406206990 | SIMVASTATIN | 80 MG |
| 00781507431 | SIMVASTATIN | 80 MG |
| 00781507492 | SIMVASTATIN | 80 MG |
| 16252050930 | SIMVASTATIN | 80 MG |
| 16252050950 | SIMVASTATIN | 80 MG |
| 16252050990 | SIMVASTATIN | 80 MG |
| 16590072690 | SIMVASTATIN | 80 MG |
| 16714068501 | SIMVASTATIN | 80 MG |
| 16714068502 | SIMVASTATIN | 80 MG |
| 16714068503 | SIMVASTATIN | 80 MG |
| 16729000710 | SIMVASTATIN | 80 MG |
| 16729000715 | SIMVASTATIN | 80 MG |
| 16729000717 | SIMVASTATIN | 80 MG |
| 21695074230 | SIMVASTATIN | 80 MG |
| 21695074290 | SIMVASTATIN | 80 MG |
| 23490935703 | SIMVASTATIN | 80 MG |
| 23490935706 | SIMVASTATIN | 80 MG |
| 23490935709 | SIMVASTATIN | 80 MG |
| 24658021410 | SIMVASTATIN | 80 MG |
| 24658021430 | SIMVASTATIN | 80 MG |
| 24658021445 | SIMVASTATIN | 80 MG |
| 24658021490 | SIMVASTATIN | 80 MG |
| 24658030410 | SIMVASTATIN | 80 MG |
| 24658030415 | SIMVASTATIN | 80 MG |
| 24658030430 | SIMVASTATIN | 80 MG |
| 24658030445 | SIMVASTATIN | 80 MG |
| 24658030490 | SIMVASTATIN | 80 MG |
| 31722051490 | SIMVASTATIN | 80 MG |
| 35356060030 | SIMVASTATIN | 80 MG |
| 42254016890 | SIMVASTATIN | 80 MG |
| 42571008005 | SIMVASTATIN | 80 MG |
| 42571008090 | SIMVASTATIN | 80 MG |
| 43063008030 | SIMVASTATIN | 80 MG |
| 43063008090 | SIMVASTATIN | 80 MG |
| 43063073330 | SIMVASTATIN | 80 MG |
| 45802029265 | SIMVASTATIN | 80 MG |
| 45802029275 | SIMVASTATIN | 80 MG |
| 50090112100 | SIMVASTATIN | 80 MG |
| 50090112101 | SIMVASTATIN | 80 MG |
| 50268071611 | SIMVASTATIN AVPAK | 80 MG |
| 50268071615 | SIMVASTATIN AVPAK | 80 MG |
| 50742014010 | SIMVASTATIN | 80 MG |
| 52343002545 | SIMVASTATIN | 80 MG |
| 52343002590 | SIMVASTATIN | 80 MG |
| 54569564000 | ZOCOR | 80 MG |
| 54569611300 | SIMVASTATIN | 80 MG |
| 54569611301 | SIMVASTATIN | 80 MG |
| 54868418100 | ZOCOR | 80 MG |
| 54868418101 | ZOCOR | 80 MG |
| 54868563000 | SIMVASTATIN | 80 MG |
| 54868563001 | SIMVASTATIN | 80 MG |
| 55048077630 | SIMVASTATIN | 80 MG |
| 55048077690 | SIMVASTATIN | 80 MG |
| 55111026805 | SIMVASTATIN | 80 MG |
| 55111026830 | SIMVASTATIN | 80 MG |
| 55111026890 | SIMVASTATIN | 80 MG |
| 55111075010 | SIMVASTATIN | 80 MG |
| 55111075030 | SIMVASTATIN | 80 MG |
| 55111075090 | SIMVASTATIN | 80 MG |
| 55887031830 | SIMVASTATIN | 80 MG |
| 55887031860 | SIMVASTATIN | 80 MG |
| 55887031890 | SIMVASTATIN | 80 MG |
| 60760001930 | SIMVASTATIN | 80 MG |
| 63304079310 | SIMVASTATIN | 80 MG |
| 63304079330 | SIMVASTATIN | 80 MG |
| 63304079350 | SIMVASTATIN | 80 MG |
| 63304079390 | SIMVASTATIN | 80 MG |
| 65862005430 | SIMVASTATIN | 80 MG |
| 65862005439 | SIMVASTATIN | 80 MG |
| 65862005490 | SIMVASTATIN | 80 MG |
| 65862005499 | SIMVASTATIN | 80 MG |
| 66336098630 | SIMVASTATIN | 80 MG |
| 66336098690 | SIMVASTATIN | 80 MG |
| 68084016501 | SIMVASTATIN | 80 MG |
| 68084051401 | SIMVASTATIN | 80 MG |
| 68084051411 | SIMVASTATIN | 80 MG |
| 68115075930 | ZOCOR | 80 MG |
| 68180046503 | SIMVASTATIN | 80 MG |
| 68180046506 | SIMVASTATIN | 80 MG |
| 68180046509 | SIMVASTATIN | 80 MG |
| 68180048101 | SIMVASTATIN | 80 MG |
| 68180048102 | SIMVASTATIN | 80 MG |
| 68180048103 | SIMVASTATIN | 80 MG |
| 68382006905 | SIMVASTATIN | 80 MG |
| 68382006906 | SIMVASTATIN | 80 MG |
| 68382006910 | SIMVASTATIN | 80 MG |
| 68382006914 | SIMVASTATIN | 80 MG |
| 68382006916 | SIMVASTATIN | 80 MG |
| 68645026354 | SIMVASTATIN | 80 MG |
| 68645047254 | SIMVASTATIN | 80 MG |
| 70377000512 | SIMVASTATIN | 80 MG |
| 70377000514 | SIMVASTATIN | 80 MG |
| 70377000515 | SIMVASTATIN | 80 MG |
| 66582032330 | LIPTRUZET | 80 MG-10 MG |
| 66582032354 | LIPTRUZET | 80 MG-10 MG |
| 00003516811 | PRAVIGARD PAC | 81 MG; 20 MG |
| 00003517311 | PRAVIGARD PAC | 81 MG; 40 MG |
| 00003518311 | PRAVIGARD PAC | 81 MG; 80 MG |

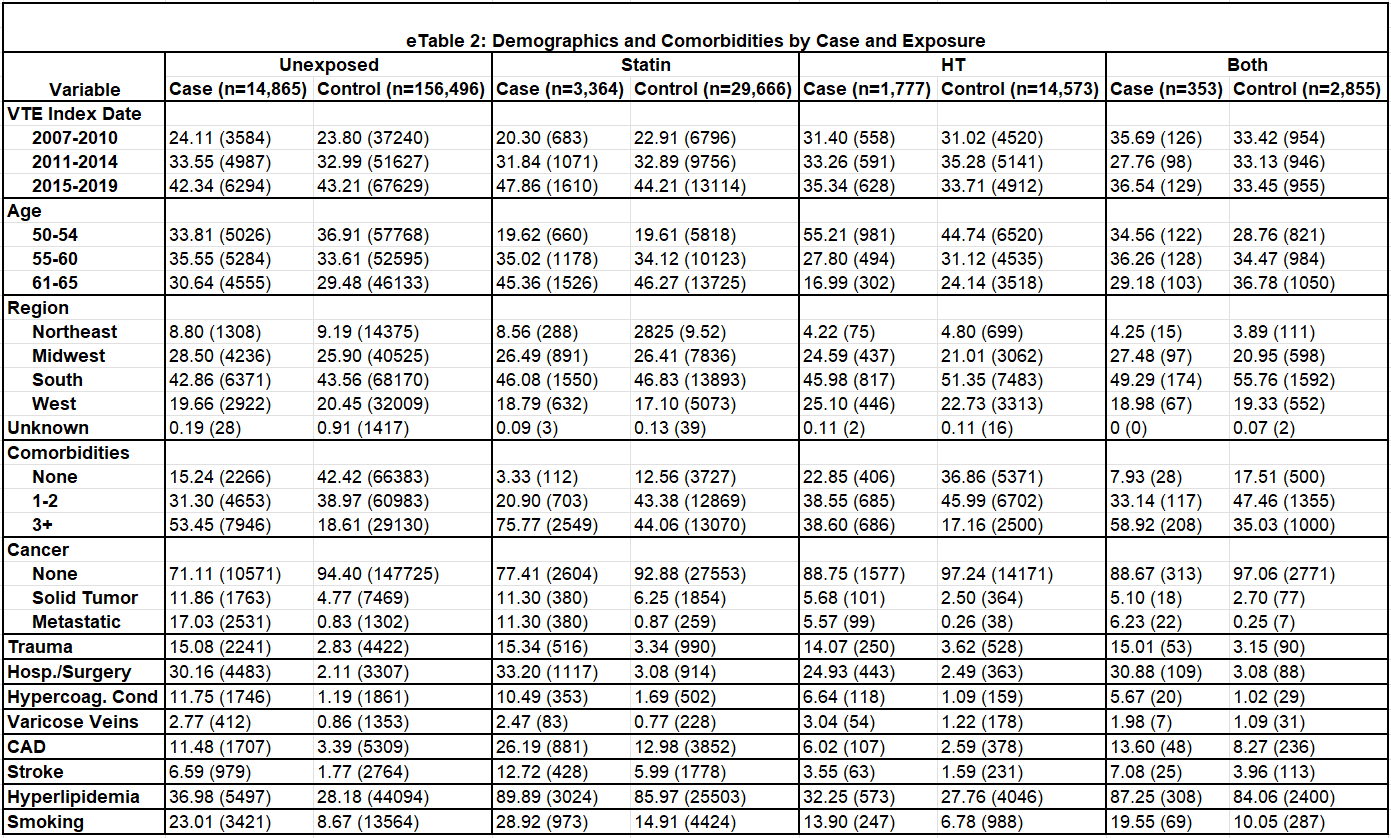
